# The health sector cost of different policy responses to COVID-19 in low- and middle-income countries

**DOI:** 10.1101/2020.08.23.20180299

**Authors:** Sergio Torres-Rueda, Sedona Sweeney, Fiammetta Bozzani, Anna Vassall

## Abstract

Much attention has focussed in recent months on the impact that COVID-19 has on health sector capacity, including critical care bed capacity and resources such as personal protective equipment. However, much less attention has focussed on the overall cost to health sectors, including the full human resource costs and the health system costs to address the pandemic. Here we present estimates of the total costs of COVID-19 response in low- and middle-income countries for different scenarios of COVID-19 mitigation over a one year period. We find costs vary substantially by setting, but in some settings even mitigation scenarios place a substantial fiscal impact on the health system. We conclude that the choices facing many low- and middle-income countries, without further rapid emergency financial support, are stark, between fully funding an effective COVID-19 reponse or other core essential health services.

This is preliminary report that has not yet been peer reviewed. These estimates should not yet guide policy in specific countries nor be reported as established information. We are placing these in the public domain to inform those who are estimaing Covid costs in low- and middle-income countries about the methods and assumptions required; higlight the broad level of fiscal impact and to invite comments for others working in this field, prior to submission to peer review publication.

These estimates have been subjected to a detailed validation and error checking process internally. Nevertheless, given the dearth of data to inform Covid cost estimates at this time, our results depend on a range of assumptions. Comments and suggestions to improve this work are welcomed and can be sent to the authors below

## Introduction

Since the declaration of Coronavirus Disease 2019 (COVID-19) as a public health emergency of international concern by the World Health Organization (WHO) in January 2020 [1], the disease has affected more than 22 million people and caused nearly 290,000 deaths worldwide [2]. While only a minority of cases will experience severe (15%) or critical (5%) disease that requires hospitalisation [3], the estimated costs of implementing WHO pandemic response guidelines in low- and middle-income countries (LMICs) are potentially substantial [4]. These estimates are all the more staggering in the context of over-stretched LMIC health systems, whose capacity to expand the provision of critical care was already limited before the COVID-19 pandemic [5, 6].

While there are financial costing tools available for countries to plan their short term resource requirements [7–9], there are few published estimates of the medium-term economic costs of prevention, tracing and treatment of COVID-19 cases for different epidemic scenarios to inform policy choice and longer term investment. Reliable economic cost estimates are essential for economic evaluations and priority setting around COVID-19 interventions. They can be used for example, to estimate potential cost savings from new COVID-19 vaccines as these become available’ as well as for informing policy choices seeking to balance the broader macro-economic costs of mitigation strategies against the costs to the health system. Although to date, much of the policy concern rightly focuses on the sizeable health and macro-economic impact of COVID-19 in LMICs, it is important that the health sector costs of COVID-19 are also considered as the opportunity costs of the health sector response to COVID-19 on other health areas may be high [10, 11].

Most available cost data to inform these analyses is from high-income countries and/or does not encompass the full COVID-19 response cascade, nor the underlying health system costs [12–14]. To date, there is only one set of economic cost estimates from Kenya that takes into account low-cost case management scenarios appropriate to resource-constrained LMIC settings [15]. This paper presents the first study aiming to project the economic burden of COVID-19 response to LMIC health systems under different pandemic mitigation scenarios in the medium term.

## Methods

### Epidemiological scenarios

Our estimates of COVID-19 cases for different scenarios come from the epidemiological model produced by the London School of Hygiene and Tropical Medicine, estimating the health impact of COVID-19 for 92 LMICs (https://cmmid.github.io/topics/covid19/LMIC-proiection-reports.html). For each country, the model produces estimates on the number of cases, hospitalisations, number of days in hospital for severe cases (general ward) and critical cases (intensive care unit), and deaths for 57 distinct epidemiological scenarios over a one year period [16].

For the costing, four epidemiological scenarios were chosen out of the set of 57 possible scenarios. Scenario 1 represents an unmitigated epidemic. The other three scenarios were chosen because they represent a range of plausible policy options from highly mitigated to more moderate approaches (scenarios 4,14 and 49). Descriptions of the scenarios are presented below in Table 1. Further details can be found in the references above, and the numbers of cases, hospitalisations and bed-days for each country in Supplement S1.

**Table 1.**
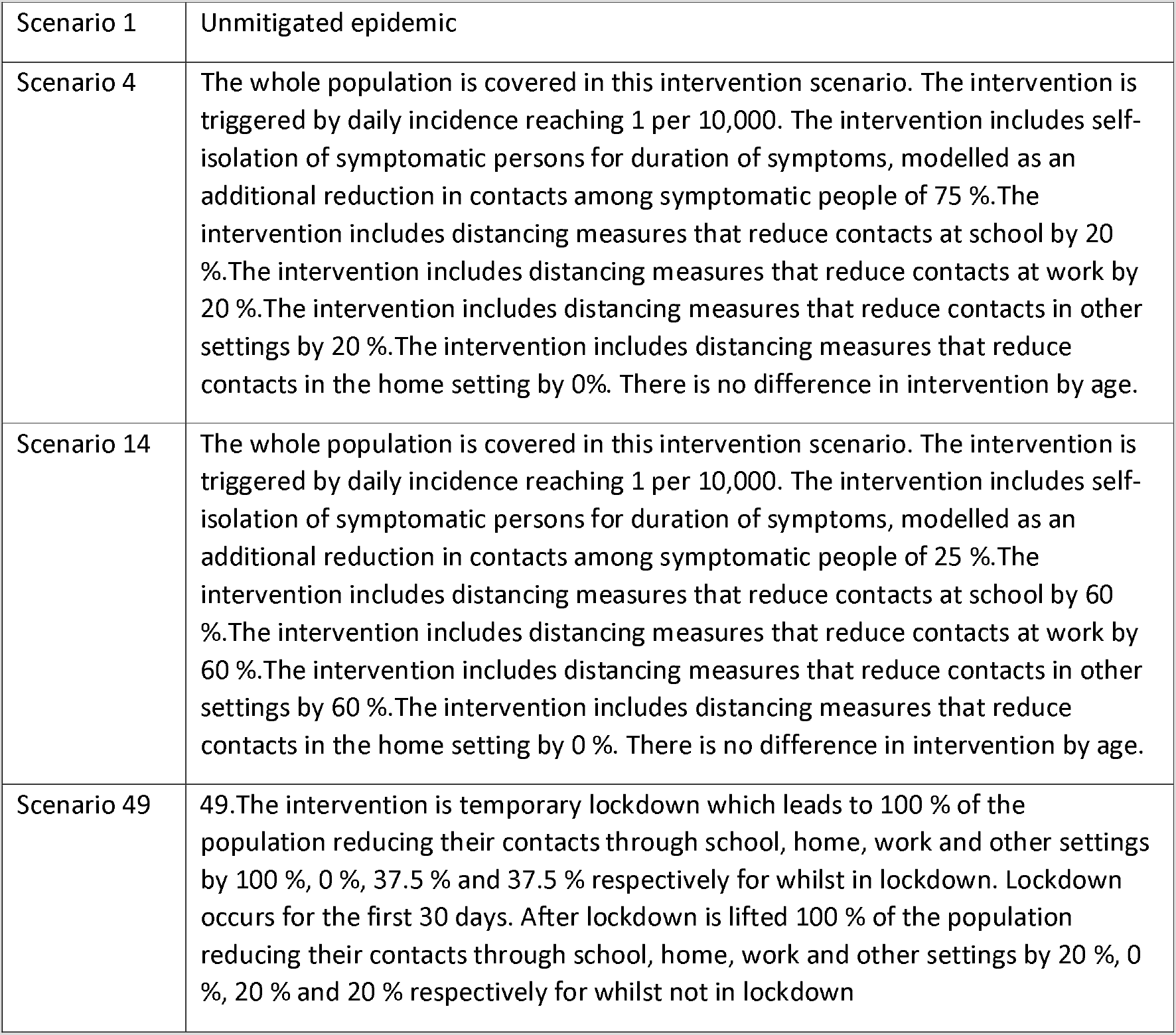
Scenario Descriptions

## Defining the COVID-19 health sector response

In line with the World Health Organisation we costed six priority areas of health sector response COVID-19: a) emergency response mechanisms at the national level; b) risk communication and community engagement; c) case finding, contact tracing and management; d) surveillance; e) public health measures; and, f) case management. For each of these we estimated a unit cost as per Table 2 below.

**Table 2.**
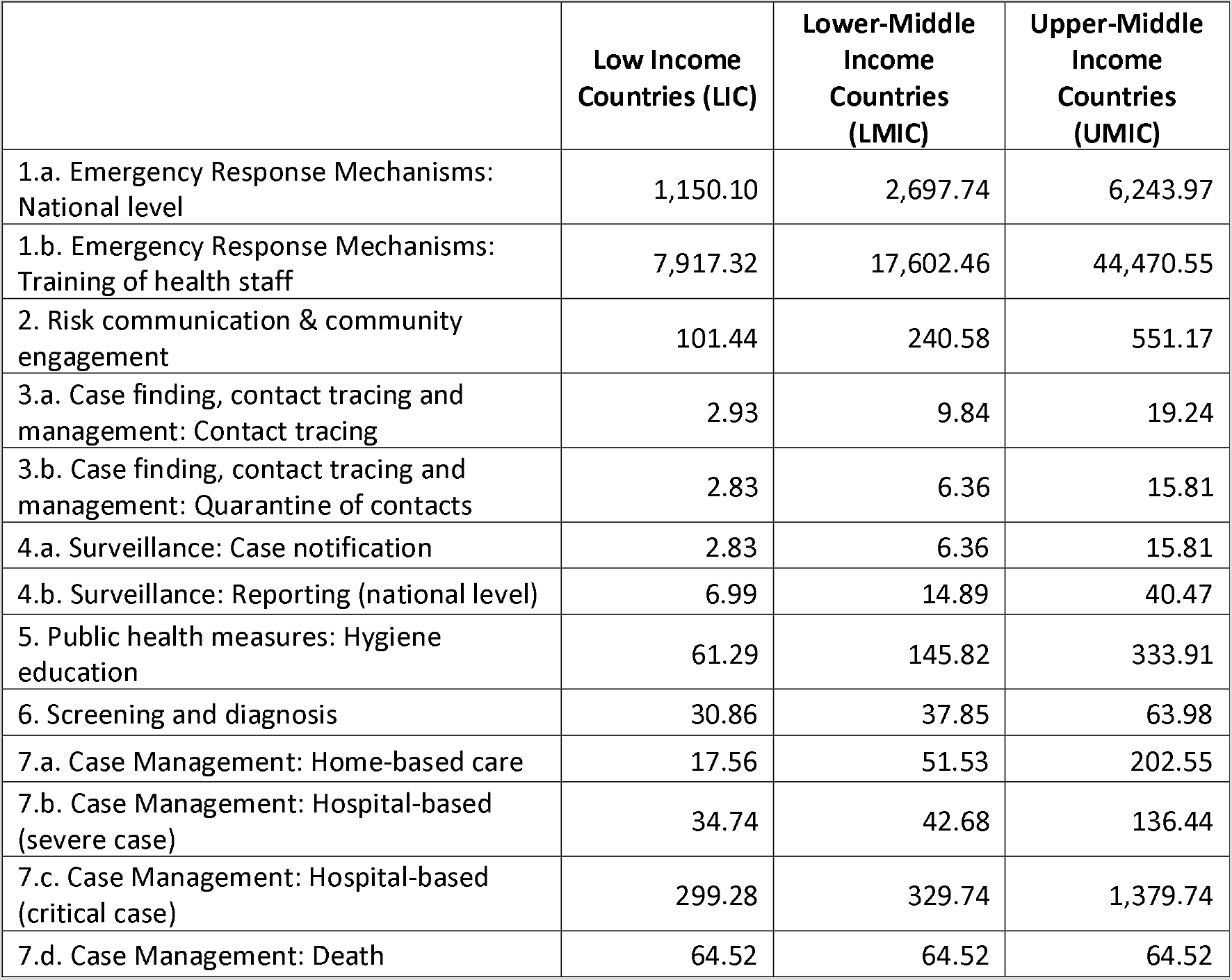
Weighted average (mean) unit cost per activity by country income category (2019 US$)

|  | **Low Income Countries (LIC)** | **Lower-Middle Income Countries (LMIC)** | **Upper-Middle Income Countries (UMIC)** |
| --- | --- | --- | --- |
| 1.a. Emergency Response Mechanisms: National level | 1,150.10 | 2,697.74 | 6,243.97 |
| 1.b. Emergency Response Mechanisms: Training of health staff | 7,917.32 | 17,602.46 | 44,470.55 |
| 2. Risk communication & community engagement | 101.44 | 240.58 | 551.17 |
| 3.a. Case finding, contact tracing and management: Contact tracing | 2.93 | 9.84 | 19.24 |
| 3.b. Case finding, contact tracing and management: Quarantine of contacts | 2.83 | 6.36 | 15.81 |
| 4.a. Surveillance: Case notification | 2.83 | 6.36 | 15.81 |
| 4.b. Surveillance: Reporting (national level) | 6.99 | 14.89 | 40.47 |
| 5. Public health measures: Hygiene education | 61.29 | 145.82 | 333.91 |
| 6. Screening and diagnosis | 30.86 | 37.85 | 63.98 |
| 7.a. Case Management: Home-based care | 17.56 | 51.53 | 202.55 |
| 7.b. Case Management: Hospital-based (severe case) | 34.74 | 42.68 | 136.44 |
| 7.c. Case Management: Hospital-based (critical case) | 299.28 | 329.74 | 1,379.74 |
| 7.d. Case Management: Death | 64.52 | 64.52 | 64.52 |

Our aim was to estimate a ‘real world’ cost that reflects local prices and resource use, and models of service delivery that are considered feasibile given resource constraints in low- and middle-income countries, rather than a full normative costing of national or global guidelines.

To estimate the activity level average (unit) costs for each of the above we used an ingredients based costing approach. Our basis for prices and resource use was recent local cost and resource use data from three countries: Ethiopia (low income country), Pakistan (lower middle income country, and South Africa (upper middle income country). We selected these countries as we were not able to collect primary cost data directly from COVID-19 service delivery points, so chose countries where we had recently conducted large scale costing exercises around either tuberculosis (TB) or general health services. This gave us current local data on actual resource use, input prices and health system unit cost data for services such as outpatient consultations, inpatient bed-days, a range of laboratory tests, including PCR tests and contact tracing. In the case of Ethiopia and South Africa, we had recent data from TB studies. In the case of Pakistan, the research team had been working with the Ministry of Health in 2019/20 to collect ingredients costs for all essential services as part of the Disease Control Priorities Project (DCP). While not primary data collection, all costs were subjected to a review by technical working groups as part of DCP that included practitioners at all levels of the health system. Members of the working groups were asked to consider feasibility as they reviewed and adapted the costs of different essential services.

To adapt this prior data on costs to COVID-19, we then used a combination of: review of COVID-19 resource planning tools and budgets, detailed expert consultation both with international and local experts and literature searches on primary cost data collection on the costs of clinical care in LMICs. Clinical management costs were based on expert consultation on essential critical care, including a detailed estimation of oxygen therapy needs; and considering recent cost data collection on critical care in LMICs^20^; and, length of stay and prevalence of complications from China. While we had access and reviewed length of stay data from different LMICs, this revealed either exceptionally high (early cases) or low (during surge) lengths of stay, and therefore the early data from China across the whole epidemic were felt to be the best estimate of a length of stay (considering those who die during care). Supplement S1 contains the summary level units used for each activity and the unit costs of each, and all assumptions used, including a comparison between normative recommendations for clinical case management and the assumptions used in this costing, based on expert consultation of feasibilty.

We then extrapolated the detailed ingredients costing done for Ethiopia, Pakistan and South Africa to low, lower middle, and upper middle income countries respectively. Each cost input in the ingredients costing was classified as a tradeable good, non-tradeable good, or staff cost. To convert the tradeable good from the base country (e.g. Ethiopia) to a ‘second’ country (e.g. Afghanistan) we apportioned the percentage of the unit cost that was composed of tradeable goods in 2019 US$ from the base country to the second country.

Non-tradeable goods include buildings, heavy machinery, and other equipment. To convert these costs from a base country to a second country we used purchasing power parity (PPP) conversion rates. We multiplied the proportion of the unit cost that was defined as non-tradeable (in 2019 US$) by the ratio of the 2019 GDP per capita (adjusted for PPP) of the second county, divided by the 2019 GDP per capita (adjusted for PPP) of the base country. Data on GDP per capita (adjusted for PPP) can be found in the World Bank database [17].

To convert staff costs from a base country to a second country we used conversion rates from Serje et al (2018) [18]. Serje et al (2018) use regression analysis on a dataset containing wages from health workers of different skill levels for 193 countries in order to predict wages by country income level relative to GDP per capita. We used the GDP per capita multipliers presented in the paper in order to convert the staff wages from the base country to the second country.

Finally, we then estimate the total cost of each scenario, against the % of government and total health expenditure in each country and gross domestic product.

## Results

Our combined primary data and expert reviewed ingreidents costs of managing a mild case with home visits from health care providers ranged from $13 (Pakistan) to $147 (South Africa) per case. Hospital-based care for a severe case ranged from $33.32 (Pakistan) to $106 (South Africa) per day, while costs for a critical case were much higher at $221 (Pakistan) – $1,082 (South Africa) per day in hospital (see supplmentary results appendix Table SR1). Table 2 shows our estimates of the mean unit cost per activity when these costs are extrapolated for low-income, lower-middle income, and upper-middle income countries. Table 2 includes the costs of managing the national emergency response mechanisms, risk communication and community engagement, case finding and surveillance, public health measures, screening and diagnosis, and case management.

Average cost per capita by country income category are reported in Table 3, considering the total number of cases, and other units. Table SR2 and SR3 show the underlying costs per capita and total estimated annual costs per country, by intervention scenario. For example, an unmitigated epidemic would cost an average of $5.2 billion per country, ranging from $11.3 million in Sao Tome and Principe to $127.8 billion in India. This is equivalent to an average of $50 per capita in LIC, $62 in LMIC, and $84 in UMIC (Table SR2). A 30-day lockdown would reduce these annual costs to an average of $4.66 billion (10.3M – 115.1B), or $44–$74/capita. An intervention resulting in social distancing of 20% year-round would further reduce costs to 3.84 billion (8.86M–96.08B), or $37–$60/capita. Finally, an intervention leading to generalized social distancing of 60% year-round would to costs that are on average less than half those of the unmitigated epidemic (5.15M–58.56B or $21–36 percapita).

**Table 3.**
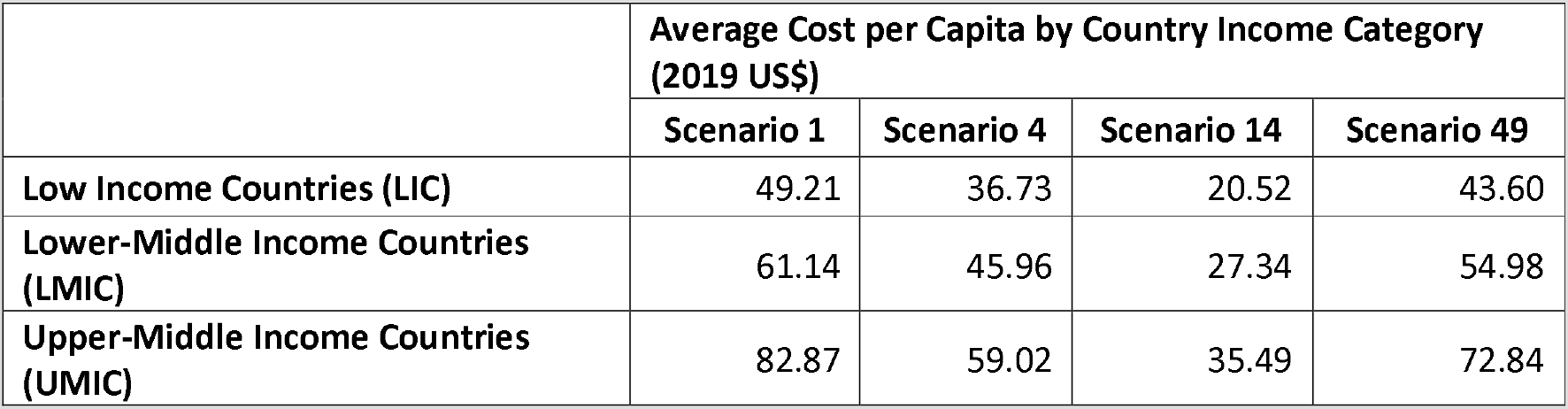
Average Cost per Capita by Country Income Category (2019 US$)

In all scenarios, costs of the COVID-19 response were predominantly attributable to screening and diagnosis and case management, particularly management of critical cases (Table 4). Case finding, contact tracing and surveillance, and public health measures in contrast made up less than 5% of the total response costs. National-level costs of coordinating the emergency response and risk communication likewise constituted a small proportion of overall costs, amounting to less than 1% of total costs on average. The maps in Figures 1a and 1b show the extent to which total costs increase with less stringent social distancing policies, and how social distancing can be more effective than temporary lockdown at containing costs.

**Figure 1a.**
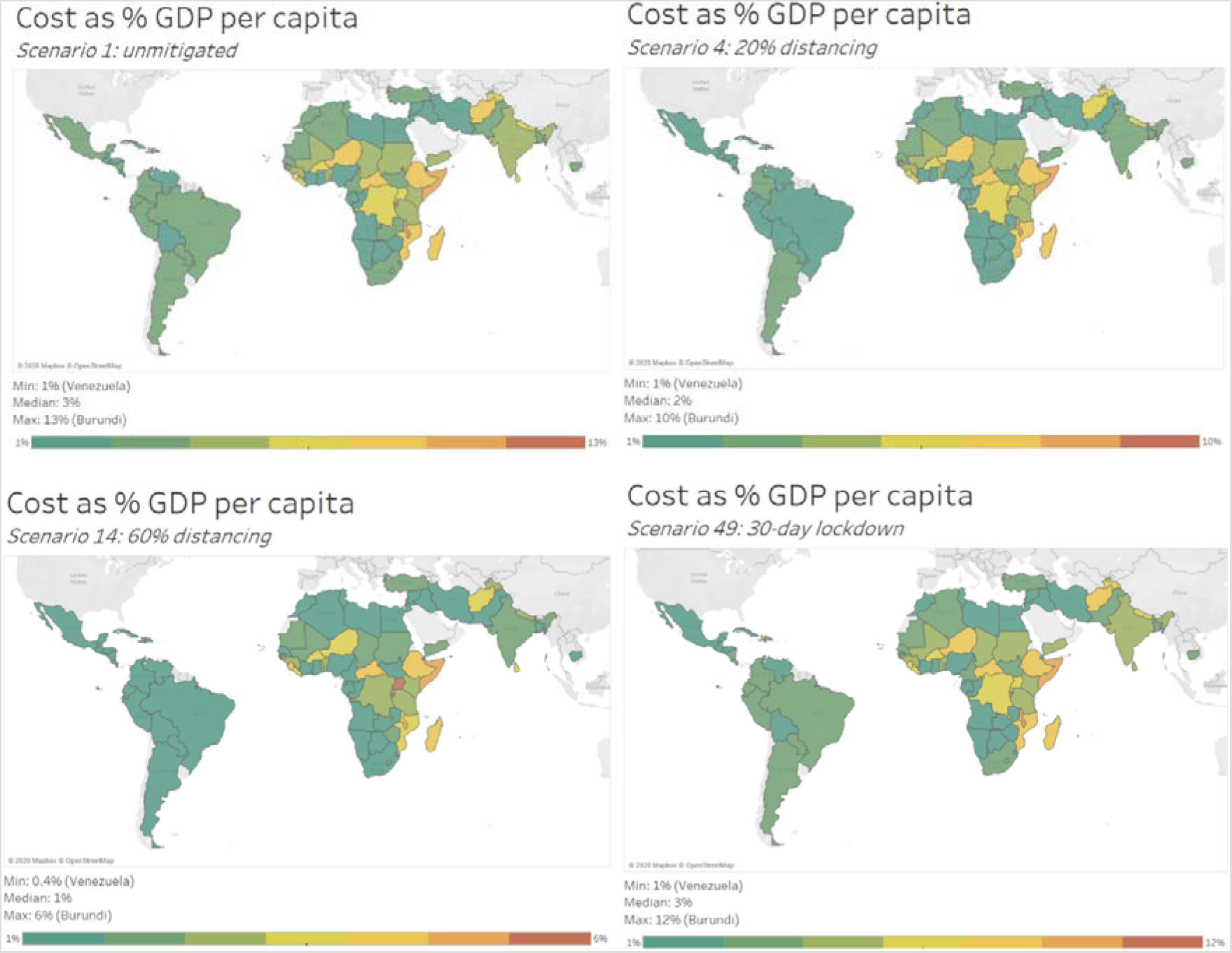
Total costs as a proportion of GDP per capita

**Figure 1b.**
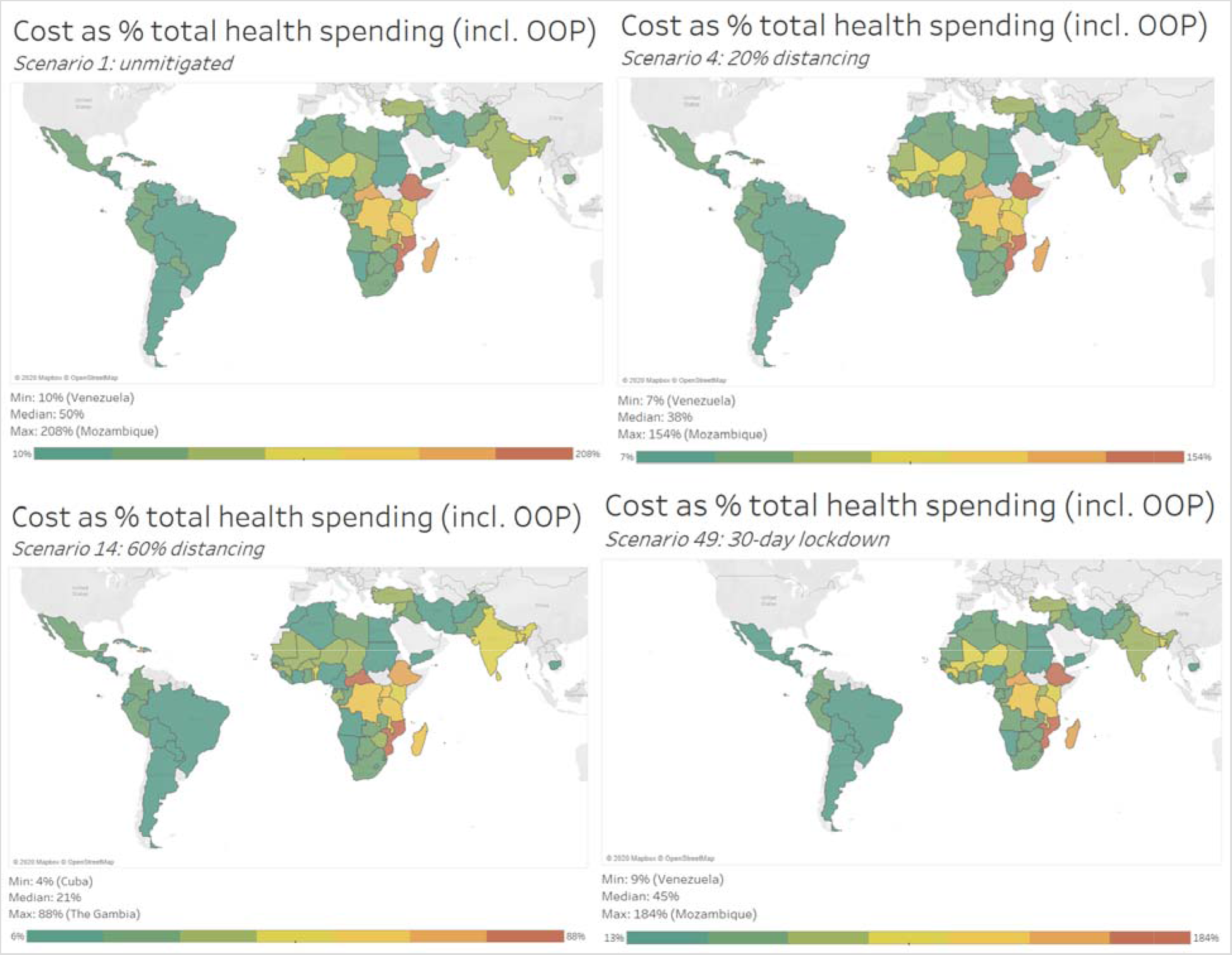
Total costs as a proportion of total health spending, including out-of-pocket payments

**Table 4.** Average % of Total Costs by Activity by Country Income Category by Scenario

|  | Scenario 1 |  |  | Scenario 4 |  |  | Scenario 14 |  |  | Scenario 49 |  |  |
| --- | --- | --- | --- | --- | --- | --- | --- | --- | --- | --- | --- | --- |
|  | LIC | LMIC | UMIC | LIC | LMIC | UMIC | LIC | LMIC | UMIC | LIC | LMIC | UMIC |
| **1.a. Emergency Response Mechanisms: National level** | 0.03% | 0.01% | 0.02% | 0.04% | 0.02% | 0.02% | 0.07% | 0.03% | 0.04% | 0.03% | 0.01% | 0.02% |
| **1.b. Emergency Response Mechanisms: Training of health staff** | 0.05% | 0.12% | 0.18% | 0.07% | 0.16% | 0.27% | 0.13% | 0.27% | 0.44% | 0.06% | 0.13% | 0.21% |
| **2. Risk communication & community engagement** | 0.00% | 0.00% | 0.00% | 0.00% | 0.00% | 0.00% | 0.01% | 0.00% | 0.00% | 0.00% | 0.00% | 0.00% |
| **3.a. Case finding, contact tracing and management: Contact tracing** | 0.96% | 2.19% | 1.24% | 0.98% | 2.27% | 1.31% | 0.96% | 2.23% | 1.25% | 0.96% | 2.22% | 1.25% |
| **3.b. Case finding, contact tracing and management: Quarantine of contacts** | 0.93% | 1.17% | 1.04% | 0.95% | 1.21% | 1.10% | 0.93% | 1.17% | 1.05% | 0.93% | 1.18% | 1.06% |
| **4.a. Surveillance: Case notification** | 0.13% | 0.17% | 0.15% | 0.14% | 0.17% | 0.16% | 0.13% | 0.17% | 0.15% | 0.13% | 0.17% | 0.15% |
| **4.b. Surveillance: Reporting (national level)** | 0.00% | 0.00% | 0.01% | 0.00% | 0.01% | 0.01% | 0.01% | 0.01% | 0.02% | 0.00% | 0.01% | 0.01% |
| **5. Public health measures: Hygiene education** | 0.00% | 0.00% | 0.00% | 0.00% | 0.00% | 0.00% | 0.00% | 0.00% | 0.00% | 0.00% | 0.00% | 0.00% |
| **6. Screening and diagnosis** | 58.42% | 48.42% | 25.10% | 59.71% | 50.07% | 26.54% | 58.25% | 48.58% | 25.26% | 58.78% | 48.91% | 25.45% |
| **7.a. Case Management: Home-based care** | 1.19% | 2.37% | 2.56% | 1.22% | 2.45% | 2.72% | 1.19% | 2.40% | 2.59% | 1.20% | 2.39% | 2.60% |
| **7.b. Case Management: Hospital-based (severe case)** | 6.65% | 8.50% | 10.72% | 6.41% | 8.14% | 10.44% | 6.67% | 8.40% | 10.67% | 6.58% | 8.39% | 10.65% |
| **7.c. Case Management: Hospital-based (critical case)** | 31.23% | 36.62% | 58.79% | 30.10% | 35.10% | 57.25% | 31.26% | 36.33% | 58.34% | 30.92% | 36.16% | 58.40% |
| **7.d. Case Management: Death** | 0.40% | 0.43% | 0.19% | 0.39% | 0.41% | 0.18% | 0.40% | 0.42% | 0.18% | 0.40% | 0.42% | 0.18% |

Our sensitivity analysis (presented in Table SR8) shows that our estimates are partricularly sensitive to our assumptions on the number of symptomatic cases tested. Doubling the estimated number of symptomatic cases tested from 10% to 20% increased our ‘unmitigated’ cost estimates from $50-$84/capita to $57-$90/capita. A ten-fold increase in the number of symptomatic cases tested, from 10% to 100%, resulted in nearly doubling our ‘unmitigated’ cost estimates to $121–137 per capita. The effect of this assumption was constant across all scenarios.

## Discussion

We find that the costs to health sector of responding to COVID-19 are substantial in LMICs, even when estimating the costs of essential critical care only. High levels of social distancing, however, may halve these costs within a one year period compared to allowing the pandemic to proceed in an unmitigated manner in the first year. The total cost and the costs as a proportion of health expenditue and GDP vary substantially across countries, with some countries likely to be able to ‘afford’ the costs, and for others the financial impact of COVID-19 on the health sector in its first year being higher than normal annual costs for the health sector in totality.

Our methods are subject to many limitations. Normally we would estimate ‘real world’ costs collecting extensive primary cost data on actual service delivery. In the case of COVID-19, we have not been able to do this. We have therefore had to rely substantially on data collected for other purposes and on expert opinion from LMICs to make key assumptions on how services may be delivered. We aimed to include the total costs to the health system and our aim was to estimate ‘real world costs’ in terms of the resource needs to deliver the most essential care. However, our costs are unlikely to reflect actual expenditures, as countries may either provide more care to specific patients, or struggle to provide even the most essential care given the current restrictions on expenditure. Likewise, the case numbers that our estimates rely upon are unlikely to match the real case numbers currently being observed in many LMICs, as they represent single policy options, and a time period of one year from the start of the epidemic. Finally, we do not include the costs of protecting health care workers delivering other essential services outside the COVID-19 response.

Despite these limitations, our work highlights several critical qualitative recommendations for those working in COVID-19 policy. First, it is imperative, that global agencies and funders continue to act to ensure sufficient resources are made available globally for LMICs to respond, as the epidemic evolves. While much of the focus is on the macro-economic impact of COVID-19 and mortality impact, the fiscal impact on the health sector is likely to be substantial also needs to be considered if health systems in LMICs are to continue to deliver health services effectively. Second, it is clear that some countries are much more vulnerable to fiscal impact on the health sector than others. While specific short term financing needs for COVID-19 will fluctuate considerably over time, we highlight that some countries fundamentally will struggle to cope with almost any COVID-19 scenario, whereas others may be less vulnerable; which could help target support. Finally, our results demonstrate the myriad decisions about care, protection and patient experience that are required to plan resource needs of the response for which there is little discussion or data on what is feasible in LMICs. This is a task that cannot be met using a global perspective, but need country specific input to reflect the specific health system characteristics of each country. We therefore also call for urgent support to encourage interaction of economists, planners, service managers and epidemiological modellers to inform COVID-19 policy at the country level accross LMICs.

## Data Availability

We have included the data in the main text of the report, which contains supplementary material

## Supplemental Results Tables

**Table SR1.**
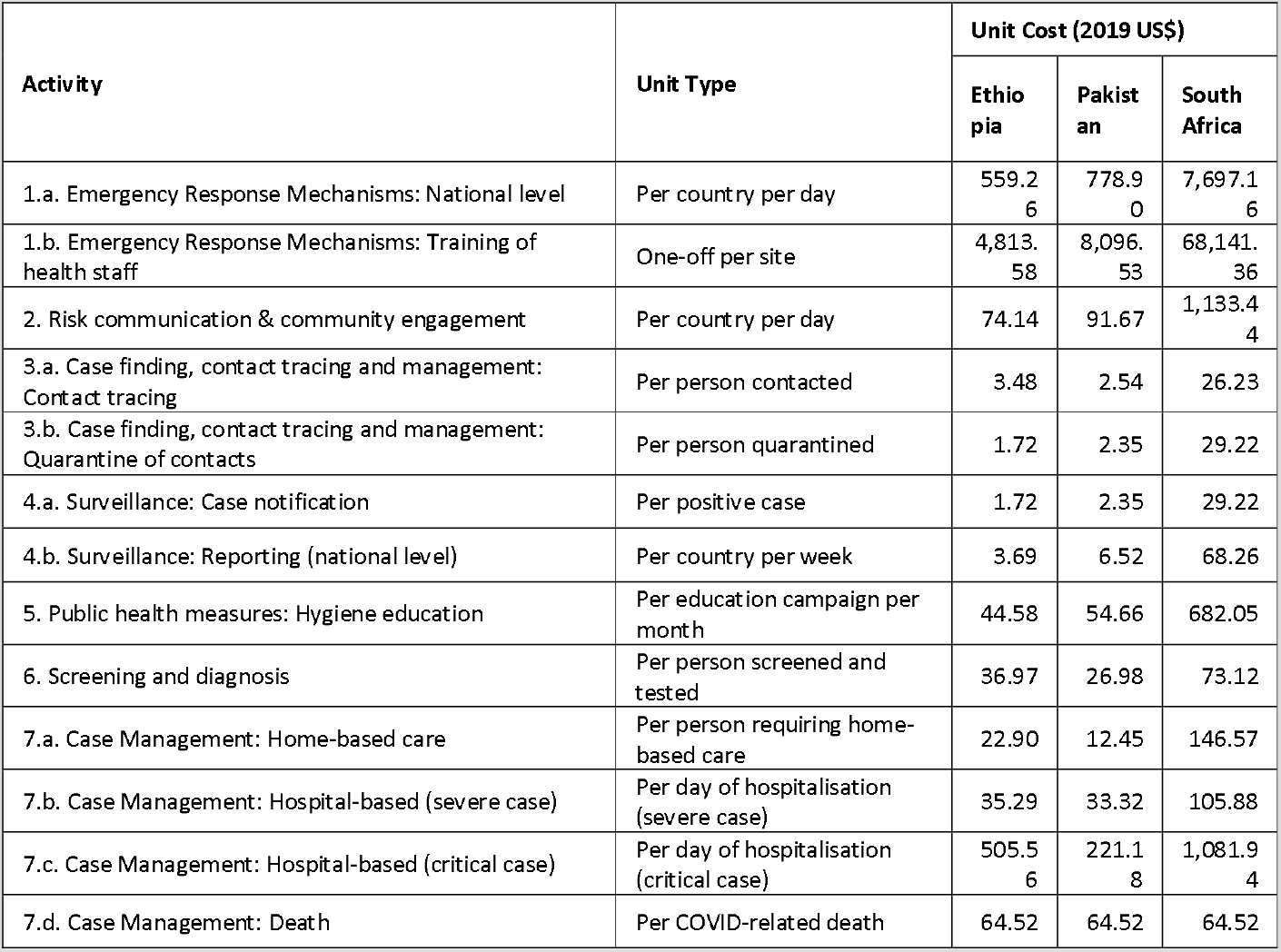
Unit costs per activity for base countries (2019 US$)

**Table SR2:**
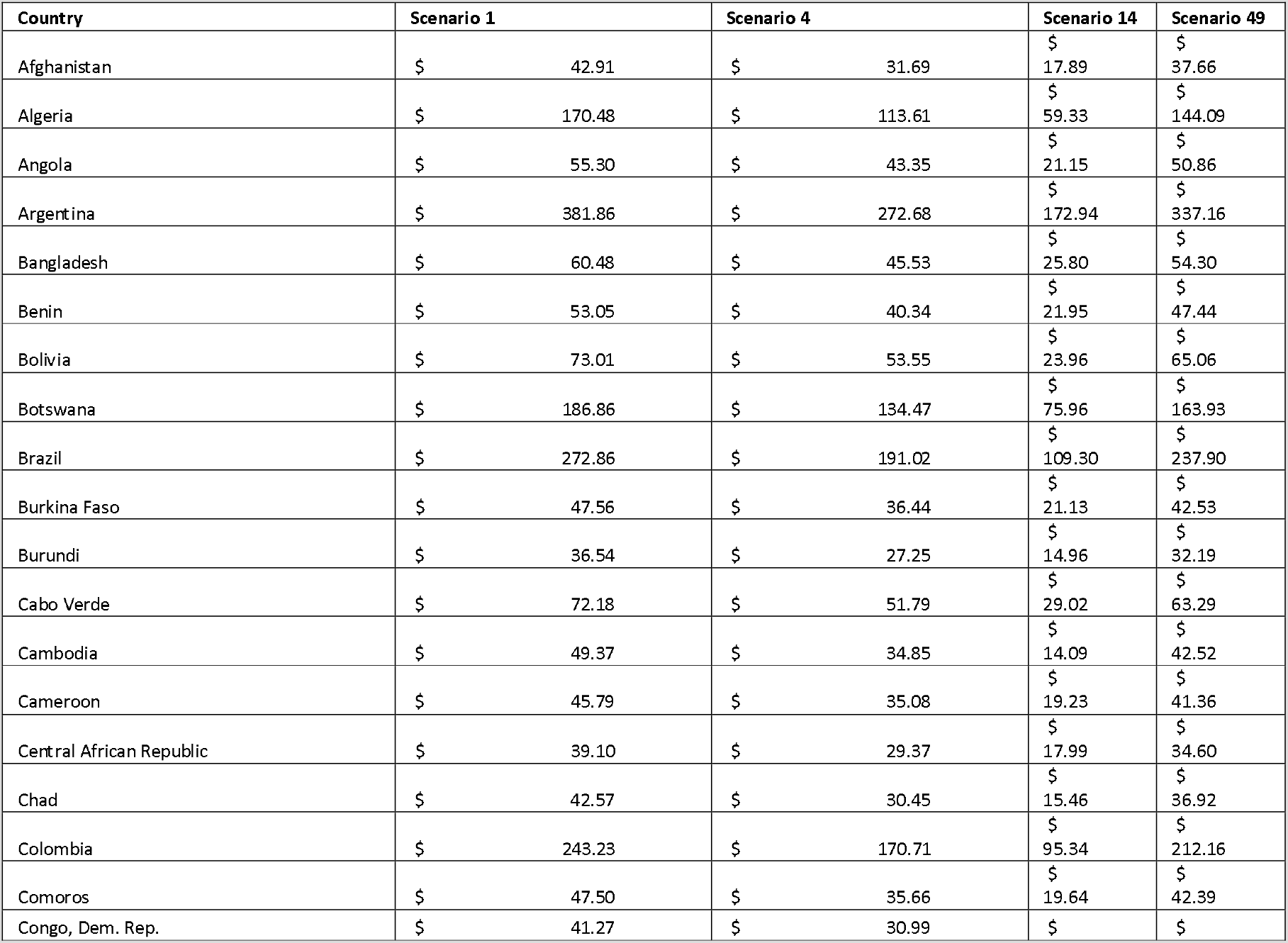
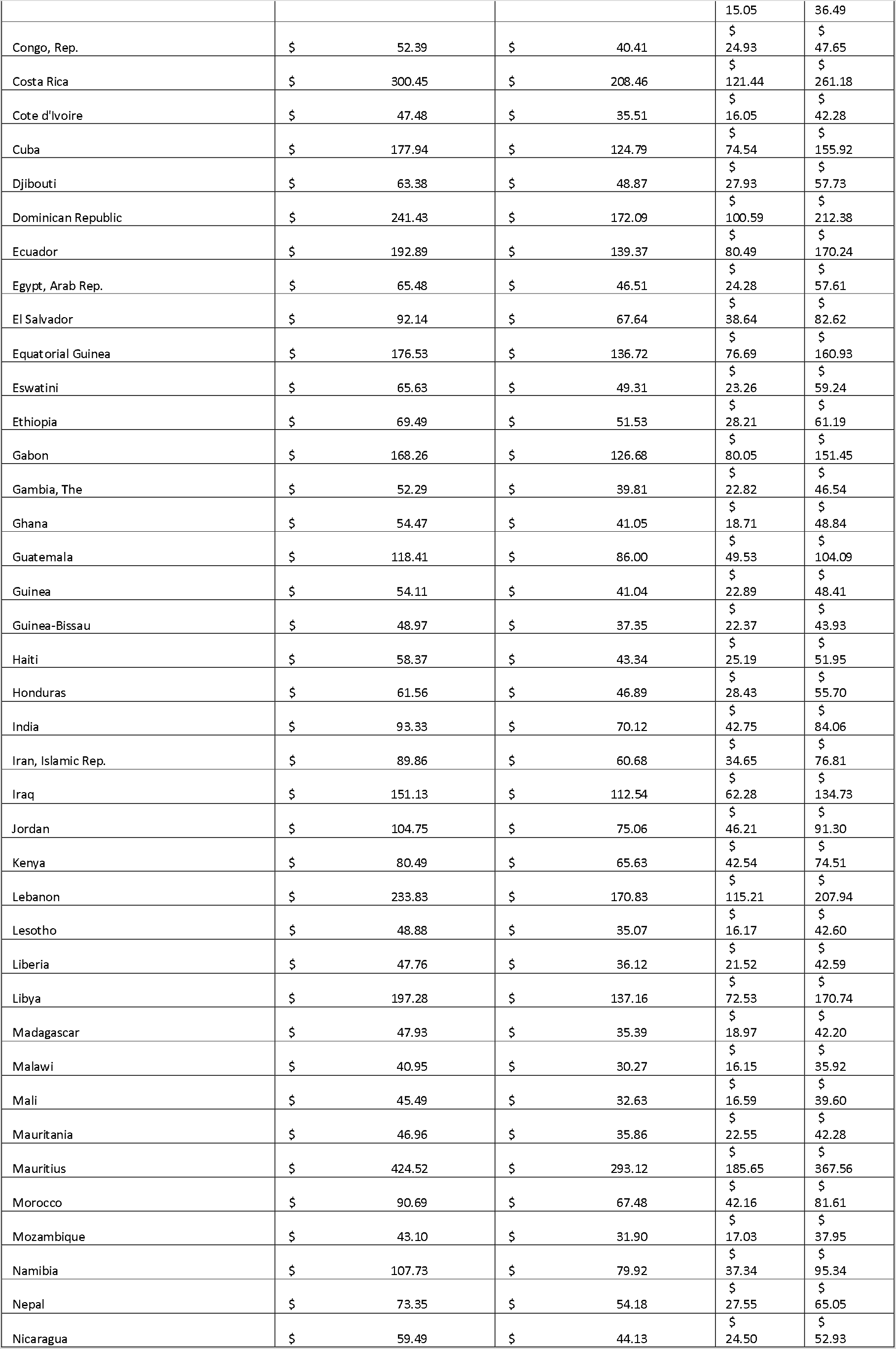
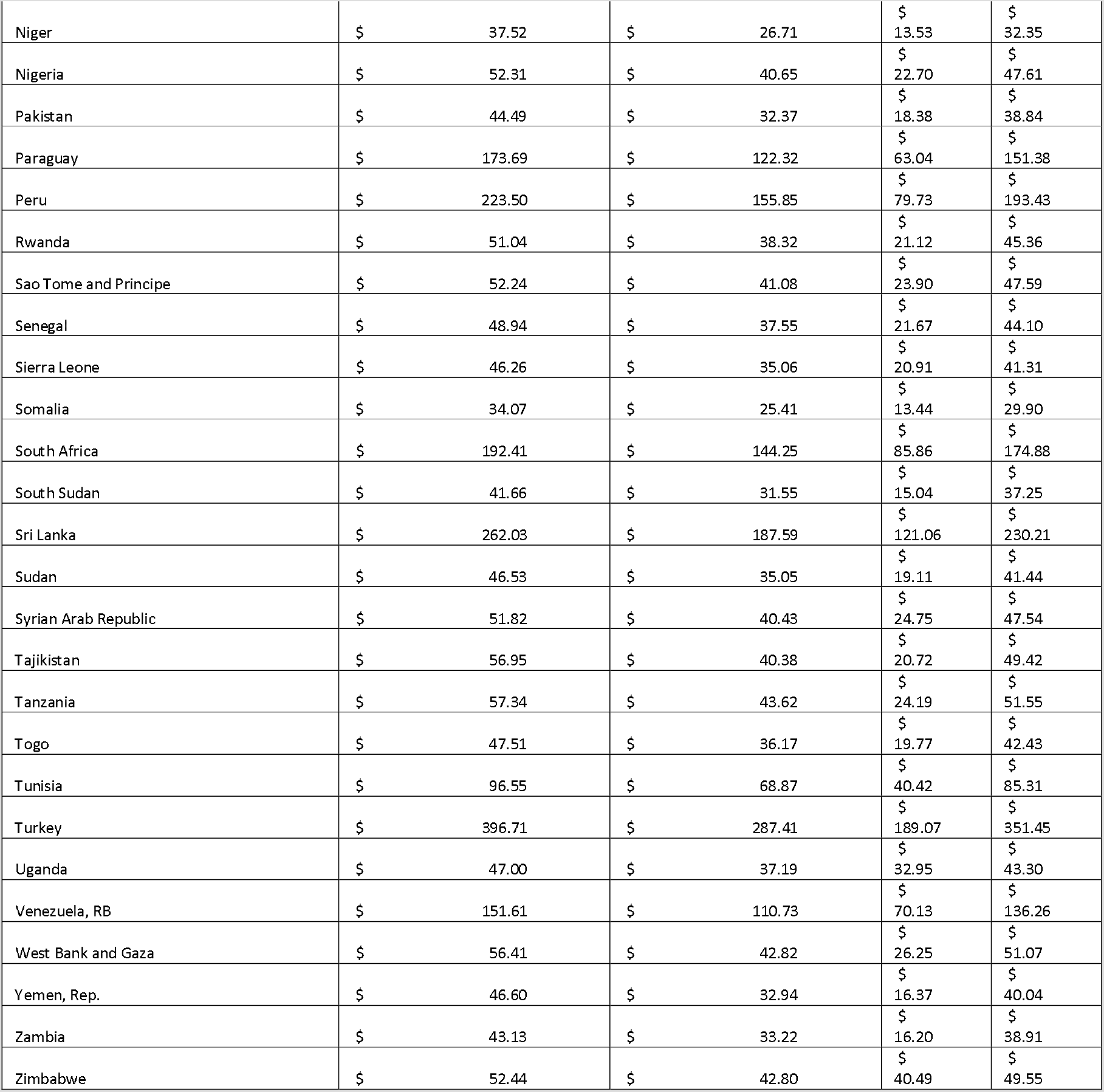
Cost per Capita by Country per Scenario (2019 US$)

**Table SR3.** Total annual costs per country (2019 US$)

| Country | Total Annual Costs per Country (2019 US\$) | | | |
| --- | --- | --- | --- | --- |
|  | Scenario 1 | Scenario 4 | Scenario 14 | Scenario 49 |
| Afghanistan | 1,595,006,688 | 1,177,904,724 | 664,951,614 | 1,399,789,895 |
| Algeria | 7,198,927,335 | 4,797,609,340 | 2,505,281,318 | 6,084,882,367 |
| Angola | 1,703,804,507 | 1,335,741,419 | 651,587,956 | 1,567,094,020 |
| Argentina | 16,990,791,939 | 12,132,966,200 | 7,694,951,524 | 15,001,801,214 |
| Bangladesh | 9,758,429,693 | 7,346,626,409 | 4,162,921,283 | 8,762,131,868 |
| Benin | 609,231,361 | 463,323,321 | 252,112,394 | 544,803,973 |
| Bolivia | 828,941,402 | 607,984,156 | 272,020,642 | 738,627,623 |
| Botswana | 421,205,098 | 303,116,161 | 171,230,132 | 369,522,644 |
| Brazil | 57,156,312,626 | 40,013,304,222 | 22,894,275,352 | 49,832,168,384 |
| Burkina Faso | 939,419,926 | 719,748,836 | 417,278,324 | 840,119,232 |
| Burundi | 408,399,996 | 304,544,873 | 167,162,430 | 359,734,439 |
| Cabo Verde | 39,247,227 | 28,162,920 | 15,780,012 | 34,414,567 |
| Cambodia | 802,267,483 | 566,274,013 | 229,000,547 | 690,985,182 |
| Cameroon | 1,154,599,786 | 884,467,637 | 484,883,954 | 1,043,034,826 |
| Central African Republic | 182,433,804 | 137,035,951 | 83,939,501 | 161,434,468 |
| Chad | 658,884,973 | 471,303,516 | 239,244,463 | 571,410,658 |
| Colombia | 12,075,940,244 | 8,475,322,329 | 4,733,454,628 | 10,533,617,075 |
| Comoros | 39,533,969 | 29,677,244 | 16,343,351 | 35,281,984 |
| Congo, Dem. Rep. | 3,469,615,019 | 2,605,619,705 | 1,265,377,537 | 3,067,476,753 |
| Congo, Rep. | 274,760,274 | 211,912,215 | 130,725,594 | 249,919,561 |
| Costa Rica | 1,502,087,635 | 1,042,161,571 | 607,146,919 | 1,305,750,553 |
| Cote d'Ivoire | 1,190,399,892 | 890,204,132 | 402,280,265 | 1,059,989,337 |
| Cuba | 2,017,494,968 | 1,414,934,979 | 845,099,624 | 1,767,876,988 |
| Djibouti | 60,775,939 | 46,862,116 | 26,784,302 | 55,355,583 |
| Dominican Republic | 2,565,684,173 | 1,828,777,716 | 1,068,959,610 | 2,256,950,992 |
| Ecuador | 3,295,395,303 | 2,380,985,925 | 1,375,043,118 | 2,908,422,164 |
| Egypt, Arab Rep. | 6,445,190,291 | 4,577,530,923 | 2,389,669,661 | 5,670,659,187 |
| El Salvador | 591,579,720 | 434,268,407 | 248,098,581 | 530,457,154 |
| Equatorial Guinea | 231,075,219 | 178,959,540 | 100,391,379 | 210,653,340 |
| Eswatini | 74,567,166 | 56,029,505 | 26,432,806 | 67,309,663 |
| Ethiopia | 7,589,595,244 | 5,627,795,574 | 3,081,548,357 | 6,683,485,989 |
| Gabon | 356,596,194 | 268,464,549 | 169,638,184 | 320,961,919 |
| Gambia, The | 119,218,198 | 90,763,358 | 52,035,644 | 106,107,646 |
| Ghana | 1,621,297,684 | 1,221,891,384 | 556,920,614 | 1,453,846,283 |
| Guatemala | 2,042,320,674 | 1,483,251,542 | 854,210,602 | 1,795,402,698 |
| Guinea | 671,793,449 | 509,527,263 | 284,208,408 | 600,953,264 |
| Guinea-Bissau | 91,777,597 | 70,013,569 | 41,923,342 | 82,337,549 |
| Haiti | 649,227,561 | 482,130,225 | 280,151,525 | 577,852,090 |
| Honduras | 590,251,293 | 449,603,344 | 272,545,259 | 534,058,049 |
| India | 126,241,607,727 | 94,845,714,832 | 57,820,205,064 | 113,697,199,394 |
| Iran, Islamic Rep. | 7,350,914,852 | 4,963,443,606 | 2,834,585,552 | 6,283,486,292 |
| Iraq | 5,808,461,053 | 4,325,177,524 | 2,393,641,505 | 5,178,286,305 |
| Jordan | 1,042,846,812 | 747,309,105 | 460,019,255 | 908,958,207 |
| Kenya | 4,136,465,948 | 3,372,960,785 | 2,186,416,908 | 3,829,322,849 |
| Lebanon | 1,601,468,785 | 1,169,981,518 | 789,069,252 | 1,424,189,709 |
| Lesotho | 103,041,897 | 73,935,288 | 34,089,165 | 89,815,951 |
| Liberia | 230,138,172 | 174,077,339 | 103,699,634 | 205,216,814 |
| Libya | 1,317,562,603 | 916,036,535 | 484,376,095 | 1,140,287,493 |
| Madagascar | 1,258,737,705 | 929,396,816 | 498,242,703 | 1,108,391,195 |
| Malawi | 742,886,055 | 549,184,096 | 293,026,156 | 651,787,586 |
| Mali | 867,812,732 | 622,492,840 | 316,411,256 | 755,421,554 |
| Mauritania | 206,771,925 | 157,899,063 | 99,305,744 | 186,164,959 |
| Mauritius | 537,143,681 | 370,884,267 | 234,900,728 | 465,076,712 |
| Mexico | 34,115,201,144 | 24,625,827,163 | 15,782,810,293 | 30,216,622,750 |
| Morocco | 3,267,515,013 | 2,431,152,726 | 1,519,075,135 | 2,940,380,785 |
| Mozambique | 1,271,233,930 | 940,866,088 | 502,228,251 | 1,119,434,522 |
| Namibia | 263,758,483 | 195,667,209 | 91,410,118 | 233,404,625 |
| Nepal | 2,060,381,058 | 1,521,796,884 | 773,785,915 | 1,827,252,564 |
| Nicaragua | 384,638,289 | 285,335,078 | 158,382,871 | 342,217,217 |
| Niger | 842,039,860 | 599,341,108 | 303,663,836 | 725,945,647 |
| Nigeria | 10,247,169,054 | 7,962,255,548 | 4,445,505,322 | 9,326,514,000 |
| Pakistan | 9,441,146,849 | 6,868,761,344 | 3,901,350,552 | 8,242,469,426 |
| Paraguay | 1,208,234,685 | 850,884,034 | 438,489,428 | 1,053,013,278 |
| Peru | 7,149,625,157 | 4,985,660,537 | 2,550,356,751 | 6,187,688,798 |
| Rwanda | 627,950,026 | 471,398,249 | 259,768,907 | 558,014,561 |
| Sao Tome and Principe | 11,024,075 | 8,669,252 | 5,044,582 | 10,042,513 |
| Senegal | 775,913,554 | 595,256,052 | 343,558,437 | 699,179,799 |
| Sierra Leone | 353,894,747 | 268,216,325 | 159,990,783 | 316,044,465 |
| Somalia | 511,309,434 | 381,354,484 | 201,668,843 | 448,722,521 |
| South Africa | 11,117,131,659 | 8,334,446,288 | 4,960,920,248 | 10,104,268,308 |
| South Sudan | 457,247,980 | 346,307,159 | 165,112,181 | 408,857,053 |
| Sri Lanka | 5,678,100,834 | 4,065,040,082 | 2,623,300,200 | 4,988,596,480 |
| Sudan | 1,944,960,100 | 1,465,299,051 | 798,971,465 | 1,732,370,209 |
| Syrian Arab Republic | 876,059,081 | 683,467,982 | 418,397,444 | 803,740,032 |
| Tajikistan | 518,284,390 | 367,451,171 | 188,545,177 | 449,773,206 |
| Tanzania | 3,229,078,738 | 2,456,724,397 | 1,362,187,767 | 2,903,091,232 |
| Togo | 374,774,447 | 285,340,906 | 155,932,903 | 334,768,272 |
| Tunisia | 1,116,665,762 | 796,503,718 | 467,489,081 | 986,577,100 |
| Turkey | 32,657,064,433 | 23,659,504,785 | 15,564,550,540 | 28,931,632,252 |
| Uganda | 2,007,915,732 | 1,588,879,797 | 1,407,932,788 | 1,849,941,504 |
| Venezuela, RB | 4,377,122,572 | 3,196,922,891 | 2,024,556,702 | 3,933,821,378 |
| West Bank and Gaza | 257,722,952 | 195,636,416 | 119,932,408 | 233,328,972 |
| Yemen, Rep. | 1,328,176,781 | 938,682,001 | 466,480,766 | 1,141,026,798 |
| Zambia | 748,338,033 | 576,387,647 | 281,151,582 | 675,092,378 |
| Zimbabwe | 757,131,633 | 617,966,425 | 584,680,182 | 715,488,782 |

**Table SR4:** Health System Costs of COVID 19 Response per capita as % of GDP per capita (nominal)

| Country | Scenario 1 | Scenario 4 | Scenario 14 | Scenario 49 |
| --- | --- | --- | --- | --- |
| Afghanistan | 8.24% | 6.08% | 3.43% | 7.23% |
| Algeria | 4.14% | 2.76% | 1.44% | 3.50% |
| Angola | 1.61% | 1.26% | 0.62% | 1.48% |
| Argentina | 3.27% | 2.33% | 1.48% | 2.89% |
| Bangladesh | 3.56% | 2.68% | 1.52% | 3.20% |
| Benin | 5.88% | 4.47% | 2.43% | 5.26% |
| Bolivia | 2.06% | 1.51% | 0.68% | 1.83% |
| Botswana | 2.26% | 1.63% | 0.92% | 1.98% |
| Brazil | 3.06% | 2.14% | 1.23% | 2.67% |
| Burkina Faso | 6.65% | 5.10% | 2.95% | 5.95% |
| Burundi | 13.45% | 10.03% | 5.50% | 11.85% |
| Cabo Verde | 1.99% | 1.42% | 0.80% | 1.74% |
| Cambodia | 3.27% | 2.31% | 0.93% | 2.82% |
| Cameroon | 2.99% | 2.29% | 1.25% | 2.70% |
| Central African Republic | 8.22% | 6.17% | 3.78% | 7.27% |
| Chad | 5.84% | 4.18% | 2.12% | 5.07% |
| Colombia | 3.65% | 2.56% | 1.43% | 3.18% |
| Comoros | 3.36% | 2.52% | 1.39% | 3.00% |
| Congo, Dem. Rep. | 7.35% | 5.52% | 2.68% | 6.50% |
| Congo, Rep. | 2.44% | 1.88% | 1.16% | 2.22% |
| Costa Rica | 2.50% | 1.73% | 1.01% | 2.17% |
| Cote d'Ivoire | 2.77% | 2.07% | 0.94% | 2.46% |
| Cuba | 2.02% | 1.41% | 0.84% | 1.77% |
| Djibouti | 2.06% | 1.59% | 0.91% | 1.87% |
| Dominican Republic | 3.00% | 2.14% | 1.25% | 2.64% |
| Ecuador | 3.04% | 2.20% | 1.27% | 2.68% |
| Egypt, Arab Rep. | 2.57% | 1.82% | 0.95% | 2.26% |
| El Salvador | 2.27% | 1.67% | 0.95% | 2.04% |
| Equatorial Guinea | 1.72% | 1.33% | 0.75% | 1.57% |
| Eswatini | 1.58% | 1.19% | 0.56% | 1.43% |
| Ethiopia | 9.00% | 6.67% | 3.65% | 7.92% |
| Gabon | 2.12% | 1.59% | 1.01% | 1.90% |
| Gambia, The | 7.30% | 5.56% | 3.19% | 6.50% |
| Ghana | 2.47% | 1.86% | 0.85% | 2.22% |
| Guatemala | 2.60% | 1.89% | 1.09% | 2.29% |
| Guinea | 6.16% | 4.67% | 2.61% | 5.51% |
| Guinea-Bissau | 6.29% | 4.80% | 2.88% | 5.65% |
| Haiti | 6.72% | 4.99% | 2.90% | 5.98% |
| Honduras | 2.46% | 1.88% | 1.14% | 2.23% |
| India | 4.64% | 3.49% | 2.13% | 4.18% |
| Iran, Islamic Rep. | 1.60% | 1.08% | 0.62% | 1.36% |
| Jordan | 2.47% | 1.77% | 1.09% | 2.15% |
| Kenya | 4.71% | 3.84% | 2.49% | 4.36% |
| Lebanon | 2.83% | 2.07% | 1.39% | 2.51% |
| Lesotho | 3.76% | 2.70% | 1.24% | 3.28% |
| Liberia | 7.05% | 5.33% | 3.18% | 6.29% |
| Libya | 2.72% | 1.89% | 1.00% | 2.36% |
| Madagascar | 9.09% | 6.71% | 3.60% | 8.00% |
| Malawi | 10.52% | 7.77% | 4.15% | 9.23% |
| Mali | 5.06% | 3.63% | 1.84% | 4.40% |
| Mauritania | 3.95% | 3.02% | 1.90% | 3.56% |
| Mauritius | 3.78% | 2.61% | 1.65% | 3.27% |
| Mexico | 2.79% | 2.02% | 1.29% | 2.48% |
| Morocco | 2.80% | 2.08% | 1.30% | 2.52% |
| Mozambique | 8.64% | 6.39% | 3.41% | 7.61% |
| Namibia | 1.82% | 1.35% | 0.63% | 1.61% |
| Nepal | 7.09% | 5.24% | 2.66% | 6.29% |
| Nicaragua | 2.93% | 2.18% | 1.21% | 2.61% |
| Niger | 9.06% | 6.45% | 3.27% | 7.81% |
| Nigeria | 2.58% | 2.00% | 1.12% | 2.35% |
| Pakistan | 3.00% | 2.18% | 1.24% | 2.62% |
| Paraguay | 2.98% | 2.10% | 1.08% | 2.60% |
| Peru | 3.22% | 2.25% | 1.15% | 2.79% |
| Rwanda | 6.60% | 4.96% | 2.73% | 5.87% |
| Sao Tome and Principe | 2.61% | 2.05% | 1.19% | 2.38% |
| Senegal | 3.22% | 2.47% | 1.42% | 2.90% |
| Sierra Leone | 8.66% | 6.57% | 3.92% | 7.74% |
| South Africa | 3.02% | 2.26% | 1.35% | 2.74% |
| Sri Lanka | 6.39% | 4.57% | 2.95% | 5.61% |
| Sudan | 4.76% | 3.59% | 1.96% | 4.24% |
| Syrian Arab Republic | 2.55% | 1.99% | 1.22% | 2.34% |
| Tajikistan | 6.89% | 4.88% | 2.51% | 5.98% |
| Tanzania | 5.46% | 4.15% | 2.30% | 4.91% |
| Togo | 6.99% | 5.32% | 2.91% | 6.25% |
| Tunisia | 2.80% | 2.00% | 1.17% | 2.47% |
| Turkey | 4.23% | 3.07% | 2.02% | 3.75% |
| Venezuela, RB | 0.94% | 0.69% | 0.44% | 0.85% |
| Yemen, Rep. | 4.93% | 3.49% | 1.73% | 4.24% |
| Zambia | 2.80% | 2.16% | 1.05% | 2.53% |
| Zimbabwe | 2.44% | 1.99% | 1.89% | 2.31% |

**Table SR5:**
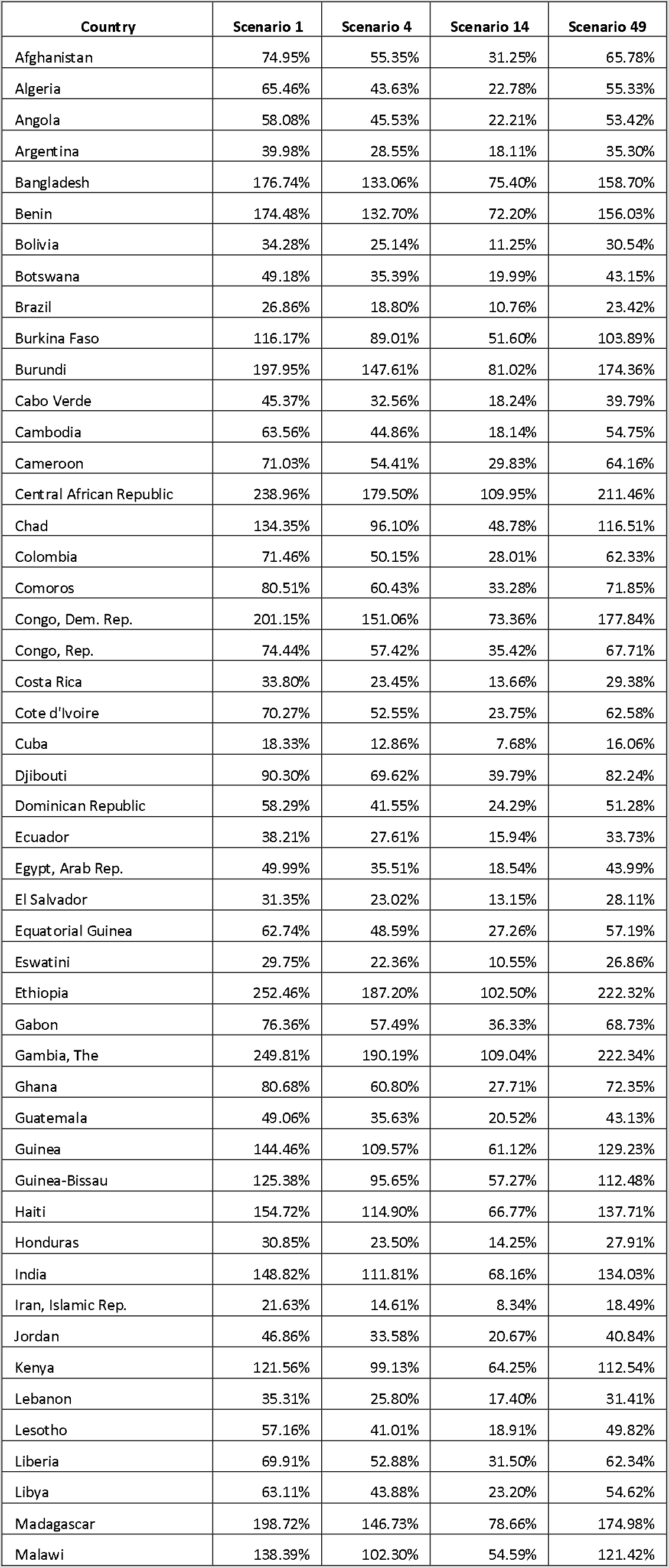
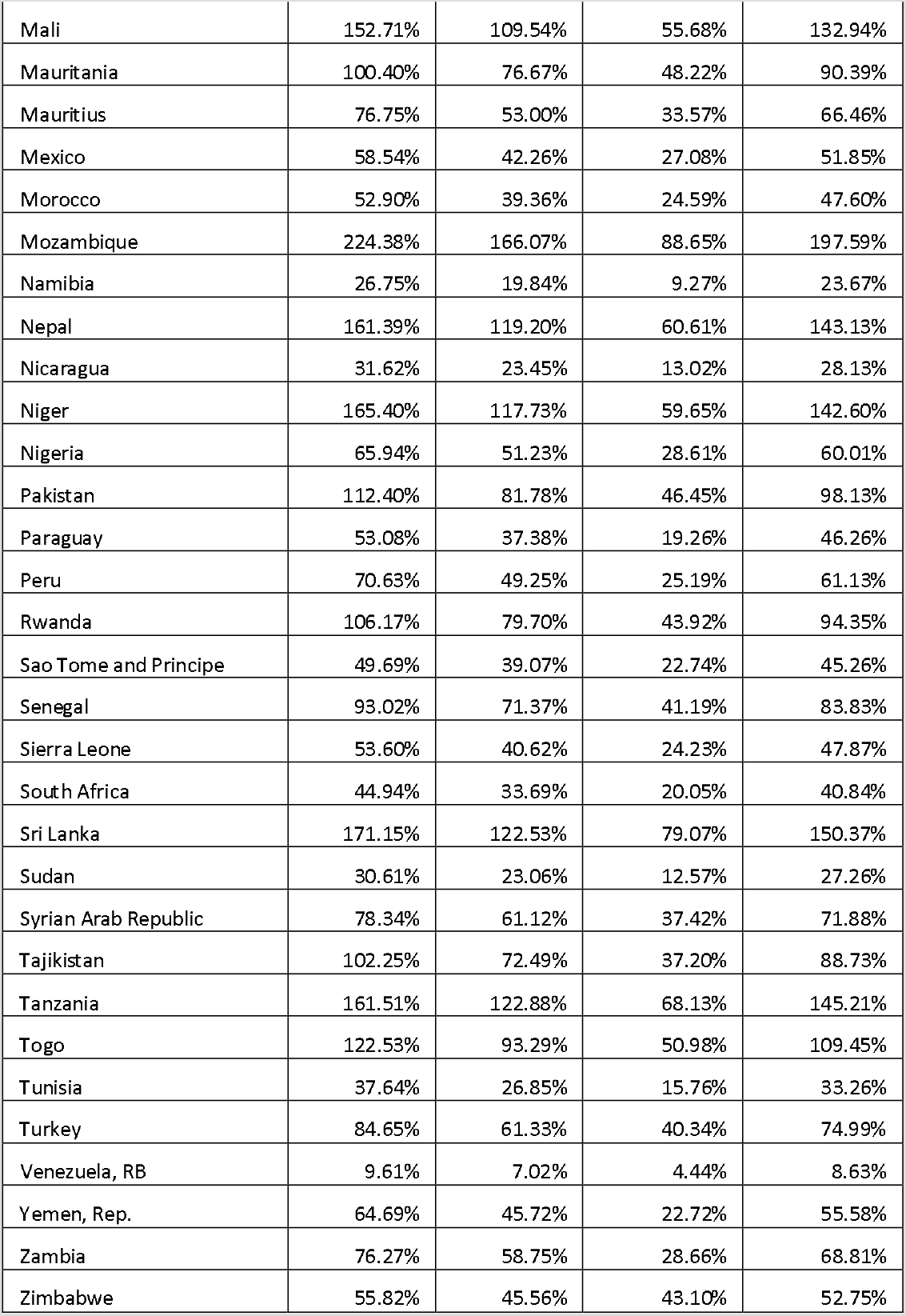
Health System Costs of COVID 19 Response as % of total health spending (excl. OOP)

**Table SR6:**
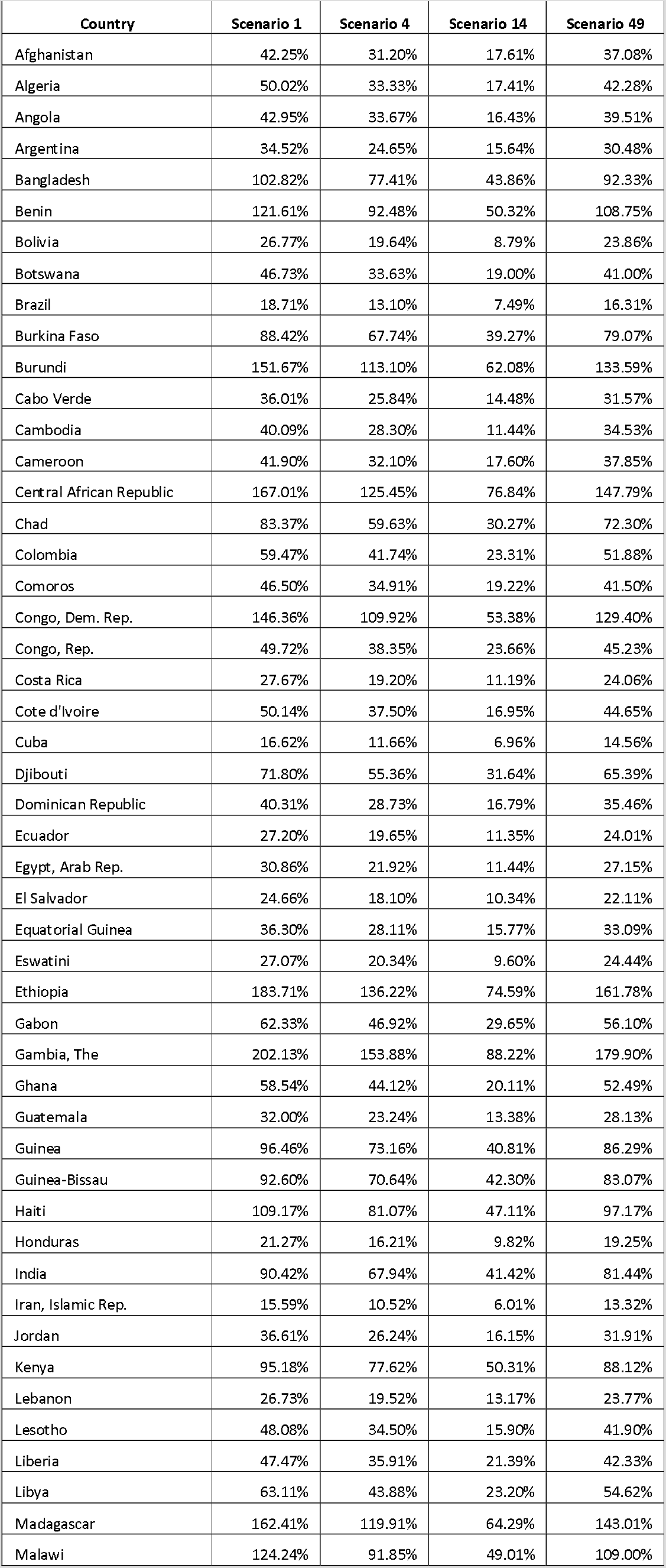
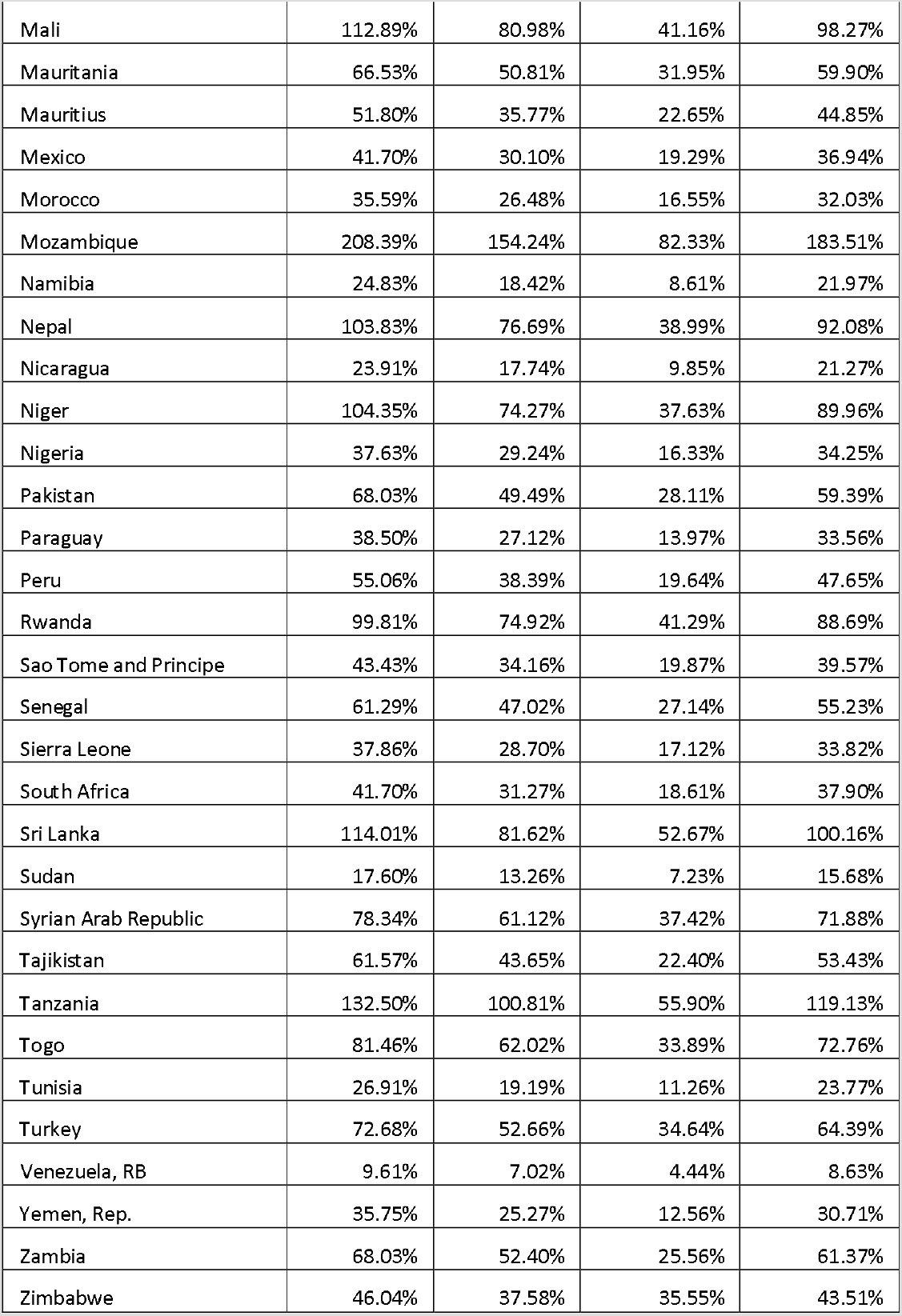
Health System Costs of COVID 19 Response as % of total health spending (incl. OOP)

**Table SR7:** Health System Costs of COVID 19 Response as % of government health spending

| Country | Scenario 1 | Scenario 4 | Scenario 14 | Scenario 49 |
| --- | --- | --- | --- | --- |
| Afghanistan | 1461.58% | 1079.37% | 609.33% | 1282.69% |
| Algeria | 96.72% | 81.75% | 33.66% | 81.75% |
| Angola | 131.58% | 121.02% | 50.32% | 121.02% |
| Argentina | 53.71% | 47.42% | 24.33% | 47.42% |
| Bangladesh | 984.27% | 883.78% | 419.89% | 883.78% |
| Benin | 849.59% | 759.74% | 351.58% | 759.74% |
| Bolivia | 52.17% | 46.49% | 17.12% | 46.49% |
| Botswana | 87.93% | 77.14% | 35.75% | 77.14% |
| Brazil | 80.84% | 70.48% | 32.38% | 70.48% |
| Burkina Faso | 289.72% | 259.10% | 128.69% | 259.10% |
| Burundi | 679.59% | 598.61% | 278.16% | 598.61% |
| Cabo Verde | 79.91% | 70.07% | 32.13% | 70.07% |
| Cambodia | 291.48% | 251.05% | 83.20% | 251.05% |
| Cameroon | 532.62% | 481.16% | 223.68% | 481.16% |
| Central African Republic | 1607.91% | 1422.83% | 739.81% | 1422.83% |
| Chad | 712.01% | 617.48% | 258.53% | 617.48% |
| Colombia | 112.68% | 98.29% | 44.17% | 98.29% |
| Comoros | 553.08% | 493.60% | 228.65% | 493.60% |
| Congo, Dem. Rep. | 1641.60% | 1451.34% | 598.70% | 1451.34% |
| Congo, Rep. | 176.09% | 160.17% | 83.78% | 160.17% |
| Costa Rica | 45.22% | 39.31% | 18.28% | 39.31% |
| Cote d'Ivoire | 272.68% | 242.80% | 92.15% | 242.80% |
| Cuba | 20.46% | 17.93% | 8.57% | 17.93% |
| Djibouti | 197.29% | 179.69% | 86.95% | 179.69% |
| Dominican Republic | 127.48% | 112.14% | 53.11% | 112.14% |
| Ecuador | 74.65% | 65.88% | 31.15% | 65.88% |
| Egypt, Arab Rep. | 170.63% | 150.12% | 63.26% | 150.12% |
| El Salvador | 48.63% | 43.61% | 20.40% | 43.61% |
| Equatorial Guinea | 266.82% | 243.24% | 115.92% | 243.24% |
| Eswatini | 42.93% | 38.75% | 15.22% | 38.75% |
| Ethiopia | 914.16% | 805.02% | 371.17% | 805.02% |
| Gabon | 118.23% | 106.41% | 56.24% | 106.41% |
| Gambia, The | 1345.47% | 1197.51% | 587.26% | 1197.51% |
| Ghana | 210.35% | 188.62% | 72.25% | 188.62% |
| Guatemala | 131.83% | 115.89% | 55.14% | 115.89% |
| Guinea | 1174.62% | 1050.76% | 496.94% | 1050.76% |
| Guinea-Bissau | 283.34% | 254.19% | 129.43% | 254.19% |
| Haiti | 1008.94% | 898.02% | 435.37% | 898.02% |
| Honduras | 67.20% | 60.80% | 31.03% | 60.80% |
| India | 585.28% | 527.12% | 268.06% | 527.12% |
| Iran, Islamic Rep. | 39.68% | 33.91% | 15.30% | 33.91% |
| Jordan | 74.11% | 64.60% | 32.69% | 64.60% |
| Kenya | 336.03% | 311.08% | 177.61% | 311.08% |
| Lebanon | 67.74% | 60.24% | 33.38% | 60.24% |
| Lesotho | 89.58% | 78.08% | 29.63% | 78.08% |
| Liberia | 490.72% | 437.58% | 221.12% | 437.58% |
| Libya | 99.72% | 86.30% | 36.66% | 86.30% |
| Madagascar | 417.01% | 367.20% | 165.06% | 367.20% |
| Malawi | 493.40% | 432.89% | 194.62% | 432.89% |
| Mali | 483.29% | 420.70% | 176.21% | 420.70% |
| Mauritania | 273.93% | 246.63% | 131.56% | 246.63% |
| Mauritius | 173.99% | 150.65% | 76.09% | 150.65% |
| Mexico | 112.26% | 99.44% | 51.94% | 99.44% |
| Morocco | 112.89% | 101.59% | 52.48% | 101.59% |
| Mozambique | 420.65% | 370.42% | 166.19% | 370.42% |
| Namibia | 43.22% | 38.24% | 14.98% | 38.24% |
| Nepal | 868.62% | 770.34% | 326.22% | 770.34% |
| Nicaragua | 51.54% | 45.85% | 21.22% | 45.85% |
| Niger | 680.99% | 587.10% | 245.58% | 587.10% |
| Nigeria | 506.26% | 460.77% | 219.63% | 460.77% |
| Pakistan | 402.92% | 351.76% | 166.50% | 351.76% |
| Paraguay | 102.82% | 89.61% | 37.32% | 89.61% |
| Peru | 110.23% | 95.40% | 39.32% | 95.40% |
| Rwanda | 313.36% | 278.46% | 129.63% | 278.46% |
| Sao Tome and Principe | 124.55% | 113.46% | 56.99% | 113.46% |
| Senegal | 269.12% | 242.51% | 119.16% | 242.51% |
| Sierra Leone | 479.73% | 428.42% | 216.88% | 428.42% |
| South Africa | 83.63% | 76.01% | 37.32% | 76.01% |
| Sri Lanka | 397.17% | 348.94% | 183.49% | 348.94% |
| Sudan | 157.07% | 139.90% | 64.52% | 139.90% |
| Syrian Arab Republic | 173.02% | 158.74% | 82.63% | 158.74% |
| Tajikistan | 357.52% | 310.26% | 130.06% | 310.26% |
| Tanzania | 397.58% | 357.44% | 167.72% | 357.44% |
| Togo | 611.16% | 545.92% | 254.28% | 545.92% |
| Tunisia | 66.46% | 58.72% | 27.83% | 58.72% |
| Turkey | 107.91% | 95.60% | 51.43% | 95.60% |
| Venezuela, RB | 25.19% | 22.64% | 11.65% | 22.64% |
| Yemen, Rep. | 635.34% | 545.82% | 223.14% | 545.82% |
| Zambia | 199.20% | 179.70% | 74.84% | 179.70% |
| Zimbabwe | 120.03% | 113.43% | 92.69% | 113.43% |

**Table SR8.**
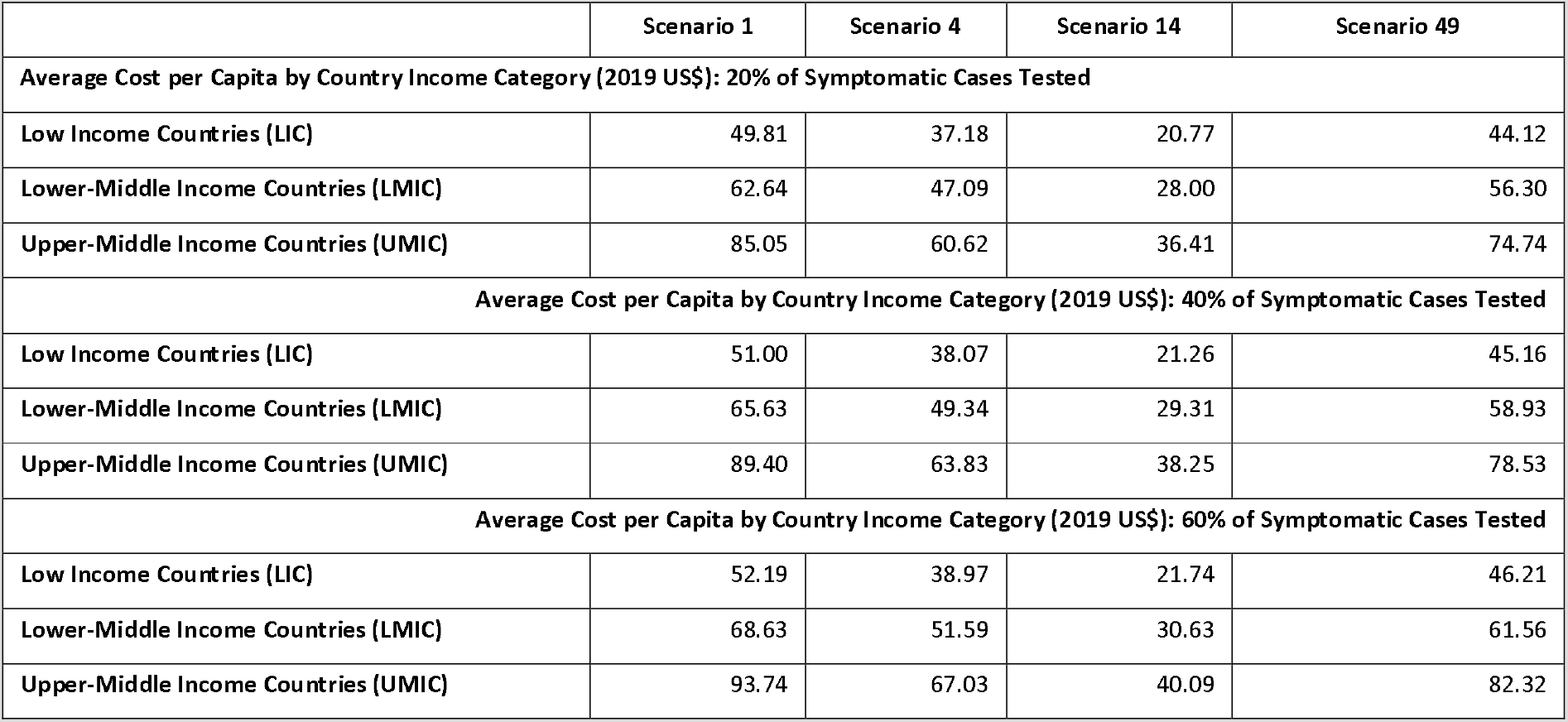
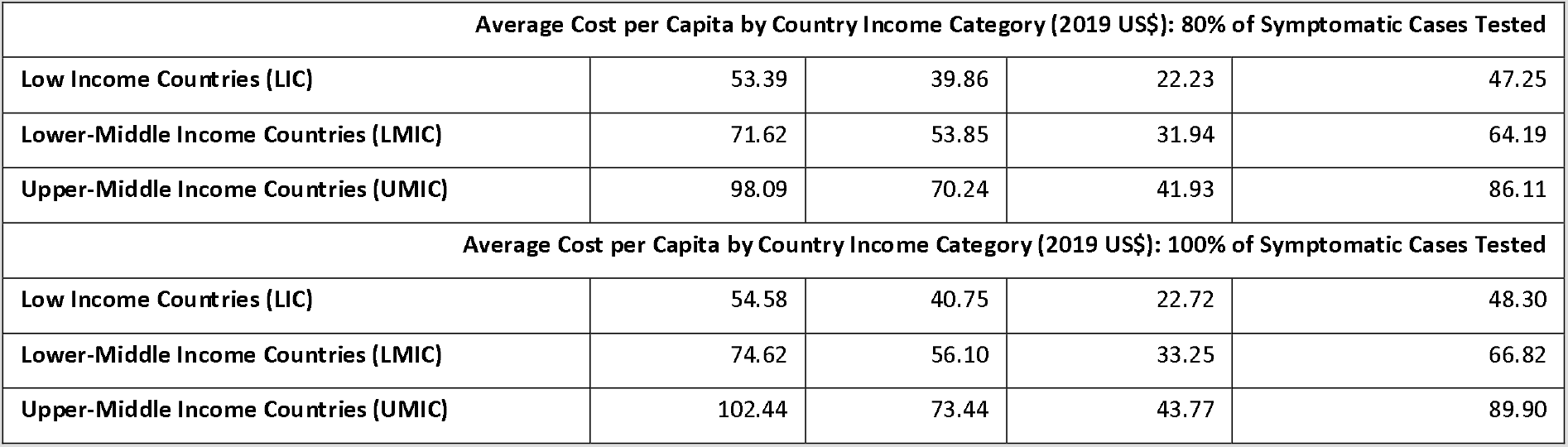
Average Cost per Capita by Country Income Category (2019 US$): Sensitivity Analysis on % of symptomatic cases tested

|  | Scenario 1 | Scenario 4 | Scenario 14 | Scenario 49 |
| --- | --- | --- | --- | --- |
| **Average Cost per Capita by Country Income Category (2019 US\$): 20% of Symptomatic Cases Tested** | | | | |
| Low Income Countries (LIC) | 49.81 | 37.18 | 20.77 | 44.12 |
| Lower-Middle Income Countries (LMIC) | 62.64 | 47.09 | 28.00 | 56.30 |
| Upper-Middle Income Countries (UMIC) | 85.05 | 60.62 | 36.41 | 74.74 |
| **Average Cost per Capita by Country Income Category (2019 US\$): 40% of Symptomatic Cases Tested** | | | | |
| Low Income Countries (LIC) | 51.00 | 38.07 | 21.26 | 45.16 |
| Lower-Middle Income Countries (LMIC) | 65.63 | 49.34 | 29.31 | 58.93 |
| Upper-Middle Income Countries (UMIC) | 89.40 | 63.83 | 38.25 | 78.53 |
| **Average Cost per Capita by Country Income Category (2019 US\$): 60% of Symptomatic Cases Tested** | | | | |
| Low Income Countries (LIC) | 52.19 | 38.97 | 21.74 | 46.21 |
| Lower-Middle Income Countries (LMIC) | 68.63 | 51.59 | 30.63 | 61.56 |
| Upper-Middle Income Countries (UMIC) | 93.74 | 67.03 | 40.09 | 82.32 |

| Average Cost per Capita by Country Income Category (2019 US\$): 80% of Symptomatic Cases Tested | | | | |
| --- | --- | --- | --- | --- |
| Low Income Countries (LIC) | 53.39 | 39.86 | 22.23 | 47.25 |
| Lower-Middle Income Countries (LMIC) | 71.62 | 53.85 | 31.94 | 64.19 |
| Upper-Middle Income Countries (UMIC) | 98.09 | 70.24 | 41.93 | 86.11 |
| Average Cost per Capita by Country Income Category (2019 US\$): 100% of Symptomatic Cases Tested | | | | |
| Low Income Countries (LIC) | 54.58 | 40.75 | 22.72 | 48.30 |
| Lower-Middle Income Countries (LMIC) | 74.62 | 56.10 | 33.25 | 66.82 |
| Upper-Middle Income Countries (UMIC) | 102.44 | 73.44 | 43.77 | 89.90 |

## Supplemental Material Methods

### Supplement S1. Epidemiological model

#### S1.1 Parameters used in epidemiological model

**Figure S1.1.1.**
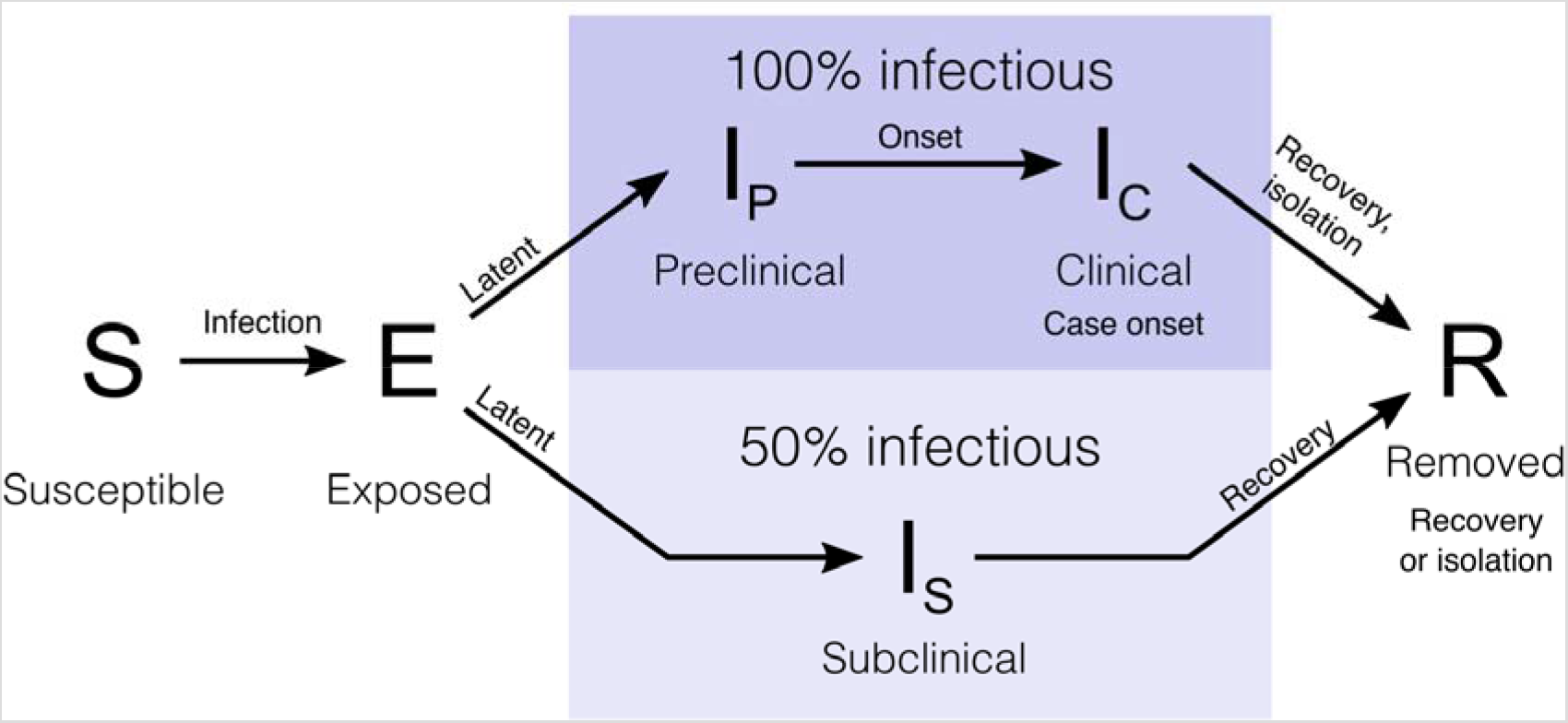
Flow diagram showing compartments and flows in the epidemiological model.

**Table S1.1.1.**
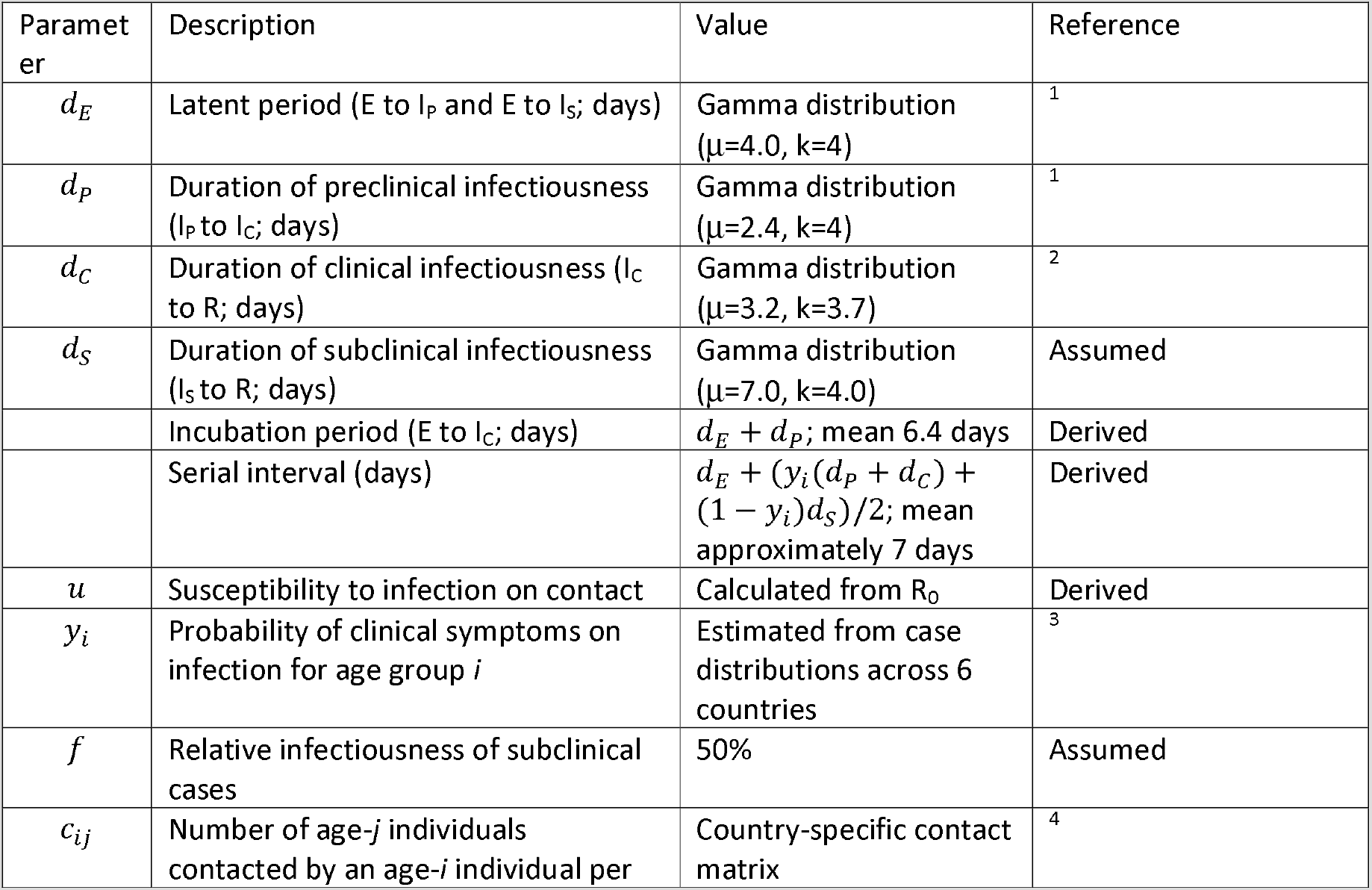
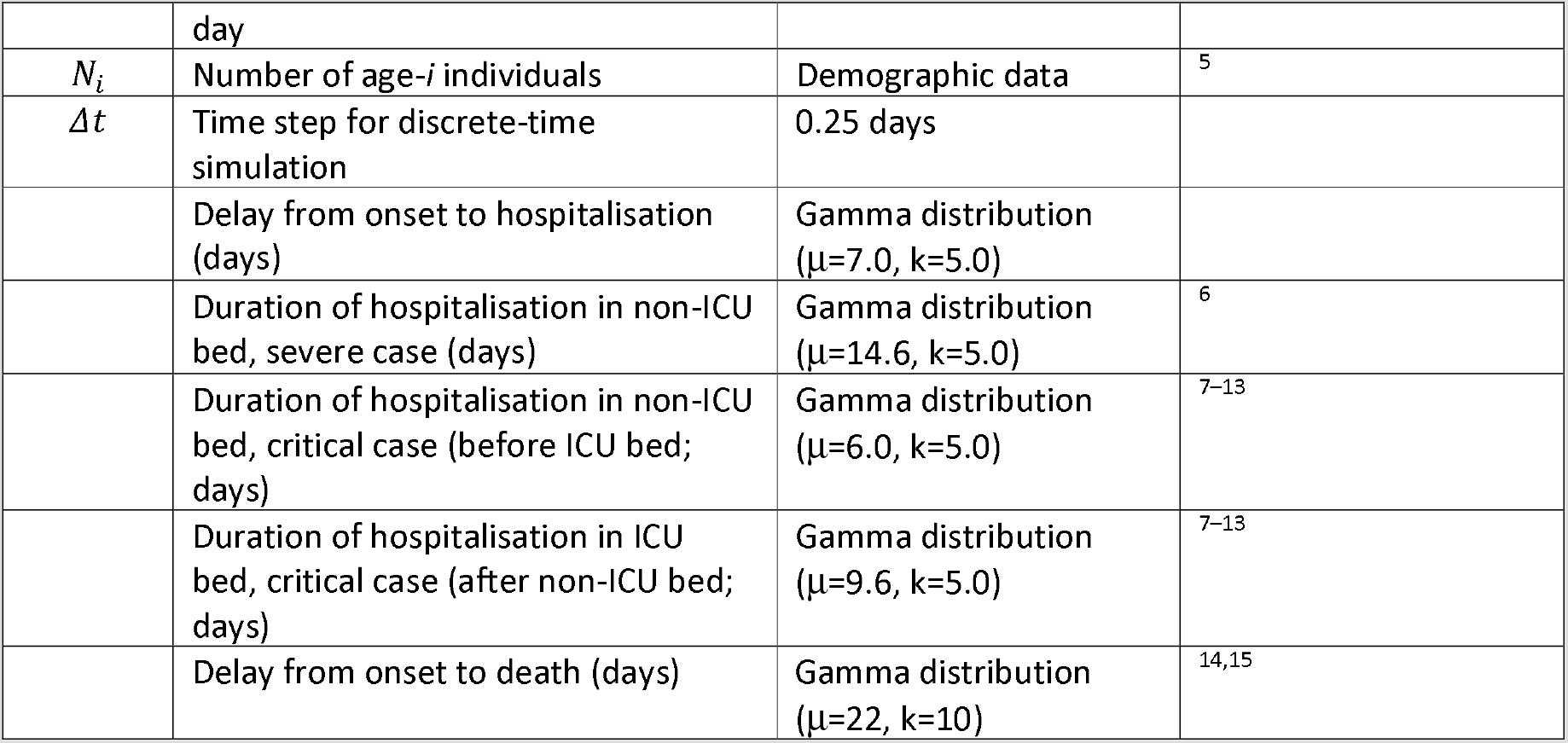
General model parameters

| Parameter | Description | Value | Reference |
| --- | --- | --- | --- |
| $d_E$ | Latent period (E to $I_P$ and E to $I_S$; days) | Gamma distribution ($\mu=4.0$, $k=4$) | <sup>1</sup> |
| $d_P$ | Duration of preclinical infectiousness ($I_P$ to $I_C$; days) | Gamma distribution ($\mu=2.4$, $k=4$) | <sup>1</sup> |
| $d_C$ | Duration of clinical infectiousness ($I_C$ to R; days) | Gamma distribution ($\mu=3.2$, $k=3.7$) | <sup>2</sup> |
| $d_S$ | Duration of subclinical infectiousness ($I_S$ to R; days) | Gamma distribution ($\mu=7.0$, $k=4.0$) | Assumed |
| | Incubation period (E to $I_C$; days) | $d_E + d_P$; mean 6.4 days | Derived |
| | Serial interval (days) | $d_E + (y_i(d_P + d_C) + (1 - y_i)d_S)/2$; mean approximately 7 days | Derived |
| $u$ | Susceptibility to infection on contact | Calculated from $R_0$ | Derived |
| $y_i$ | Probability of clinical symptoms on infection for age group $i$ | Estimated from case distributions across 6 countries | <sup>3</sup> |
| $f$ | Relative infectiousness of subclinical cases | 50% | Assumed |
| $c_{ij}$ | Number of age- $j$ individuals contacted by an age- $i$ individual per day | Country-specific contact matrix | <sup>4</sup> |
| $N_i$ | Number of age-$i$ individuals | Demographic data | <sup>5</sup> |
| $\Delta t$ | Time step for discrete-time simulation | 0.25 days | |
| | Delay from onset to hospitalisation (days) | Gamma distribution ($\mu=7.0$, $k=5.0$) | |
| | Duration of hospitalisation in non-ICU bed, severe case (days) | Gamma distribution ($\mu=14.6$, $k=5.0$) | <sup>6</sup> |
| | Duration of hospitalisation in non-ICU bed, critical case (before ICU bed; days) | Gamma distribution ($\mu=6.0$, $k=5.0$) | <sup>7-13</sup> |
| | Duration of hospitalisation in ICU bed, critical case (after non-ICU bed; days) | Gamma distribution ($\mu=9.6$, $k=5.0$) | <sup>7-13</sup> |
| | Delay from onset to death (days) | Gamma distribution ($\mu=22$, $k=10$) | <sup>14,15</sup> |

**Table S1.1.2.** Age-specific hospitalisation and fatality risk. From^16^

| Age group | Case-fatality risk | % of cases hospitalised | % of hospital patients needing ICU |
| --- | --- | --- | --- |
| 0–9 | 0.00% | 0.0% | 30% |
| 1–10 | 0.09% | 0.8% | 30% |
| 20–29 | 0.10% | 0.8% | 30% |
| 30–39 | 0.12% | 1.0% | 30% |
| 40–49 | 0.23% | 1.9% | 30% |
| 50–59 | 0.68% | 5.4% | 30% |
| 60–69 | 1.87% | 15.1% | 30% |
| 70–79 | 4.14% | 33.3% | 30% |
| 80+ | 7.68% | 61.8% | 30% |

#### S1.2 Scenarios

The epidemiological model uses data from 92 low- and middle-income countries. For each country, the model produces estimates on the number of cases, hospitalisations, number of days in hospital for severe cases (general ward) and critical cases (intensive care unit), and deaths for 57 distinct epidemiological scenarios [16].

For this study, four epidemiological scenarios were chosen out of the set of 57 possible scenarios. Scenario 1 represents an unmitigated epidemic. The other three scenarios were chosen because they represent a variety of plausible policy options (scenarios 4,14 and 49). Descriptions of the scenarios are presented below.

**Table S1.2.1.**
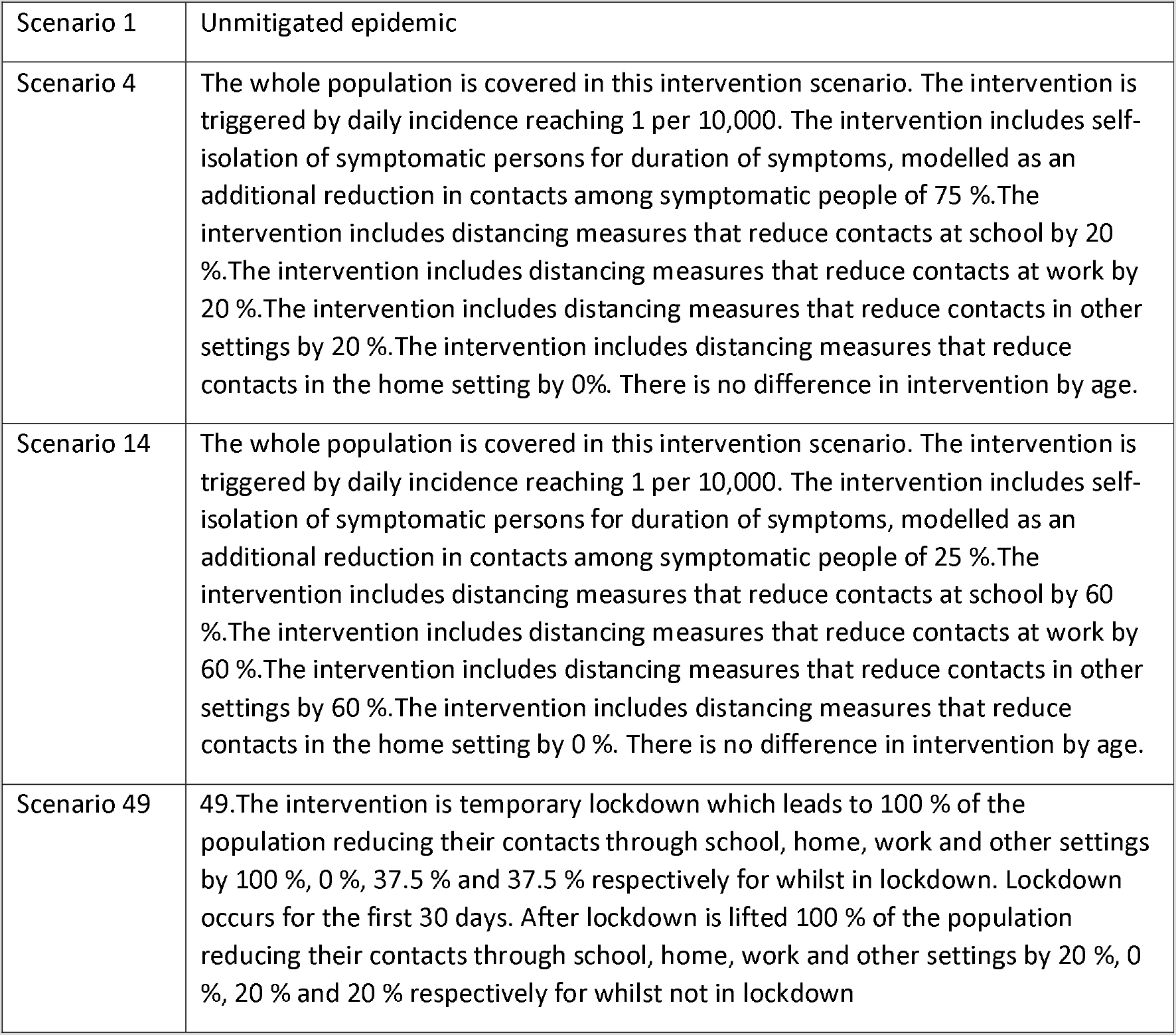
Scenario Descriptions

#### S1.3 The number of cases

The total number of expected cases, deaths, and days of hospitalisation in ICU and general ward per country per scenario used in this study are in the table below **[16]**.

**Table S1.3.1.**
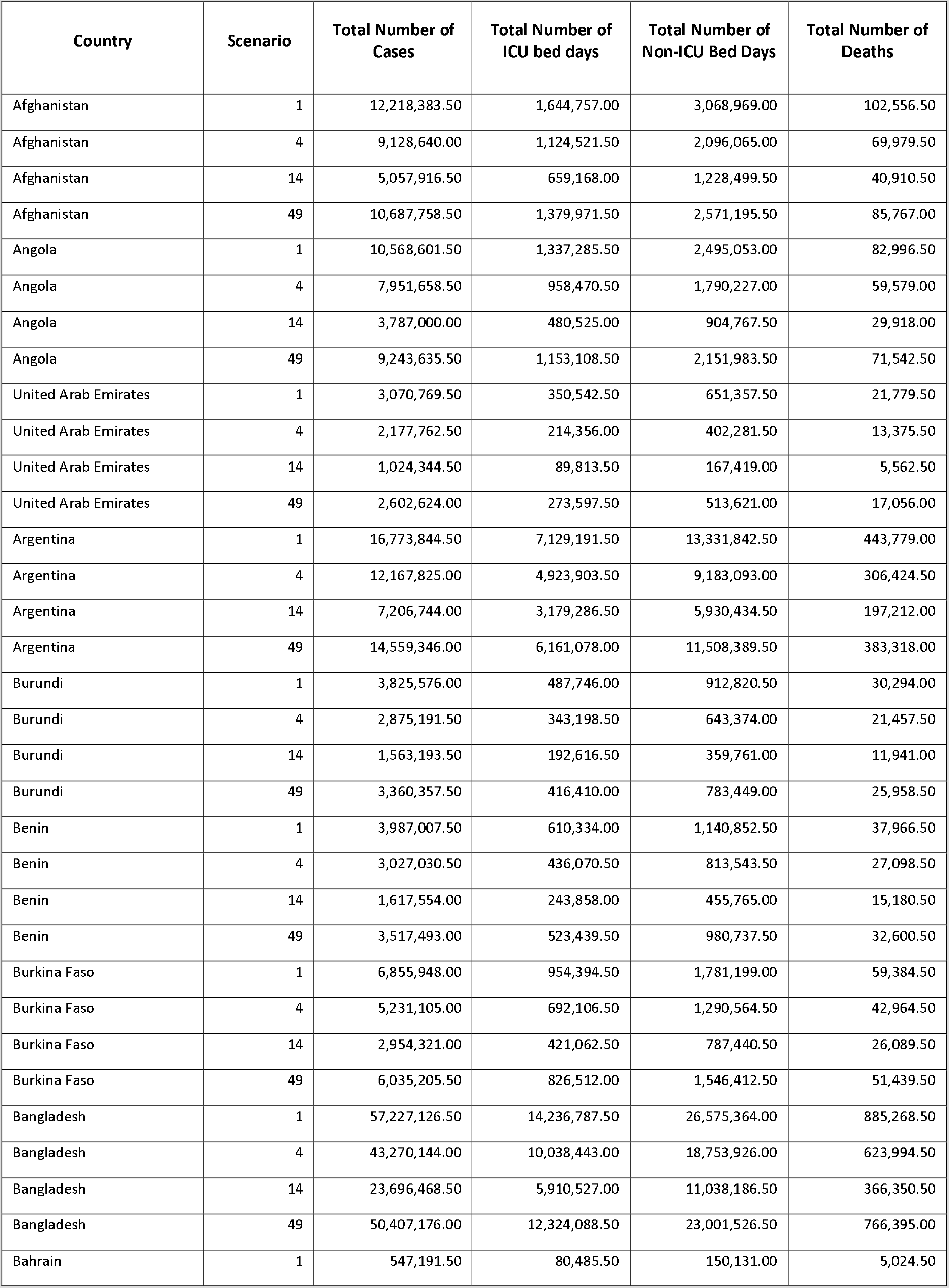
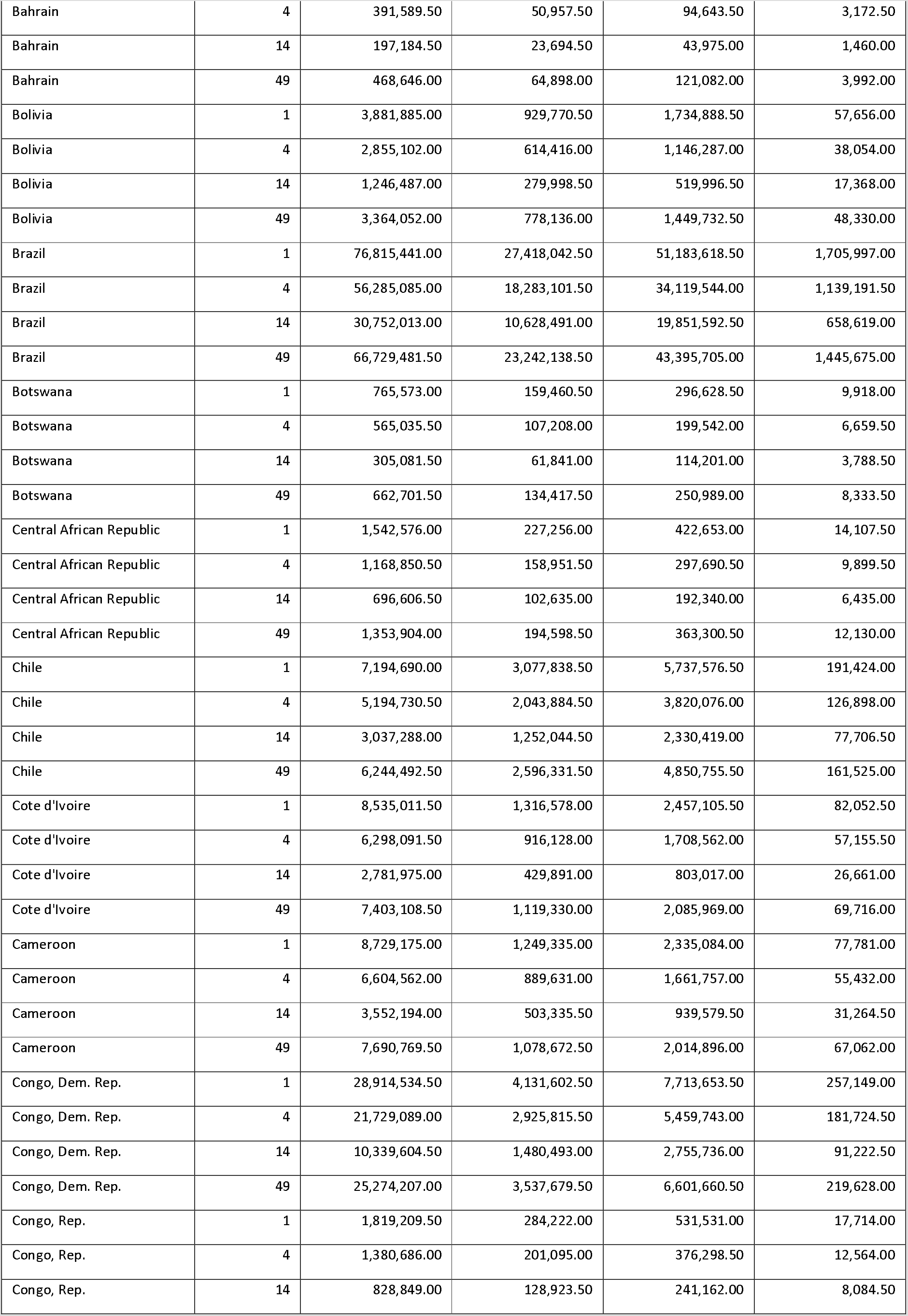
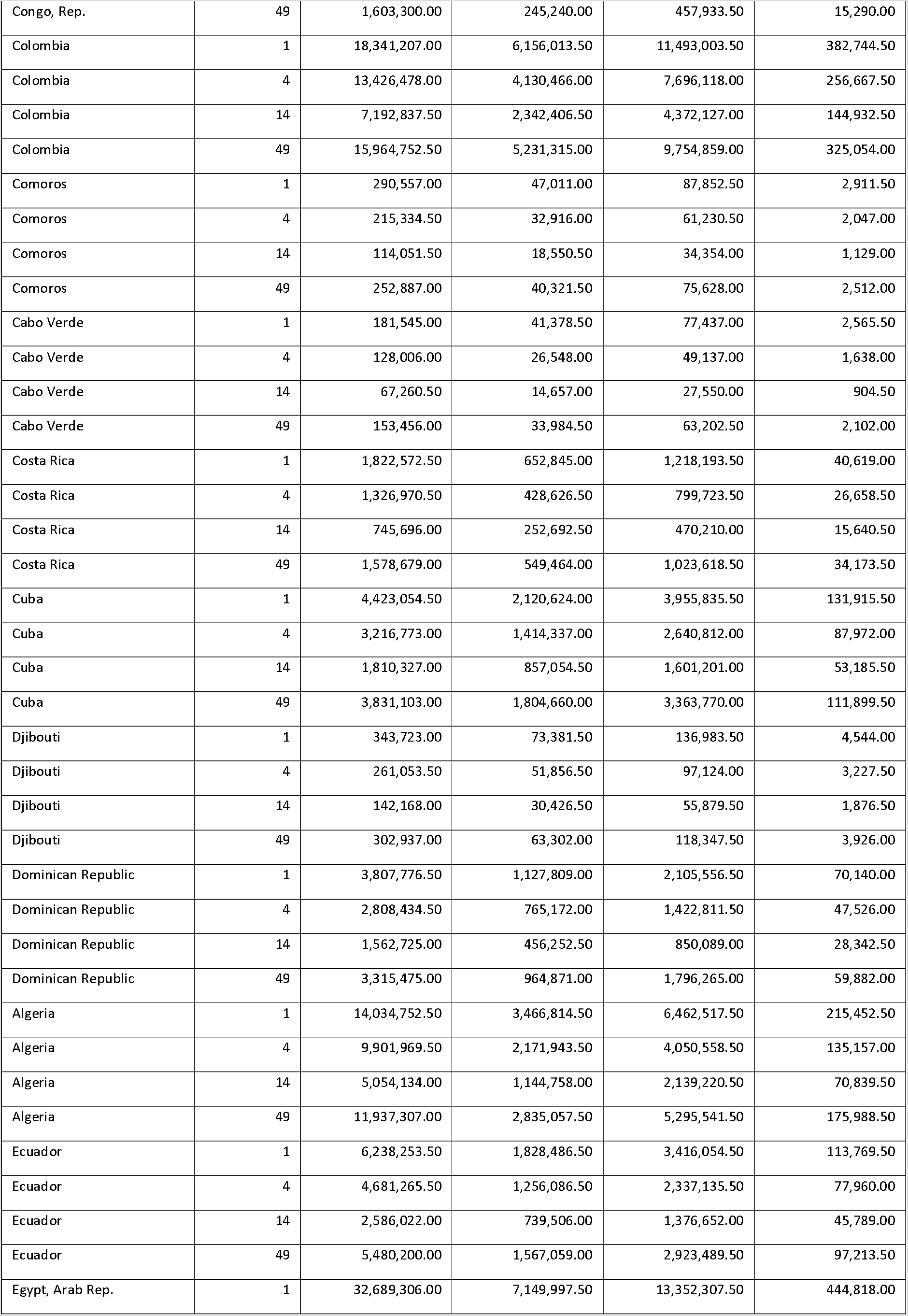
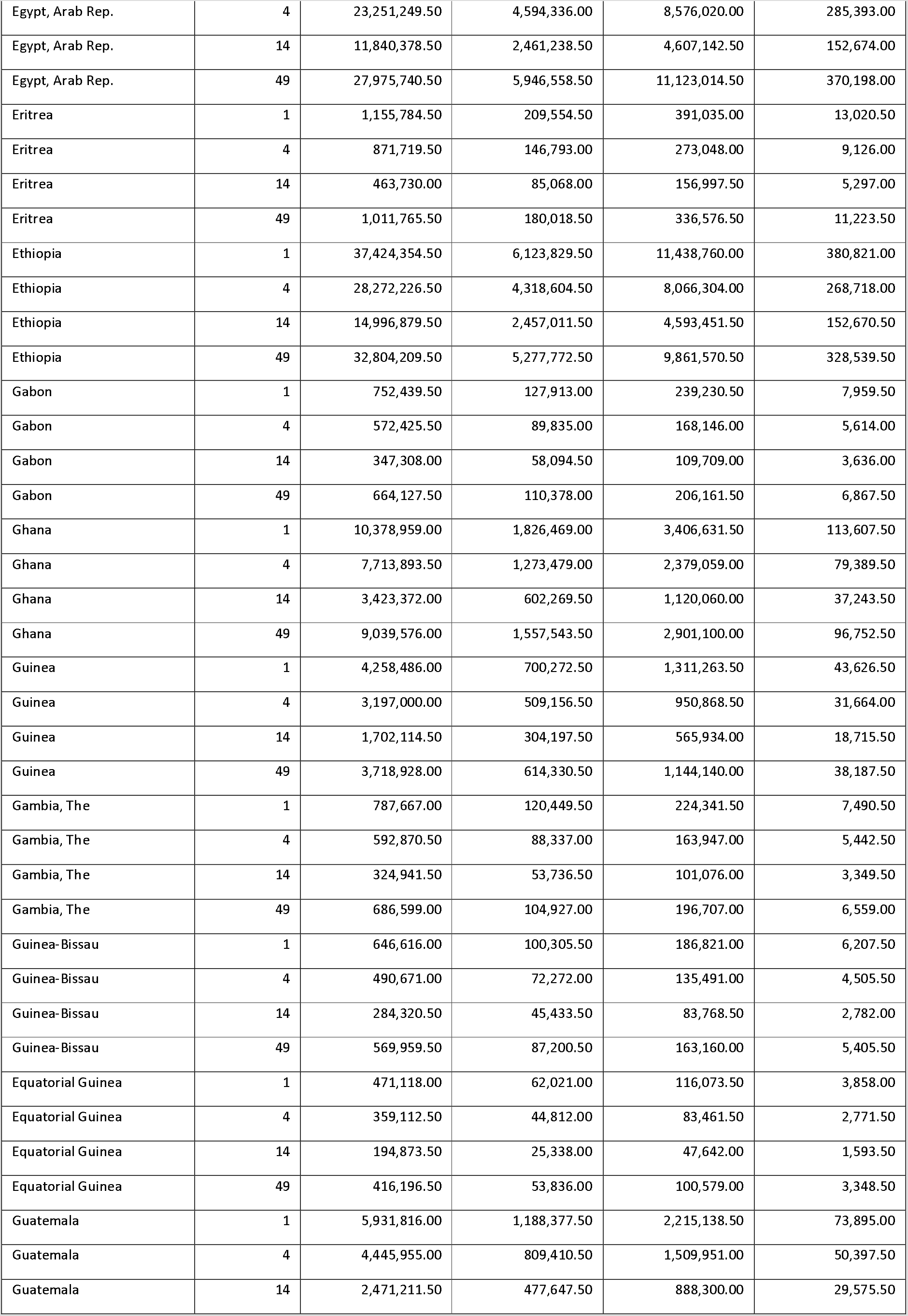
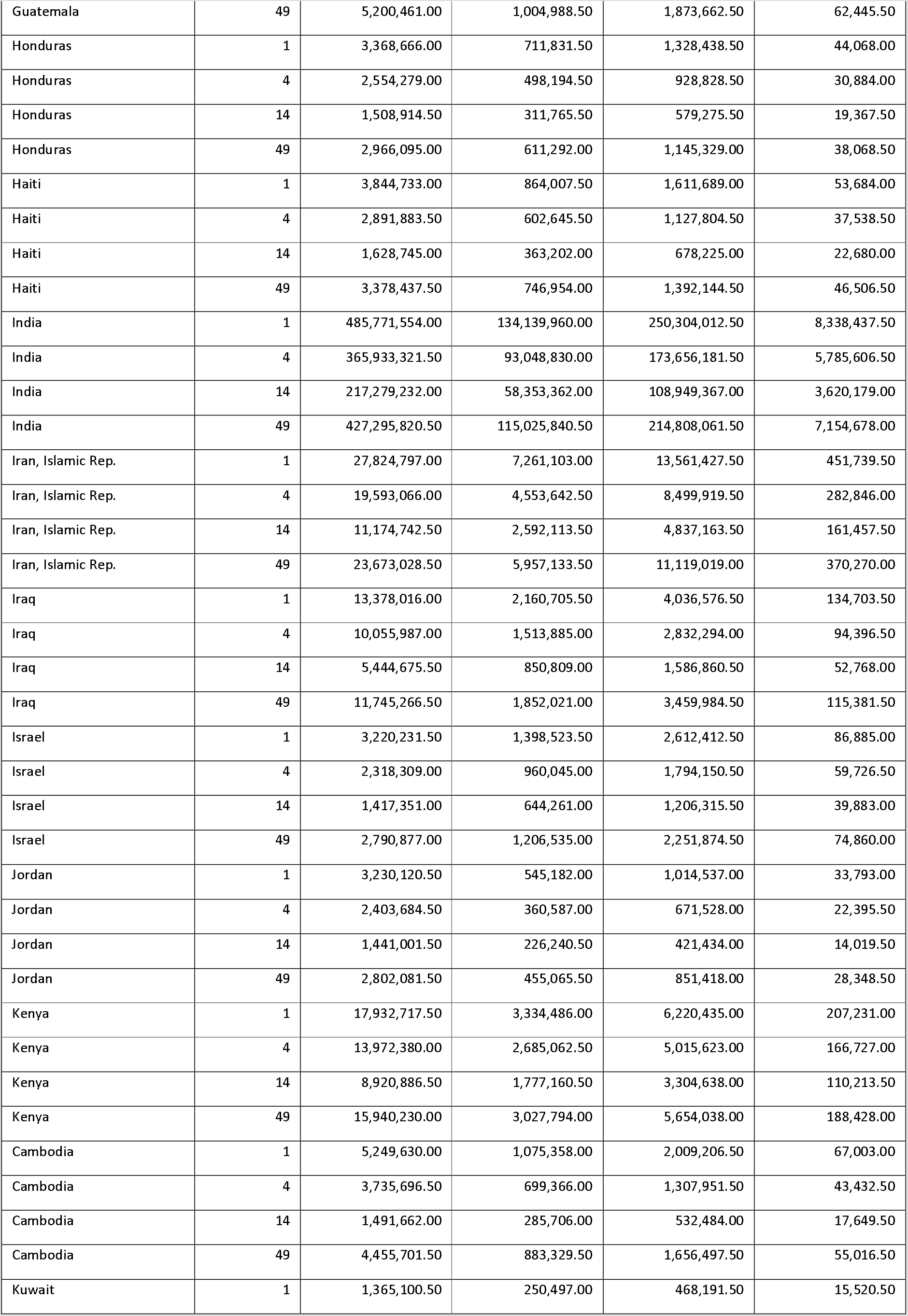
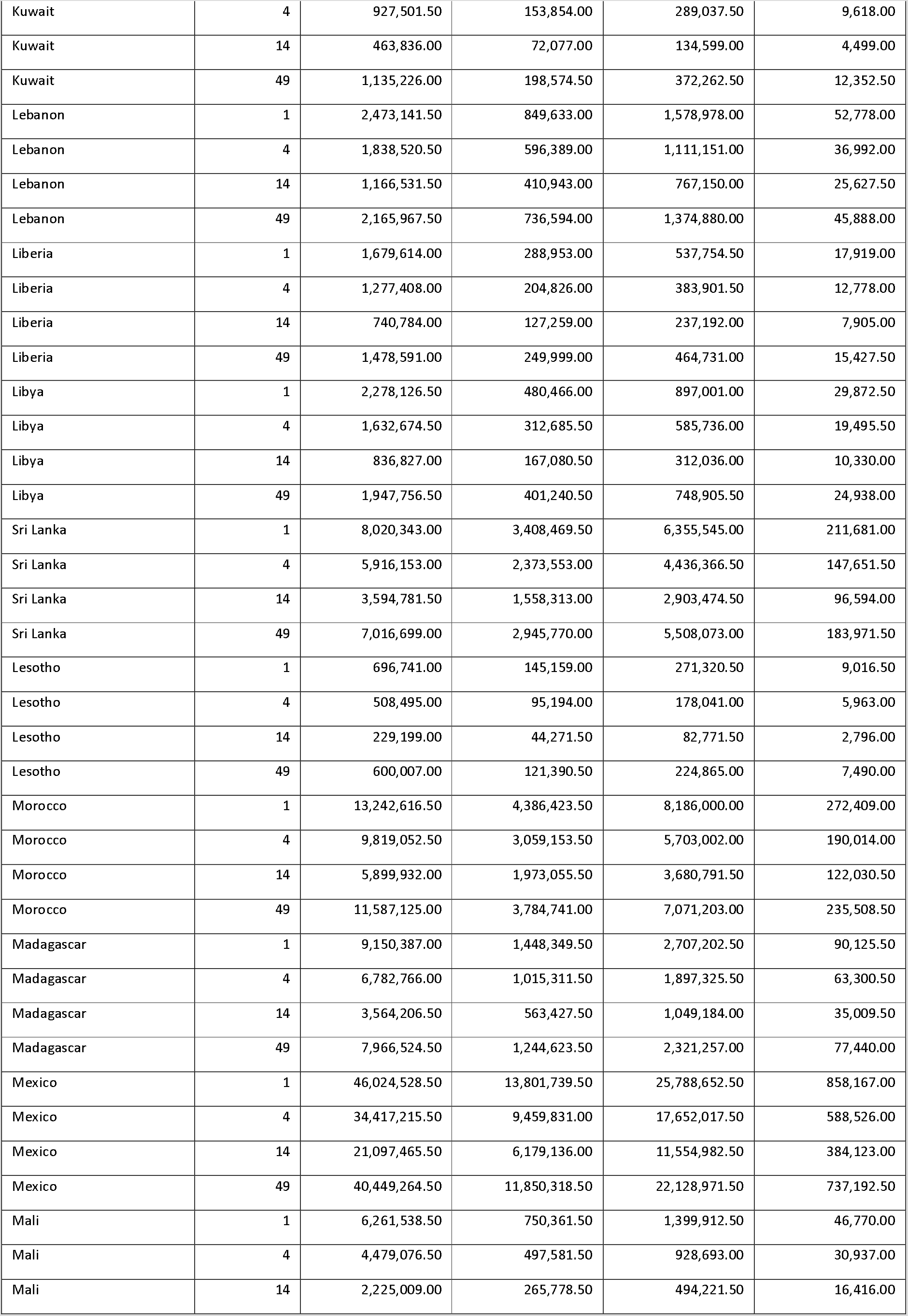
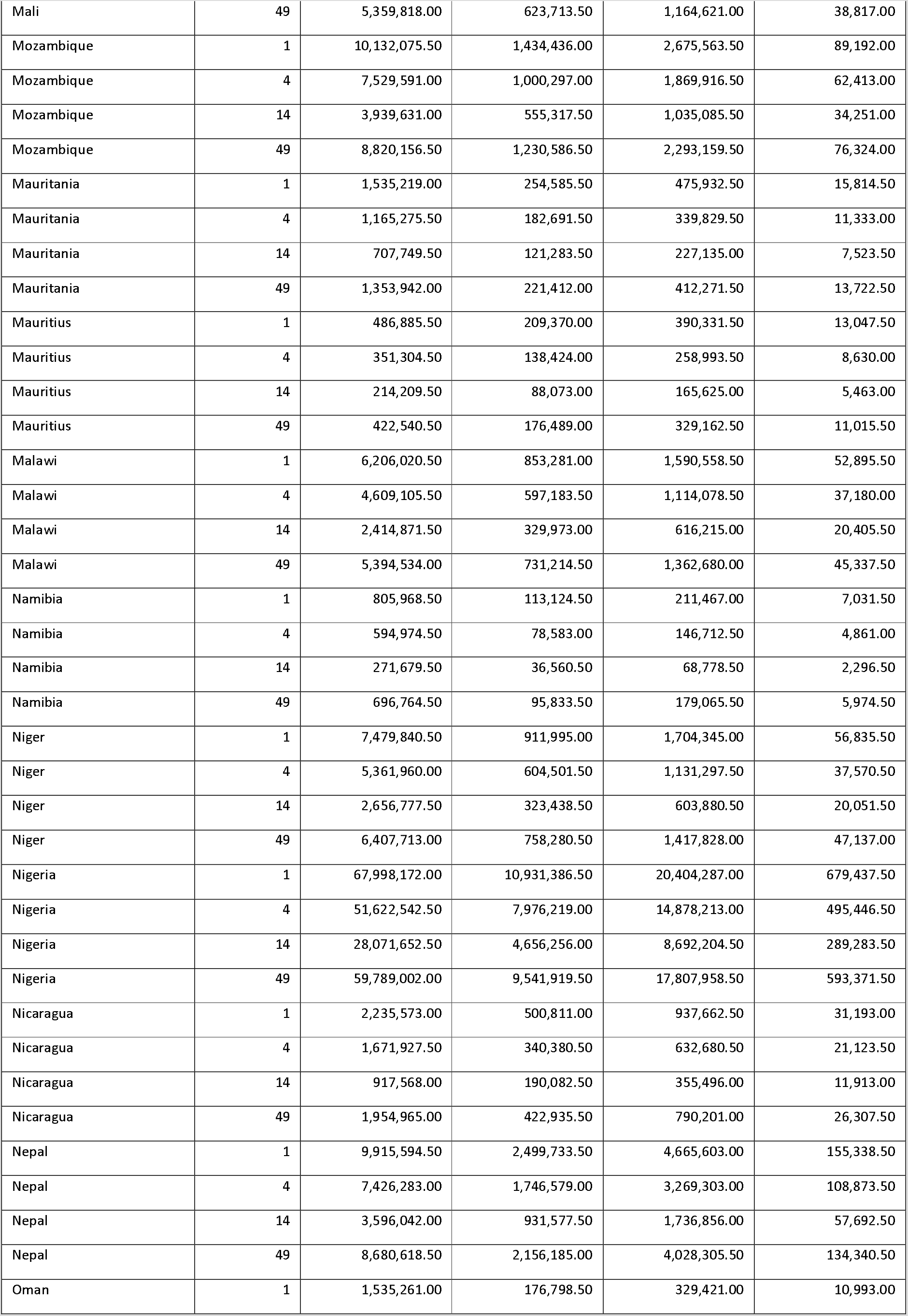
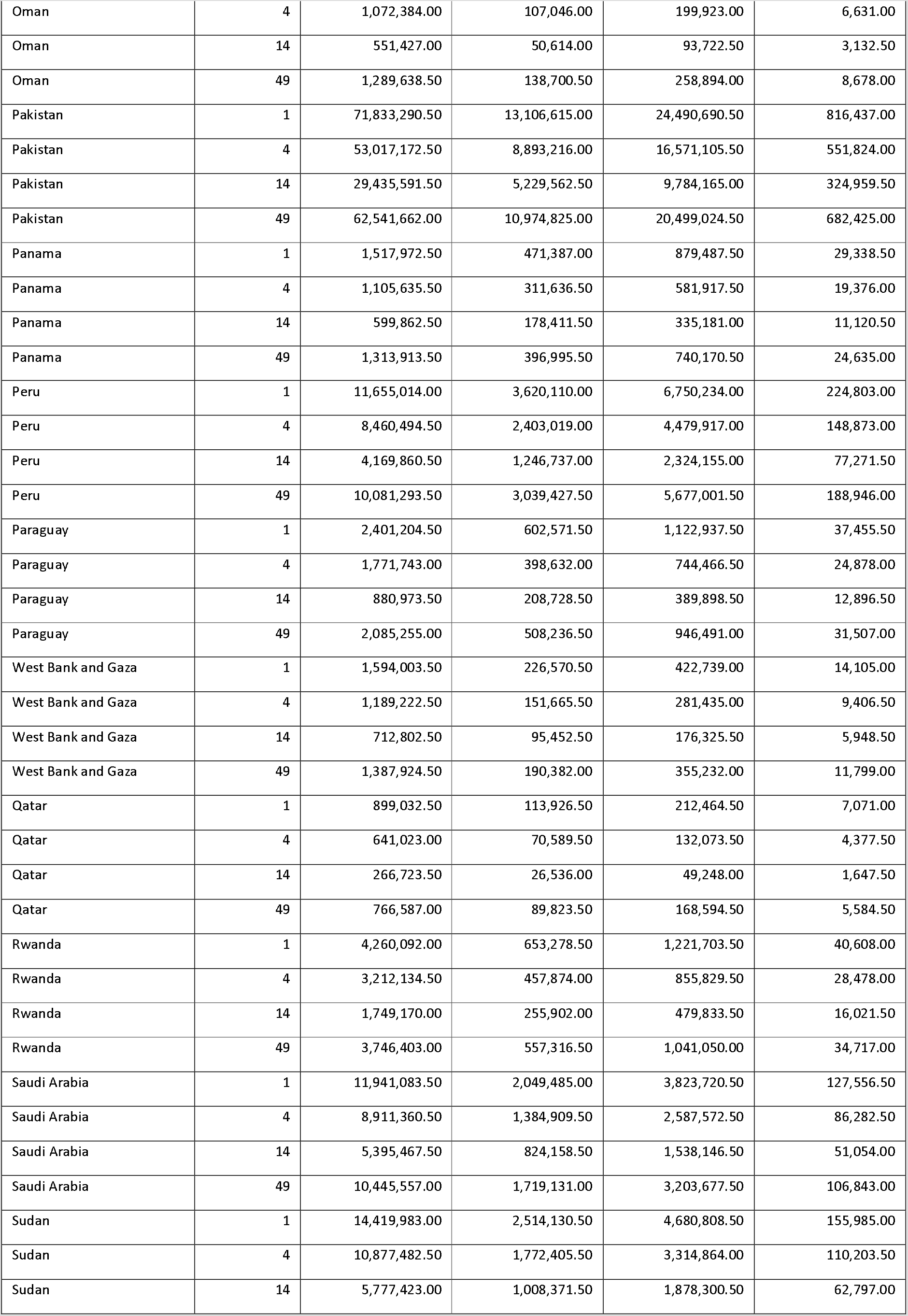
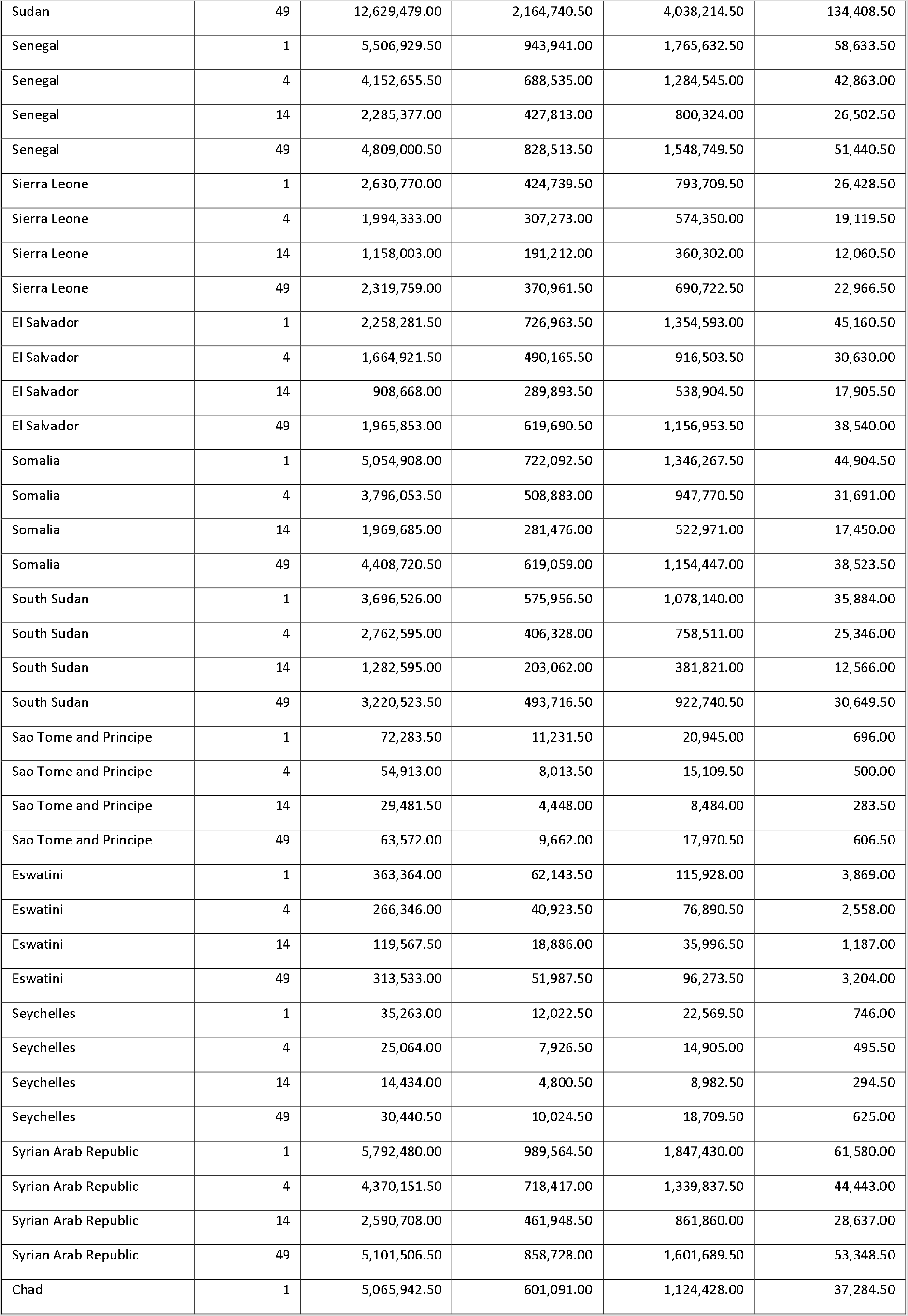
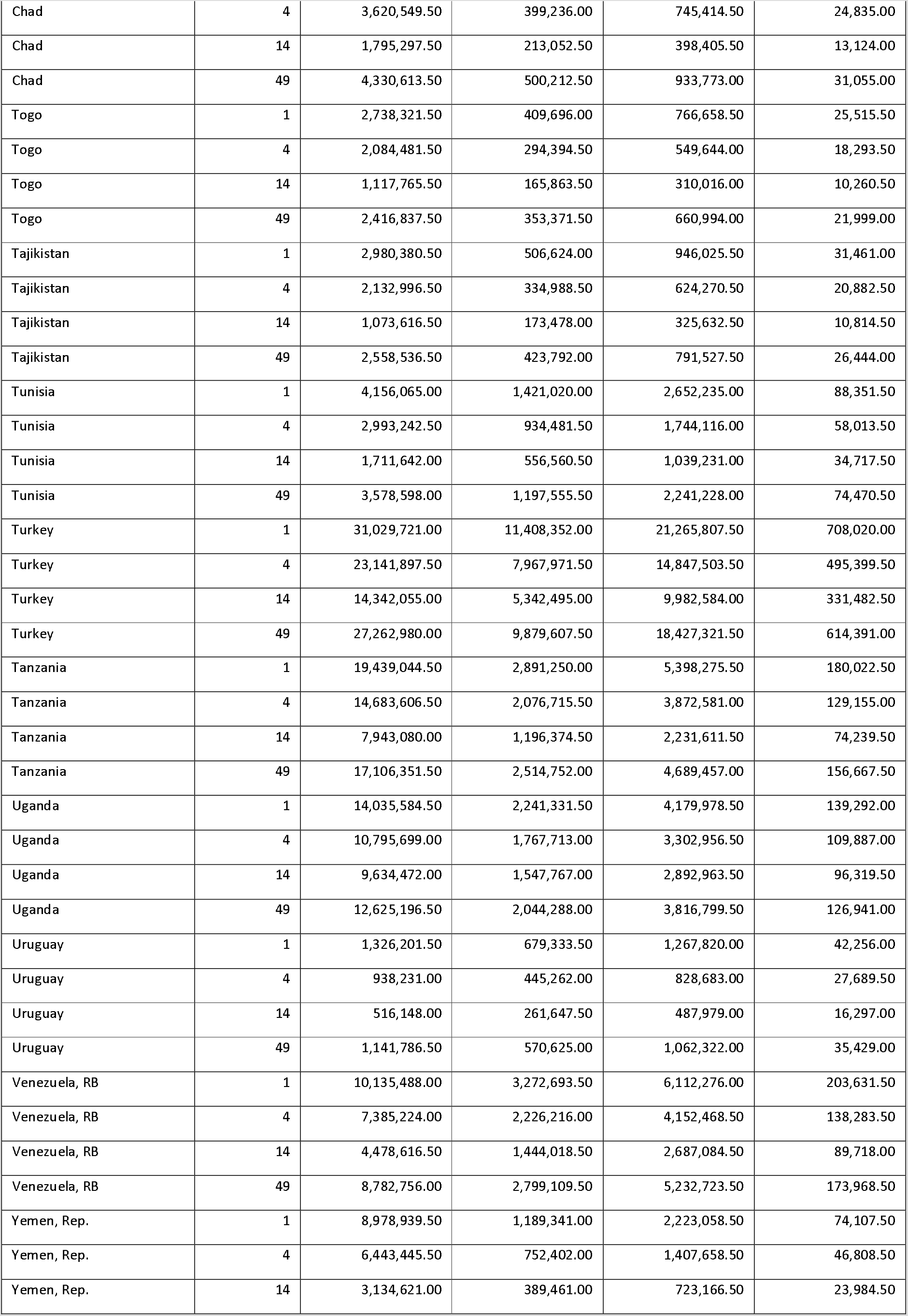
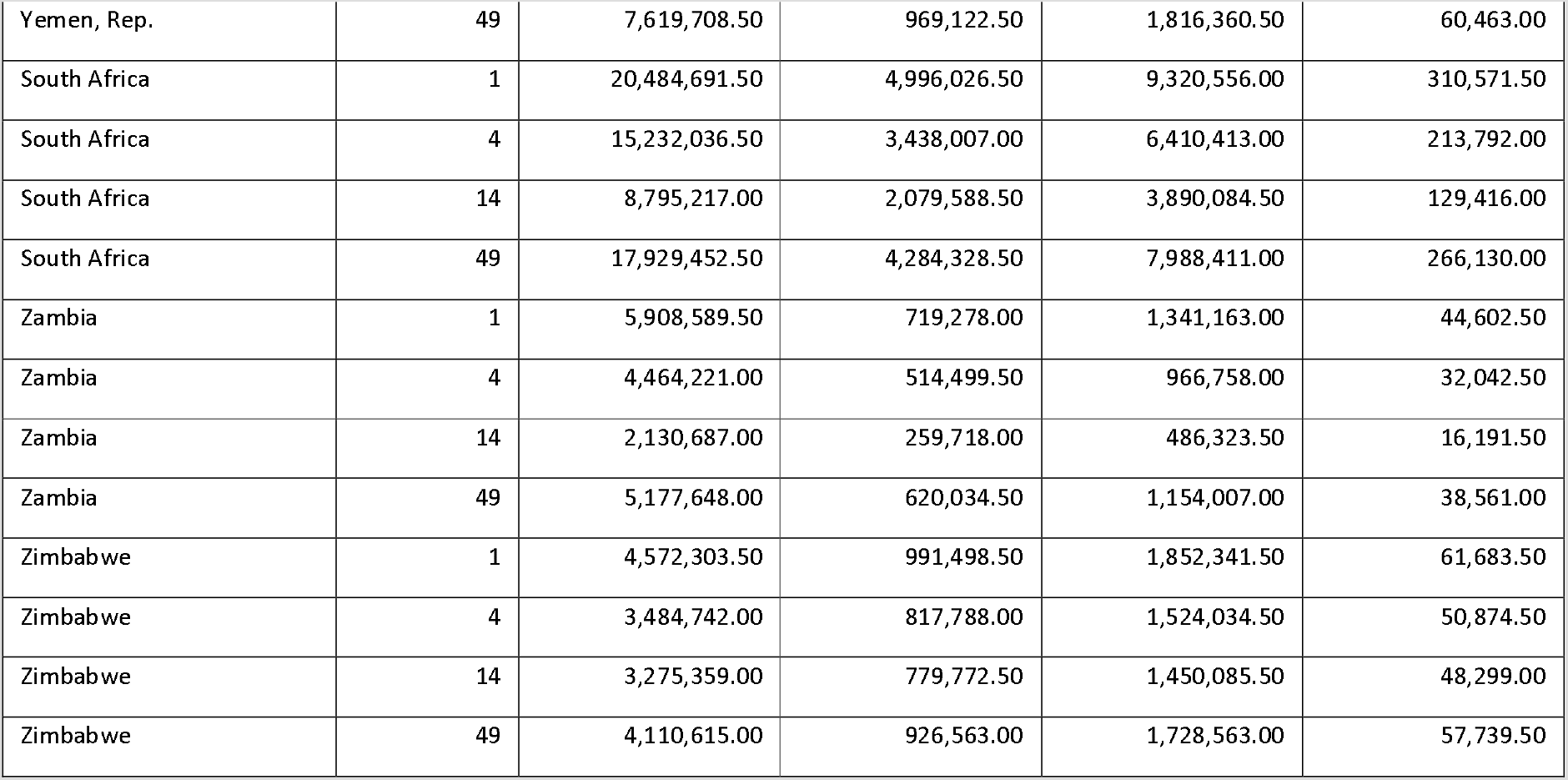
Number of cases, deaths, and days of hospitalisation per country per scenario

| Country | Scenario | Total Number of Cases | Total Number of ICU bed days | Total Number of Non-ICU Bed Days | Total Number of Deaths |
| --- | --- | --- | --- | --- | --- |
| Afghanistan | 1 | 12,218,383.50 | 1,644,757.00 | 3,068,969.00 | 102,556.50 |
| Afghanistan | 4 | 9,128,640.00 | 1,124,521.50 | 2,096,065.00 | 69,979.50 |
| Afghanistan | 14 | 5,057,916.50 | 659,168.00 | 1,228,499.50 | 40,910.50 |
| Afghanistan | 49 | 10,687,758.50 | 1,379,971.50 | 2,571,195.50 | 85,767.00 |
| Angola | 1 | 10,568,601.50 | 1,337,285.50 | 2,495,053.00 | 82,996.50 |
| Angola | 4 | 7,951,658.50 | 958,470.50 | 1,790,227.00 | 59,579.00 |
| Angola | 14 | 3,787,000.00 | 480,525.00 | 904,767.50 | 29,918.00 |
| Angola | 49 | 9,243,635.50 | 1,153,108.50 | 2,151,983.50 | 71,542.50 |
| United Arab Emirates | 1 | 3,070,769.50 | 350,542.50 | 651,357.50 | 21,779.50 |
| United Arab Emirates | 4 | 2,177,762.50 | 214,356.00 | 402,281.50 | 13,375.50 |
| United Arab Emirates | 14 | 1,024,344.50 | 89,813.50 | 167,419.00 | 5,562.50 |
| United Arab Emirates | 49 | 2,602,624.00 | 273,597.50 | 513,621.00 | 17,056.00 |
| Argentina | 1 | 16,773,844.50 | 7,129,191.50 | 13,331,842.50 | 443,779.00 |
| Argentina | 4 | 12,167,825.00 | 4,923,903.50 | 9,183,093.00 | 306,424.50 |
| Argentina | 14 | 7,206,744.00 | 3,179,286.50 | 5,930,434.50 | 197,212.00 |
| Argentina | 49 | 14,559,346.00 | 6,161,078.00 | 11,508,389.50 | 383,318.00 |
| Burundi | 1 | 3,825,576.00 | 487,746.00 | 912,820.50 | 30,294.00 |
| Burundi | 4 | 2,875,191.50 | 343,198.50 | 643,374.00 | 21,457.50 |
| Burundi | 14 | 1,563,193.50 | 192,616.50 | 359,761.00 | 11,941.00 |
| Burundi | 49 | 3,360,357.50 | 416,410.00 | 783,449.00 | 25,958.50 |
| Benin | 1 | 3,987,007.50 | 610,334.00 | 1,140,852.50 | 37,966.50 |
| Benin | 4 | 3,027,030.50 | 436,070.50 | 813,543.50 | 27,098.50 |
| Benin | 14 | 1,617,554.00 | 243,858.00 | 455,765.00 | 15,180.50 |
| Benin | 49 | 3,517,493.00 | 523,439.50 | 980,737.50 | 32,600.50 |
| Burkina Faso | 1 | 6,855,948.00 | 954,394.50 | 1,781,199.00 | 59,384.50 |
| Burkina Faso | 4 | 5,231,105.00 | 692,106.50 | 1,290,564.50 | 42,964.50 |
| Burkina Faso | 14 | 2,954,321.00 | 421,062.50 | 787,440.50 | 26,089.50 |
| Burkina Faso | 49 | 6,035,205.50 | 826,512.00 | 1,546,412.50 | 51,439.50 |
| Bangladesh | 1 | 57,227,126.50 | 14,236,787.50 | 26,575,364.00 | 885,268.50 |
| Bangladesh | 4 | 43,270,144.00 | 10,038,443.00 | 18,753,926.00 | 623,994.50 |
| Bangladesh | 14 | 23,696,468.50 | 5,910,527.00 | 11,038,186.50 | 366,350.50 |
| Bangladesh | 49 | 50,407,176.00 | 12,324,088.50 | 23,001,526.50 | 766,395.00 |
| Bahrain | 1 | 547,191.50 | 80,485.50 | 150,131.00 | 5,024.50 |
| Bahrain | 4 | 391,589.50 | 50,957.50 | 94,643.50 | 3,172.50 |
| Bahrain | 14 | 197,184.50 | 23,694.50 | 43,975.00 | 1,460.00 |
| Bahrain | 49 | 468,646.00 | 64,898.00 | 121,082.00 | 3,992.00 |
| Bolivia | 1 | 3,881,885.00 | 929,770.50 | 1,734,888.50 | 57,656.00 |
| Bolivia | 4 | 2,855,102.00 | 614,416.00 | 1,146,287.00 | 38,054.00 |
| Bolivia | 14 | 1,246,487.00 | 279,998.50 | 519,996.50 | 17,368.00 |
| Bolivia | 49 | 3,364,052.00 | 778,136.00 | 1,449,732.50 | 48,330.00 |
| Brazil | 1 | 76,815,441.00 | 27,418,042.50 | 51,183,618.50 | 1,705,997.00 |
| Brazil | 4 | 56,285,085.00 | 18,283,101.50 | 34,119,544.00 | 1,139,191.50 |
| Brazil | 14 | 30,752,013.00 | 10,628,491.00 | 19,851,592.50 | 658,619.00 |
| Brazil | 49 | 66,729,481.50 | 23,242,138.50 | 43,395,705.00 | 1,445,675.00 |
| Botswana | 1 | 765,573.00 | 159,460.50 | 296,628.50 | 9,918.00 |
| Botswana | 4 | 565,035.50 | 107,208.00 | 199,542.00 | 6,659.50 |
| Botswana | 14 | 305,081.50 | 61,841.00 | 114,201.00 | 3,788.50 |
| Botswana | 49 | 662,701.50 | 134,417.50 | 250,989.00 | 8,333.50 |
| Central African Republic | 1 | 1,542,576.00 | 227,256.00 | 422,653.00 | 14,107.50 |
| Central African Republic | 4 | 1,168,850.50 | 158,951.50 | 297,690.50 | 9,899.50 |
| Central African Republic | 14 | 696,606.50 | 102,635.00 | 192,340.00 | 6,435.00 |
| Central African Republic | 49 | 1,353,904.00 | 194,598.50 | 363,300.50 | 12,130.00 |
| Chile | 1 | 7,194,690.00 | 3,077,838.50 | 5,737,576.50 | 191,424.00 |
| Chile | 4 | 5,194,730.50 | 2,043,884.50 | 3,820,076.00 | 126,898.00 |
| Chile | 14 | 3,037,288.00 | 1,252,044.50 | 2,330,419.00 | 77,706.50 |
| Chile | 49 | 6,244,492.50 | 2,596,331.50 | 4,850,755.50 | 161,525.00 |
| Cote d'Ivoire | 1 | 8,535,011.50 | 1,316,578.00 | 2,457,105.50 | 82,052.50 |
| Cote d'Ivoire | 4 | 6,298,091.50 | 916,128.00 | 1,708,562.00 | 57,155.50 |
| Cote d'Ivoire | 14 | 2,781,975.00 | 429,891.00 | 803,017.00 | 26,661.00 |
| Cote d'Ivoire | 49 | 7,403,108.50 | 1,119,330.00 | 2,085,969.00 | 69,716.00 |
| Cameroon | 1 | 8,729,175.00 | 1,249,335.00 | 2,335,084.00 | 77,781.00 |
| Cameroon | 4 | 6,604,562.00 | 889,631.00 | 1,661,757.00 | 55,432.00 |
| Cameroon | 14 | 3,552,194.00 | 503,335.50 | 939,579.50 | 31,264.50 |
| Cameroon | 49 | 7,690,769.50 | 1,078,672.50 | 2,014,896.00 | 67,062.00 |
| Congo, Dem. Rep. | 1 | 28,914,534.50 | 4,131,602.50 | 7,713,653.50 | 257,149.00 |
| Congo, Dem. Rep. | 4 | 21,729,089.00 | 2,925,815.50 | 5,459,743.00 | 181,724.50 |
| Congo, Dem. Rep. | 14 | 10,339,604.50 | 1,480,493.00 | 2,755,736.00 | 91,222.50 |
| Congo, Dem. Rep. | 49 | 25,274,207.00 | 3,537,679.50 | 6,601,660.50 | 219,628.00 |
| Congo, Rep. | 1 | 1,819,209.50 | 284,222.00 | 531,531.00 | 17,714.00 |
| Congo, Rep. | 4 | 1,380,686.00 | 201,095.00 | 376,298.50 | 12,564.00 |
| Congo, Rep. | 14 | 828,849.00 | 128,923.50 | 241,162.00 | 8,084.50 |
| Congo, Rep. | 49 | 1,603,300.00 | 245,240.00 | 457,933.50 | 15,290.00 |
| Colombia | 1 | 18,341,207.00 | 6,156,013.50 | 11,493,003.50 | 382,744.50 |
| Colombia | 4 | 13,426,478.00 | 4,130,466.00 | 7,696,118.00 | 256,667.50 |
| Colombia | 14 | 7,192,837.50 | 2,342,406.50 | 4,372,127.00 | 144,932.50 |
| Colombia | 49 | 15,964,752.50 | 5,231,315.00 | 9,754,859.00 | 325,054.00 |
| Comoros | 1 | 290,557.00 | 47,011.00 | 87,852.50 | 2,911.50 |
| Comoros | 4 | 215,334.50 | 32,916.00 | 61,230.50 | 2,047.00 |
| Comoros | 14 | 114,051.50 | 18,550.50 | 34,354.00 | 1,129.00 |
| Comoros | 49 | 252,887.00 | 40,321.50 | 75,628.00 | 2,512.00 |
| Cabo Verde | 1 | 181,545.00 | 41,378.50 | 77,437.00 | 2,565.50 |
| Cabo Verde | 4 | 128,006.00 | 26,548.00 | 49,137.00 | 1,638.00 |
| Cabo Verde | 14 | 67,260.50 | 14,657.00 | 27,550.00 | 904.50 |
| Cabo Verde | 49 | 153,456.00 | 33,984.50 | 63,202.50 | 2,102.00 |
| Costa Rica | 1 | 1,822,572.50 | 652,845.00 | 1,218,193.50 | 40,619.00 |
| Costa Rica | 4 | 1,326,970.50 | 428,626.50 | 799,723.50 | 26,658.50 |
| Costa Rica | 14 | 745,696.00 | 252,692.50 | 470,210.00 | 15,640.50 |
| Costa Rica | 49 | 1,578,679.00 | 549,464.00 | 1,023,618.50 | 34,173.50 |
| Cuba | 1 | 4,423,054.50 | 2,120,624.00 | 3,955,835.50 | 131,915.50 |
| Cuba | 4 | 3,216,773.00 | 1,414,337.00 | 2,640,812.00 | 87,972.00 |
| Cuba | 14 | 1,810,327.00 | 857,054.50 | 1,601,201.00 | 53,185.50 |
| Cuba | 49 | 3,831,103.00 | 1,804,660.00 | 3,363,770.00 | 111,899.50 |
| Djibouti | 1 | 343,723.00 | 73,381.50 | 136,983.50 | 4,544.00 |
| Djibouti | 4 | 261,053.50 | 51,856.50 | 97,124.00 | 3,227.50 |
| Djibouti | 14 | 142,168.00 | 30,426.50 | 55,879.50 | 1,876.50 |
| Djibouti | 49 | 302,937.00 | 63,302.00 | 118,347.50 | 3,926.00 |
| Dominican Republic | 1 | 3,807,776.50 | 1,127,809.00 | 2,105,556.50 | 70,140.00 |
| Dominican Republic | 4 | 2,808,434.50 | 765,172.00 | 1,422,811.50 | 47,526.00 |
| Dominican Republic | 14 | 1,562,725.00 | 456,252.50 | 850,089.00 | 28,342.50 |
| Dominican Republic | 49 | 3,315,475.00 | 964,871.00 | 1,796,265.00 | 59,882.00 |
| Algeria | 1 | 14,034,752.50 | 3,466,814.50 | 6,462,517.50 | 215,452.50 |
| Algeria | 4 | 9,901,969.50 | 2,171,943.50 | 4,050,558.50 | 135,157.00 |
| Algeria | 14 | 5,054,134.00 | 1,144,758.00 | 2,139,220.50 | 70,839.50 |
| Algeria | 49 | 11,937,307.00 | 2,835,057.50 | 5,295,541.50 | 175,988.50 |
| Ecuador | 1 | 6,238,253.50 | 1,828,486.50 | 3,416,054.50 | 113,769.50 |
| Ecuador | 4 | 4,681,265.50 | 1,256,086.50 | 2,337,135.50 | 77,960.00 |
| Ecuador | 14 | 2,586,022.00 | 739,506.00 | 1,376,652.00 | 45,789.00 |
| Ecuador | 49 | 5,480,200.00 | 1,567,059.00 | 2,923,489.50 | 97,213.50 |
| Egypt, Arab Rep. | 1 | 32,689,306.00 | 7,149,997.50 | 13,352,307.50 | 444,818.00 |
| Egypt, Arab Rep. | 4 | 23,251,249.50 | 4,594,336.00 | 8,576,020.00 | 285,393.00 |
| Egypt, Arab Rep. | 14 | 11,840,378.50 | 2,461,238.50 | 4,607,142.50 | 152,674.00 |
| Egypt, Arab Rep. | 49 | 27,975,740.50 | 5,946,558.50 | 11,123,014.50 | 370,198.00 |
| Eritrea | 1 | 1,155,784.50 | 209,554.50 | 391,035.00 | 13,020.50 |
| Eritrea | 4 | 871,719.50 | 146,793.00 | 273,048.00 | 9,126.00 |
| Eritrea | 14 | 463,730.00 | 85,068.00 | 156,997.50 | 5,297.00 |
| Eritrea | 49 | 1,011,765.50 | 180,018.50 | 336,576.50 | 11,223.50 |
| Ethiopia | 1 | 37,424,354.50 | 6,123,829.50 | 11,438,760.00 | 380,821.00 |
| Ethiopia | 4 | 28,272,226.50 | 4,318,604.50 | 8,066,304.00 | 268,718.00 |
| Ethiopia | 14 | 14,996,879.50 | 2,457,011.50 | 4,593,451.50 | 152,670.50 |
| Ethiopia | 49 | 32,804,209.50 | 5,277,772.50 | 9,861,570.50 | 328,539.50 |
| Gabon | 1 | 752,439.50 | 127,913.00 | 239,230.50 | 7,959.50 |
| Gabon | 4 | 572,425.50 | 89,835.00 | 168,146.00 | 5,614.00 |
| Gabon | 14 | 347,308.00 | 58,094.50 | 109,709.00 | 3,636.00 |
| Gabon | 49 | 664,127.50 | 110,378.00 | 206,161.50 | 6,867.50 |
| Ghana | 1 | 10,378,959.00 | 1,826,469.00 | 3,406,631.50 | 113,607.50 |
| Ghana | 4 | 7,713,893.50 | 1,273,479.00 | 2,379,059.00 | 79,389.50 |
| Ghana | 14 | 3,423,372.00 | 602,269.50 | 1,120,060.00 | 37,243.50 |
| Ghana | 49 | 9,039,576.00 | 1,557,543.50 | 2,901,100.00 | 96,752.50 |
| Guinea | 1 | 4,258,486.00 | 700,272.50 | 1,311,263.50 | 43,626.50 |
| Guinea | 4 | 3,197,000.00 | 509,156.50 | 950,868.50 | 31,664.00 |
| Guinea | 14 | 1,702,114.50 | 304,197.50 | 565,934.00 | 18,715.50 |
| Guinea | 49 | 3,718,928.00 | 614,330.50 | 1,144,140.00 | 38,187.50 |
| Gambia, The | 1 | 787,667.00 | 120,449.50 | 224,341.50 | 7,490.50 |
| Gambia, The | 4 | 592,870.50 | 88,337.00 | 163,947.00 | 5,442.50 |
| Gambia, The | 14 | 324,941.50 | 53,736.50 | 101,076.00 | 3,349.50 |
| Gambia, The | 49 | 686,599.00 | 104,927.00 | 196,707.00 | 6,559.00 |
| Guinea-Bissau | 1 | 646,616.00 | 100,305.50 | 186,821.00 | 6,207.50 |
| Guinea-Bissau | 4 | 490,671.00 | 72,272.00 | 135,491.00 | 4,505.50 |
| Guinea-Bissau | 14 | 284,320.50 | 45,433.50 | 83,768.50 | 2,782.00 |
| Guinea-Bissau | 49 | 569,959.50 | 87,200.50 | 163,160.00 | 5,405.50 |
| Equatorial Guinea | 1 | 471,118.00 | 62,021.00 | 116,073.50 | 3,858.00 |
| Equatorial Guinea | 4 | 359,112.50 | 44,812.00 | 83,461.50 | 2,771.50 |
| Equatorial Guinea | 14 | 194,873.50 | 25,338.00 | 47,642.00 | 1,593.50 |
| Equatorial Guinea | 49 | 416,196.50 | 53,836.00 | 100,579.00 | 3,348.50 |
| Guatemala | 1 | 5,931,816.00 | 1,188,377.50 | 2,215,138.50 | 73,895.00 |
| Guatemala | 4 | 4,445,955.00 | 809,410.50 | 1,509,951.00 | 50,397.50 |
| Guatemala | 14 | 2,471,211.50 | 477,647.50 | 888,300.00 | 29,575.50 |
| Guatemala | 49 | 5,200,461.00 | 1,004,988.50 | 1,873,662.50 | 62,445.50 |
| Honduras | 1 | 3,368,666.00 | 711,831.50 | 1,328,438.50 | 44,068.00 |
| Honduras | 4 | 2,554,279.00 | 498,194.50 | 928,828.50 | 30,884.00 |
| Honduras | 14 | 1,508,914.50 | 311,765.50 | 579,275.50 | 19,367.50 |
| Honduras | 49 | 2,966,095.00 | 611,292.00 | 1,145,329.00 | 38,068.50 |
| Haiti | 1 | 3,844,733.00 | 864,007.50 | 1,611,689.00 | 53,684.00 |
| Haiti | 4 | 2,891,883.50 | 602,645.50 | 1,127,804.50 | 37,538.50 |
| Haiti | 14 | 1,628,745.00 | 363,202.00 | 678,225.00 | 22,680.00 |
| Haiti | 49 | 3,378,437.50 | 746,954.00 | 1,392,144.50 | 46,506.50 |
| India | 1 | 485,771,554.00 | 134,139,960.00 | 250,304,012.50 | 8,338,437.50 |
| India | 4 | 365,933,321.50 | 93,048,830.00 | 173,656,181.50 | 5,785,606.50 |
| India | 14 | 217,279,232.00 | 58,353,362.00 | 108,949,367.00 | 3,620,179.00 |
| India | 49 | 427,295,820.50 | 115,025,840.50 | 214,808,061.50 | 7,154,678.00 |
| Iran, Islamic Rep. | 1 | 27,824,797.00 | 7,261,103.00 | 13,561,427.50 | 451,739.50 |
| Iran, Islamic Rep. | 4 | 19,593,066.00 | 4,553,642.50 | 8,499,919.50 | 282,846.00 |
| Iran, Islamic Rep. | 14 | 11,174,742.50 | 2,592,113.50 | 4,837,163.50 | 161,457.50 |
| Iran, Islamic Rep. | 49 | 23,673,028.50 | 5,957,133.50 | 11,119,019.00 | 370,270.00 |
| Iraq | 1 | 13,378,016.00 | 2,160,705.50 | 4,036,576.50 | 134,703.50 |
| Iraq | 4 | 10,055,987.00 | 1,513,885.00 | 2,832,294.00 | 94,396.50 |
| Iraq | 14 | 5,444,675.50 | 850,809.00 | 1,586,860.50 | 52,768.00 |
| Iraq | 49 | 11,745,266.50 | 1,852,021.00 | 3,459,984.50 | 115,381.50 |
| Israel | 1 | 3,220,231.50 | 1,398,523.50 | 2,612,412.50 | 86,885.00 |
| Israel | 4 | 2,318,309.00 | 960,045.00 | 1,794,150.50 | 59,726.50 |
| Israel | 14 | 1,417,351.00 | 644,261.00 | 1,206,315.50 | 39,883.00 |
| Israel | 49 | 2,790,877.00 | 1,206,535.00 | 2,251,874.50 | 74,860.00 |
| Jordan | 1 | 3,230,120.50 | 545,182.00 | 1,014,537.00 | 33,793.00 |
| Jordan | 4 | 2,403,684.50 | 360,587.00 | 671,528.00 | 22,395.50 |
| Jordan | 14 | 1,441,001.50 | 226,240.50 | 421,434.00 | 14,019.50 |
| Jordan | 49 | 2,802,081.50 | 455,065.50 | 851,418.00 | 28,348.50 |
| Kenya | 1 | 17,932,717.50 | 3,334,486.00 | 6,220,435.00 | 207,231.00 |
| Kenya | 4 | 13,972,380.00 | 2,685,062.50 | 5,015,623.00 | 166,727.00 |
| Kenya | 14 | 8,920,886.50 | 1,777,160.50 | 3,304,638.00 | 110,213.50 |
| Kenya | 49 | 15,940,230.00 | 3,027,794.00 | 5,654,038.00 | 188,428.00 |
| Cambodia | 1 | 5,249,630.00 | 1,075,358.00 | 2,009,206.50 | 67,003.00 |
| Cambodia | 4 | 3,735,696.50 | 699,366.00 | 1,307,951.50 | 43,432.50 |
| Cambodia | 14 | 1,491,662.00 | 285,706.00 | 532,484.00 | 17,649.50 |
| Cambodia | 49 | 4,455,701.50 | 883,329.50 | 1,656,497.50 | 55,016.50 |
| Kuwait | 1 | 1,365,100.50 | 250,497.00 | 468,191.50 | 15,520.50 |
| Kuwait | 4 | 927,501.50 | 153,854.00 | 289,037.50 | 9,618.00 |
| Kuwait | 14 | 463,836.00 | 72,077.00 | 134,599.00 | 4,499.00 |
| Kuwait | 49 | 1,135,226.00 | 198,574.50 | 372,262.50 | 12,352.50 |
| Lebanon | 1 | 2,473,141.50 | 849,633.00 | 1,578,978.00 | 52,778.00 |
| Lebanon | 4 | 1,838,520.50 | 596,389.00 | 1,111,151.00 | 36,992.00 |
| Lebanon | 14 | 1,166,531.50 | 410,943.00 | 767,150.00 | 25,627.50 |
| Lebanon | 49 | 2,165,967.50 | 736,594.00 | 1,374,880.00 | 45,888.00 |
| Liberia | 1 | 1,679,614.00 | 288,953.00 | 537,754.50 | 17,919.00 |
| Liberia | 4 | 1,277,408.00 | 204,826.00 | 383,901.50 | 12,778.00 |
| Liberia | 14 | 740,784.00 | 127,259.00 | 237,192.00 | 7,905.00 |
| Liberia | 49 | 1,478,591.00 | 249,999.00 | 464,731.00 | 15,427.50 |
| Libya | 1 | 2,278,126.50 | 480,466.00 | 897,001.00 | 29,872.50 |
| Libya | 4 | 1,632,674.50 | 312,685.50 | 585,736.00 | 19,495.50 |
| Libya | 14 | 836,827.00 | 167,080.50 | 312,036.00 | 10,330.00 |
| Libya | 49 | 1,947,756.50 | 401,240.50 | 748,905.50 | 24,938.00 |
| Sri Lanka | 1 | 8,020,343.00 | 3,408,469.50 | 6,355,545.00 | 211,681.00 |
| Sri Lanka | 4 | 5,916,153.00 | 2,373,553.00 | 4,436,366.50 | 147,651.50 |
| Sri Lanka | 14 | 3,594,781.50 | 1,558,313.00 | 2,903,474.50 | 96,594.00 |
| Sri Lanka | 49 | 7,016,699.00 | 2,945,770.00 | 5,508,073.00 | 183,971.50 |
| Lesotho | 1 | 696,741.00 | 145,159.00 | 271,320.50 | 9,016.50 |
| Lesotho | 4 | 508,495.00 | 95,194.00 | 178,041.00 | 5,963.00 |
| Lesotho | 14 | 229,199.00 | 44,271.50 | 82,771.50 | 2,796.00 |
| Lesotho | 49 | 600,007.00 | 121,390.50 | 224,865.00 | 7,490.00 |
| Morocco | 1 | 13,242,616.50 | 4,386,423.50 | 8,186,000.00 | 272,409.00 |
| Morocco | 4 | 9,819,052.50 | 3,059,153.50 | 5,703,002.00 | 190,014.00 |
| Morocco | 14 | 5,899,932.00 | 1,973,055.50 | 3,680,791.50 | 122,030.50 |
| Morocco | 49 | 11,587,125.00 | 3,784,741.00 | 7,071,203.00 | 235,508.50 |
| Madagascar | 1 | 9,150,387.00 | 1,448,349.50 | 2,707,202.50 | 90,125.50 |
| Madagascar | 4 | 6,782,766.00 | 1,015,311.50 | 1,897,325.50 | 63,300.50 |
| Madagascar | 14 | 3,564,206.50 | 563,427.50 | 1,049,184.00 | 35,009.50 |
| Madagascar | 49 | 7,966,524.50 | 1,244,623.50 | 2,321,257.00 | 77,440.00 |
| Mexico | 1 | 46,024,528.50 | 13,801,739.50 | 25,788,652.50 | 858,167.00 |
| Mexico | 4 | 34,417,215.50 | 9,459,831.00 | 17,652,017.50 | 588,526.00 |
| Mexico | 14 | 21,097,465.50 | 6,179,136.00 | 11,554,982.50 | 384,123.00 |
| Mexico | 49 | 40,449,264.50 | 11,850,318.50 | 22,128,971.50 | 737,192.50 |
| Mali | 1 | 6,261,538.50 | 750,361.50 | 1,399,912.50 | 46,770.00 |
| Mali | 4 | 4,479,076.50 | 497,581.50 | 928,693.00 | 30,937.00 |
| Mali | 14 | 2,225,009.00 | 265,778.50 | 494,221.50 | 16,416.00 |
| Mali | 49 | 5,359,818.00 | 623,713.50 | 1,164,621.00 | 38,817.00 |
| Mozambique | 1 | 10,132,075.50 | 1,434,436.00 | 2,675,563.50 | 89,192.00 |
| Mozambique | 4 | 7,529,591.00 | 1,000,297.00 | 1,869,916.50 | 62,413.00 |
| Mozambique | 14 | 3,939,631.00 | 555,317.50 | 1,035,085.50 | 34,251.00 |
| Mozambique | 49 | 8,820,156.50 | 1,230,586.50 | 2,293,159.50 | 76,324.00 |
| Mauritania | 1 | 1,535,219.00 | 254,585.50 | 475,932.50 | 15,814.50 |
| Mauritania | 4 | 1,165,275.50 | 182,691.50 | 339,829.50 | 11,333.00 |
| Mauritania | 14 | 707,749.50 | 121,283.50 | 227,135.00 | 7,523.50 |
| Mauritania | 49 | 1,353,942.00 | 221,412.00 | 412,271.50 | 13,722.50 |
| Mauritius | 1 | 486,885.50 | 209,370.00 | 390,331.50 | 13,047.50 |
| Mauritius | 4 | 351,304.50 | 138,424.00 | 258,993.50 | 8,630.00 |
| Mauritius | 14 | 214,209.50 | 88,073.00 | 165,625.00 | 5,463.00 |
| Mauritius | 49 | 422,540.50 | 176,489.00 | 329,162.50 | 11,015.50 |
| Malawi | 1 | 6,206,020.50 | 853,281.00 | 1,590,558.50 | 52,895.50 |
| Malawi | 4 | 4,609,105.50 | 597,183.50 | 1,114,078.50 | 37,180.00 |
| Malawi | 14 | 2,414,871.50 | 329,973.00 | 616,215.00 | 20,405.50 |
| Malawi | 49 | 5,394,534.00 | 731,214.50 | 1,362,680.00 | 45,337.50 |
| Namibia | 1 | 805,968.50 | 113,124.50 | 211,467.00 | 7,031.50 |
| Namibia | 4 | 594,974.50 | 78,583.00 | 146,712.50 | 4,861.00 |
| Namibia | 14 | 271,679.50 | 36,560.50 | 68,778.50 | 2,296.50 |
| Namibia | 49 | 696,764.50 | 95,833.50 | 179,065.50 | 5,974.50 |
| Niger | 1 | 7,479,840.50 | 911,995.00 | 1,704,345.00 | 56,835.50 |
| Niger | 4 | 5,361,960.00 | 604,501.50 | 1,131,297.50 | 37,570.50 |
| Niger | 14 | 2,656,777.50 | 323,438.50 | 603,880.50 | 20,051.50 |
| Niger | 49 | 6,407,713.00 | 758,280.50 | 1,417,828.00 | 47,137.00 |
| Nigeria | 1 | 67,998,172.00 | 10,931,386.50 | 20,404,287.00 | 679,437.50 |
| Nigeria | 4 | 51,622,542.50 | 7,976,219.00 | 14,878,213.00 | 495,446.50 |
| Nigeria | 14 | 28,071,652.50 | 4,656,256.00 | 8,692,204.50 | 289,283.50 |
| Nigeria | 49 | 59,789,002.00 | 9,541,919.50 | 17,807,958.50 | 593,371.50 |
| Nicaragua | 1 | 2,235,573.00 | 500,811.00 | 937,662.50 | 31,193.00 |
| Nicaragua | 4 | 1,671,927.50 | 340,380.50 | 632,680.50 | 21,123.50 |
| Nicaragua | 14 | 917,568.00 | 190,082.50 | 355,496.00 | 11,913.00 |
| Nicaragua | 49 | 1,954,965.00 | 422,935.50 | 790,201.00 | 26,307.50 |
| Nepal | 1 | 9,915,594.50 | 2,499,733.50 | 4,665,603.00 | 155,338.50 |
| Nepal | 4 | 7,426,283.00 | 1,746,579.00 | 3,269,303.00 | 108,873.50 |
| Nepal | 14 | 3,596,042.00 | 931,577.50 | 1,736,856.00 | 57,692.50 |
| Nepal | 49 | 8,680,618.50 | 2,156,185.00 | 4,028,305.50 | 134,340.50 |
| Oman | 1 | 1,535,261.00 | 176,798.50 | 329,421.00 | 10,993.00 |
| Oman | 4 | 1,072,384.00 | 107,046.00 | 199,923.00 | 6,631.00 |
| Oman | 14 | 551,427.00 | 50,614.00 | 93,722.50 | 3,132.50 |
| Oman | 49 | 1,289,638.50 | 138,700.50 | 258,894.00 | 8,678.00 |
| Pakistan | 1 | 71,833,290.50 | 13,106,615.00 | 24,490,690.50 | 816,437.00 |
| Pakistan | 4 | 53,017,172.50 | 8,893,216.00 | 16,571,105.50 | 551,824.00 |
| Pakistan | 14 | 29,435,591.50 | 5,229,562.50 | 9,784,165.00 | 324,959.50 |
| Pakistan | 49 | 62,541,662.00 | 10,974,825.00 | 20,499,024.50 | 682,425.00 |
| Panama | 1 | 1,517,972.50 | 471,387.00 | 879,487.50 | 29,338.50 |
| Panama | 4 | 1,105,635.50 | 311,636.50 | 581,917.50 | 19,376.00 |
| Panama | 14 | 599,862.50 | 178,411.50 | 335,181.00 | 11,120.50 |
| Panama | 49 | 1,313,913.50 | 396,995.50 | 740,170.50 | 24,635.00 |
| Peru | 1 | 11,655,014.00 | 3,620,110.00 | 6,750,234.00 | 224,803.00 |
| Peru | 4 | 8,460,494.50 | 2,403,019.00 | 4,479,917.00 | 148,873.00 |
| Peru | 14 | 4,169,860.50 | 1,246,737.00 | 2,324,155.00 | 77,271.50 |
| Peru | 49 | 10,081,293.50 | 3,039,427.50 | 5,677,001.50 | 188,946.00 |
| Paraguay | 1 | 2,401,204.50 | 602,571.50 | 1,122,937.50 | 37,455.50 |
| Paraguay | 4 | 1,771,743.00 | 398,632.00 | 744,466.50 | 24,878.00 |
| Paraguay | 14 | 880,973.50 | 208,728.50 | 389,898.50 | 12,896.50 |
| Paraguay | 49 | 2,085,255.00 | 508,236.50 | 946,491.00 | 31,507.00 |
| West Bank and Gaza | 1 | 1,594,003.50 | 226,570.50 | 422,739.00 | 14,105.00 |
| West Bank and Gaza | 4 | 1,189,222.50 | 151,665.50 | 281,435.00 | 9,406.50 |
| West Bank and Gaza | 14 | 712,802.50 | 95,452.50 | 176,325.50 | 5,948.50 |
| West Bank and Gaza | 49 | 1,387,924.50 | 190,382.00 | 355,232.00 | 11,799.00 |
| Qatar | 1 | 899,032.50 | 113,926.50 | 212,464.50 | 7,071.00 |
| Qatar | 4 | 641,023.00 | 70,589.50 | 132,073.50 | 4,377.50 |
| Qatar | 14 | 266,723.50 | 26,536.00 | 49,248.00 | 1,647.50 |
| Qatar | 49 | 766,587.00 | 89,823.50 | 168,594.50 | 5,584.50 |
| Rwanda | 1 | 4,260,092.00 | 653,278.50 | 1,221,703.50 | 40,608.00 |
| Rwanda | 4 | 3,212,134.50 | 457,874.00 | 855,829.50 | 28,478.00 |
| Rwanda | 14 | 1,749,170.00 | 255,902.00 | 479,833.50 | 16,021.50 |
| Rwanda | 49 | 3,746,403.00 | 557,316.50 | 1,041,050.00 | 34,717.00 |
| Saudi Arabia | 1 | 11,941,083.50 | 2,049,485.00 | 3,823,720.50 | 127,556.50 |
| Saudi Arabia | 4 | 8,911,360.50 | 1,384,909.50 | 2,587,572.50 | 86,282.50 |
| Saudi Arabia | 14 | 5,395,467.50 | 824,158.50 | 1,538,146.50 | 51,054.00 |
| Saudi Arabia | 49 | 10,445,557.00 | 1,719,131.00 | 3,203,677.50 | 106,843.00 |
| Sudan | 1 | 14,419,983.00 | 2,514,130.50 | 4,680,808.50 | 155,985.00 |
| Sudan | 4 | 10,877,482.50 | 1,772,405.50 | 3,314,864.00 | 110,203.50 |
| Sudan | 14 | 5,777,423.00 | 1,008,371.50 | 1,878,300.50 | 62,797.00 |
| Sudan | 49 | 12,629,479.00 | 2,164,740.50 | 4,038,214.50 | 134,408.50 |
| Senegal | 1 | 5,506,929.50 | 943,941.00 | 1,765,632.50 | 58,633.50 |
| Senegal | 4 | 4,152,655.50 | 688,535.00 | 1,284,545.00 | 42,863.00 |
| Senegal | 14 | 2,285,377.00 | 427,813.00 | 800,324.00 | 26,502.50 |
| Senegal | 49 | 4,809,000.50 | 828,513.50 | 1,548,749.50 | 51,440.50 |
| Sierra Leone | 1 | 2,630,770.00 | 424,739.50 | 793,709.50 | 26,428.50 |
| Sierra Leone | 4 | 1,994,333.00 | 307,273.00 | 574,350.00 | 19,119.50 |
| Sierra Leone | 14 | 1,158,003.00 | 191,212.00 | 360,302.00 | 12,060.50 |
| Sierra Leone | 49 | 2,319,759.00 | 370,961.50 | 690,722.50 | 22,966.50 |
| El Salvador | 1 | 2,258,281.50 | 726,963.50 | 1,354,593.00 | 45,160.50 |
| El Salvador | 4 | 1,664,921.50 | 490,165.50 | 916,503.50 | 30,630.00 |
| El Salvador | 14 | 908,668.00 | 289,893.50 | 538,904.50 | 17,905.50 |
| El Salvador | 49 | 1,965,853.00 | 619,690.50 | 1,156,953.50 | 38,540.00 |
| Somalia | 1 | 5,054,908.00 | 722,092.50 | 1,346,267.50 | 44,904.50 |
| Somalia | 4 | 3,796,053.50 | 508,883.00 | 947,770.50 | 31,691.00 |
| Somalia | 14 | 1,969,685.00 | 281,476.00 | 522,971.00 | 17,450.00 |
| Somalia | 49 | 4,408,720.50 | 619,059.00 | 1,154,447.00 | 38,523.50 |
| South Sudan | 1 | 3,696,526.00 | 575,956.50 | 1,078,140.00 | 35,884.00 |
| South Sudan | 4 | 2,762,595.00 | 406,328.00 | 758,511.00 | 25,346.00 |
| South Sudan | 14 | 1,282,595.00 | 203,062.00 | 381,821.00 | 12,566.00 |
| South Sudan | 49 | 3,220,523.50 | 493,716.50 | 922,740.50 | 30,649.50 |
| Sao Tome and Principe | 1 | 72,283.50 | 11,231.50 | 20,945.00 | 696.00 |
| Sao Tome and Principe | 4 | 54,913.00 | 8,013.50 | 15,109.50 | 500.00 |
| Sao Tome and Principe | 14 | 29,481.50 | 4,448.00 | 8,484.00 | 283.50 |
| Sao Tome and Principe | 49 | 63,572.00 | 9,662.00 | 17,970.50 | 606.50 |
| Eswatini | 1 | 363,364.00 | 62,143.50 | 115,928.00 | 3,869.00 |
| Eswatini | 4 | 266,346.00 | 40,923.50 | 76,890.50 | 2,558.00 |
| Eswatini | 14 | 119,567.50 | 18,886.00 | 35,996.50 | 1,187.00 |
| Eswatini | 49 | 313,533.00 | 51,987.50 | 96,273.50 | 3,204.00 |
| Seychelles | 1 | 35,263.00 | 12,022.50 | 22,569.50 | 746.00 |
| Seychelles | 4 | 25,064.00 | 7,926.50 | 14,905.00 | 495.50 |
| Seychelles | 14 | 14,434.00 | 4,800.50 | 8,982.50 | 294.50 |
| Seychelles | 49 | 30,440.50 | 10,024.50 | 18,709.50 | 625.00 |
| Syrian Arab Republic | 1 | 5,792,480.00 | 989,564.50 | 1,847,430.00 | 61,580.00 |
| Syrian Arab Republic | 4 | 4,370,151.50 | 718,417.00 | 1,339,837.50 | 44,443.00 |
| Syrian Arab Republic | 14 | 2,590,708.00 | 461,948.50 | 861,860.00 | 28,637.00 |
| Syrian Arab Republic | 49 | 5,101,506.50 | 858,728.00 | 1,601,689.50 | 53,348.50 |
| Chad | 1 | 5,065,942.50 | 601,091.00 | 1,124,428.00 | 37,284.50 |
| Chad | 4 | 3,620,549.50 | 399,236.00 | 745,414.50 | 24,835.00 |
| Chad | 14 | 1,795,297.50 | 213,052.50 | 398,405.50 | 13,124.00 |
| Chad | 49 | 4,330,613.50 | 500,212.50 | 933,773.00 | 31,055.00 |
| Togo | 1 | 2,738,321.50 | 409,696.00 | 766,658.50 | 25,515.50 |
| Togo | 4 | 2,084,481.50 | 294,394.50 | 549,644.00 | 18,293.50 |
| Togo | 14 | 1,117,765.50 | 165,863.50 | 310,016.00 | 10,260.50 |
| Togo | 49 | 2,416,837.50 | 353,371.50 | 660,994.00 | 21,999.00 |
| Tajikistan | 1 | 2,980,380.50 | 506,624.00 | 946,025.50 | 31,461.00 |
| Tajikistan | 4 | 2,132,996.50 | 334,988.50 | 624,270.50 | 20,882.50 |
| Tajikistan | 14 | 1,073,616.50 | 173,478.00 | 325,632.50 | 10,814.50 |
| Tajikistan | 49 | 2,558,536.50 | 423,792.00 | 791,527.50 | 26,444.00 |
| Tunisia | 1 | 4,156,065.00 | 1,421,020.00 | 2,652,235.00 | 88,351.50 |
| Tunisia | 4 | 2,993,242.50 | 934,481.50 | 1,744,116.00 | 58,013.50 |
| Tunisia | 14 | 1,711,642.00 | 556,560.50 | 1,039,231.00 | 34,717.50 |
| Tunisia | 49 | 3,578,598.00 | 1,197,555.50 | 2,241,228.00 | 74,470.50 |
| Turkey | 1 | 31,029,721.00 | 11,408,352.00 | 21,265,807.50 | 708,020.00 |
| Turkey | 4 | 23,141,897.50 | 7,967,971.50 | 14,847,503.50 | 495,399.50 |
| Turkey | 14 | 14,342,055.00 | 5,342,495.00 | 9,982,584.00 | 331,482.50 |
| Turkey | 49 | 27,262,980.00 | 9,879,607.50 | 18,427,321.50 | 614,391.00 |
| Tanzania | 1 | 19,439,044.50 | 2,891,250.00 | 5,398,275.50 | 180,022.50 |
| Tanzania | 4 | 14,683,606.50 | 2,076,715.50 | 3,872,581.00 | 129,155.00 |
| Tanzania | 14 | 7,943,080.00 | 1,196,374.50 | 2,231,611.50 | 74,239.50 |
| Tanzania | 49 | 17,106,351.50 | 2,514,752.00 | 4,689,457.00 | 156,667.50 |
| Uganda | 1 | 14,035,584.50 | 2,241,331.50 | 4,179,978.50 | 139,292.00 |
| Uganda | 4 | 10,795,699.00 | 1,767,713.00 | 3,302,956.50 | 109,887.00 |
| Uganda | 14 | 9,634,472.00 | 1,547,767.00 | 2,892,963.50 | 96,319.50 |
| Uganda | 49 | 12,625,196.50 | 2,044,288.00 | 3,816,799.50 | 126,941.00 |
| Uruguay | 1 | 1,326,201.50 | 679,333.50 | 1,267,820.00 | 42,256.00 |
| Uruguay | 4 | 938,231.00 | 445,262.00 | 828,683.00 | 27,689.50 |
| Uruguay | 14 | 516,148.00 | 261,647.50 | 487,979.00 | 16,297.00 |
| Uruguay | 49 | 1,141,786.50 | 570,625.00 | 1,062,322.00 | 35,429.00 |
| Venezuela, RB | 1 | 10,135,488.00 | 3,272,693.50 | 6,112,276.00 | 203,631.50 |
| Venezuela, RB | 4 | 7,385,224.00 | 2,226,216.00 | 4,152,468.50 | 138,283.50 |
| Venezuela, RB | 14 | 4,478,616.50 | 1,444,018.50 | 2,687,084.50 | 89,718.00 |
| Venezuela, RB | 49 | 8,782,756.00 | 2,799,109.50 | 5,232,723.50 | 173,968.50 |
| Yemen, Rep. | 1 | 8,978,939.50 | 1,189,341.00 | 2,223,058.50 | 74,107.50 |
| Yemen, Rep. | 4 | 6,443,445.50 | 752,402.00 | 1,407,658.50 | 46,808.50 |
| Yemen, Rep. | 14 | 3,134,621.00 | 389,461.00 | 723,166.50 | 23,984.50 |
| Yemen, Rep. | 49 | 7,619,708.50 | 969,122.50 | 1,816,360.50 | 60,463.00 |
| South Africa | 1 | 20,484,691.50 | 4,996,026.50 | 9,320,556.00 | 310,571.50 |
| South Africa | 4 | 15,232,036.50 | 3,438,007.00 | 6,410,413.00 | 213,792.00 |
| South Africa | 14 | 8,795,217.00 | 2,079,588.50 | 3,890,084.50 | 129,416.00 |
| South Africa | 49 | 17,929,452.50 | 4,284,328.50 | 7,988,411.00 | 266,130.00 |
| Zambia | 1 | 5,908,589.50 | 719,278.00 | 1,341,163.00 | 44,602.50 |
| Zambia | 4 | 4,464,221.00 | 514,499.50 | 966,758.00 | 32,042.50 |
| Zambia | 14 | 2,130,687.00 | 259,718.00 | 486,323.50 | 16,191.50 |
| Zambia | 49 | 5,177,648.00 | 620,034.50 | 1,154,007.00 | 38,561.00 |
| Zimbabwe | 1 | 4,572,303.50 | 991,498.50 | 1,852,341.50 | 61,683.50 |
| Zimbabwe | 4 | 3,484,742.00 | 817,788.00 | 1,524,034.50 | 50,874.50 |
| Zimbabwe | 14 | 3,275,359.00 | 779,772.50 | 1,450,085.50 | 48,299.00 |
| Zimbabwe | 49 | 4,110,615.00 | 926,563.00 | 1,728,563.00 | 57,739.50 |

## Supplemental S2. Parameters and process used in health resource and costing model

### S2.1. Summary

We summarise here the main parameters used in the estimates of health resources and costing. Further details and references are then provided in the following sections.

In summary, there are four steps in our calculations:

1. Calculation of unit costs per activity for three base countries: Ethiopia (low-income country or ‘LIC’), Pakistan (lower-middle income country or ‘LMIC’) and South Africa (upper-middle income country or ‘UMIC’)
2. Extrapolation of unit costs in base countries to calculate unit costs across 84 LICs, LMICs and UMICs
3. Calculation of total costs per country using country-specific unit costs, modelled data on the number of cases, hospitalisations and deaths, as well as other epidemiological and economic assumptions
4. Calculation of country-specific per capita costs, as well as per capita costs as a proportion of gross domestic product (GDP) per capita and various measures of health expenditure per capita

### S2.2 Calculation of unit costs per activity for three base countries

#### S2.2.1 General Approach

A full economic costing was carried out over a one-year time horizon. Costs were constructed using a bottom-up ingredients-based technique. The costing was carried out from a health systems perspective and included both direct (e.g. medicines) and indirect costs (e.g. facility overheads). No above-service delivery costs were included.

The eighty-four countries chosen met three inclusion criteria: 1) be classified as low-income, lower-middle income or upper-middle income by the World Bank [17], 2) be included in the list of 92 countries for which epidemiological modelling data was available from Pearson et al (2020) [16], and 3) have recent available GDP per capita data in order to carry out cost extrapolation between countries [17].

#### S2.2.2 Intervention costs

We used official WHO guidance to identify areas related to critical preparedness, readiness and response actions for COVID-19 to define a set of interventions involved in a national response to the pandemic. This guidance identifies 6 priority areas of work and is further sub-divided into 13 activities.

- Emergency response mechanisms at the national level
- Risk communication and community engagement
- Case finding, contact tracing and management
- Surveillance
- Public health measures
- Case management

For the first five areas of work we considered only WHO guidance to define the resource use. For case management costs we assumed less resource-intensive activities that we felt were more plausible in low- and middle-income settings (‘real-world’). Assumptions on ‘real world’ resource use were based on the clinical expertise of members of the research team and are detailed below.

Following this guidance, we generated a list of unit costs to be brought together with the COVID epidemiological model to estimate resource needs, as detailed in Table S2.2.

**Table S2.2.2.1.** Intervention description and unit costs

| Activity | Activity unit costs |
| --- | --- |
| 1.a. Emergency Response Mechanisms: National level | Per country per day |
| 1.b. Emergency Response Mechanisms: Training of health staff | One-off per site |
| 2. Risk communication & community engagement | Per country per day |
| 3.a. Case finding, contact tracing and management: Contact tracing | Per person contacted |
| 3.b. Case finding, contact tracing and management: Quarantine of contacts | Per person quarantined |
| 4.a. Surveillance: Case notification | Per positive case |
| 4.b. Surveillance: Reporting (national level) | Per country per week |
| 5. Public health measures: Hygiene education | Per education campaign per month |
| 6. Screening and diagnosis | Per person screened and tested |
| 7.a. Case Management: Home-based care | Per person requiring home-based care |
| 7.b. Case Management: Hospital-based (severe case) | Per day of hospitalisation (severe case) |
| 7.c. Case Management: Hospital-based (critical case) | Per day of hospitalisation (critical case) |
| 7.d. Case Management: Death | Per COVID-related death |

#### S.2.2.3. Defining Inputs, Costs per Input and Quantities of Inputs

For each of the abovementioned activities we used an ingredients based costing to identify a series of inputs (e.g. junior-level government worker day). For each input we estimated a number of units (e.g. three days of work) and a country-specific price. The costs of each input were identified using a range of sources, according to availability of recent primary cost data and appropriateness of cost estimates to the COVID pattern of care. See the table below.

**Table S2.2.3.1.** Quantities and Unit Costs per Input per Activity per Country

| Component | Number of<br>Units per<br>Input | Unit Cost per Inputs |  |  |
| --- | --- | --- | --- | --- |
|  |  | Ethiopia | Pakistan | South Africa |
| 1. a. Emergency Response Mechanisms: National level |  |  |  |  |
| Working day (junior level govt) | 10 | \$ 12.27 | \$ 13.07 | \$ 194.66 |
| Working day (senior level govt) | 10 | \$ 17.29 | \$ 23.94 | \$ 256.72 |
| Meeting/ training costs per person per day | 20 | \$ 13.18 | \$ 20.44 | \$ 159.17 |
| Total: | | \$ 559.26 | \$ 778.90 | \$ 7,697.16 |
| 1. b. Emergency Response Mechanisms: Training of health staff |  |  |  |  |
| Working day (health care workers) | 250 | \$ 4.93 | \$ 10.43 | \$ 97.58 |
| Working day (junior level govt) | 10 | \$ 12.27 | \$ 13.07 | \$ 194.66 |
| Working day (senior level govt) | 1 | \$ 17.29 | \$ 23.94 | \$ 256.72 |
| Meeting/ training costs per person per day | 261 | \$ 13.18 | \$ 20.44 | \$ 159.17 |
| Total: | | \$ 4,813.58 | \$ 8,096.53 | \$ 68,141.36 |
| 2. Risk communication & community engagement |  |  |  |  |
| Working day (junior level govt) | 3 | \$ 12.27 | \$ 13.07 | \$ 194.66 |
| Working day (senior level govt) | 2 | \$ 17.29 | \$ 23.94 | \$ 256.72 |
| Media costs per day | 1 | \$ 2.74 | \$ 4.58 | \$ 36.00 |
| Total: | | \$ 74.14 | \$ 91.67 | \$ 1,133.44 |
| 3. a. Case finding, contact tracing and management: Contact tracing |  |  |  |  |
| Working day (junior level govt) | 0.1 | \$ 12.27 | \$ 13.07 | \$ 194.66 |
| Contact tracing household visit | 0.33 | \$ 2.08 | \$ 3.02 | \$ 13.68 |
| Contact tracing phone call | 0.67 | \$ 2.34 | \$ 0.34 | \$ 3.31 |
| Total: | | \$ 3.48 | \$ 2.54 | \$ 26.23 |
| 3. b. Case finding, contact tracing and management: Quarantine of contacts |  |  |  |  |
| Working day (health care workers) | 0.1 | \$ 4.93 | \$ 10.43 | \$ 97.58 |
| Working day (junior level govt) | 0.1 | \$ 12.27 | \$ 13.07 | \$ 194.66 |
| Total: | | \$ 1.72 | \$ 2.35 | \$ 29.22 |
| 4. a. Surveillance: Case notification |  |  |  |  |
| Working day (health care workers) | 0.1 | \$ 4.93 | \$ 10.43 | \$ 97.58 |
| Working day (junior level govt) | 0.1 | \$ 12.27 | \$ 13.07 | \$ 194.66 |
| Total: | | \$ 1.72 | \$ 2.35 | \$ 29.22 |
| 4. b. Surveillance: Reporting (national level) |  |  |  |  |
| Working day (health care workers) | 0.5 | \$ 4.93 | \$ 10.43 | \$ 97.58 |
| Working day (junior level govt) | 0.1 | \$ 12.27 | \$ 13.07 | \$ 194.66 |
| Total: | | \$ 3.69 | \$ 6.52 | \$ 68.26 |
| 5. Public health measures: Hygiene education |  |  |  |  |
| Working day (junior level govt) | 2 | \$ 12.27 | \$ 10.43 | \$ 97.58 |
| Working day (senior level govt) | 1 | \$ 17.29 | \$ 13.07 | \$ 194.66 |
| Media costs per day | 1 | \$ 2.74 | \$ 4.58 | \$ 36.00 |
| Total: | | \$ 44.58 | \$ 38.51 | \$ 425.83 |
| 6. Screening and diagnosis |  |  |  |  |
| Ambulance trip | 0.0001 | \$ 4.80 | \$ 9.51 | \$ 60.41 |
| Isolation pod/ diagnostic visit | 2 | \$ 0.49 | \$ 0.49 | \$ 7.97 |
| Outpatient visit oral history | 1 | \$ 3.57 | \$ 0.47 | \$ 8.02 |
| Outpatient visit physical exam | 1 | \$ 3.57 | \$ 0.47 | \$ 8.02 |
| Outpatient visit specimen collection | 1 | \$ 4.88 | \$ 1.09 | \$ 17.15 |
| COVID19 test (PCR) | 1 | \$ 23.98 | \$ 23.98 | \$ 23.98 |
| **Total:** | | \$ 36.97 | \$ 26.98 | \$ 73.12 |
| **7.a. Case Management: Home-based care** |  |  |  |  |
| Home-based care bed-day | 5 | \$ 0.94 | \$ 0.61 | \$ 11.65 |
| Community-based care via GP | 2 | \$ 9.11 | \$ 4.71 | \$ 44.16 |
| **Total:** | | \$ 22.90 | \$ 12.45 | \$ 146.57 |
| **7.b. Case Management: Hospital-based (severe case)** |  |  |  |  |
| Inpatient ward bed-day (severe) | 1 | \$ 29.90 | \$ 31.54 | \$ 96.66 |
| Diagnostics |  |  |  |  |
| Pulse oximetry | 0.125 | \$ 0.00 | \$ 0.00 | \$ 0.00 |
| Chest X-ray | 0.125 | \$ 27.35 | \$ 2.79 | \$ 21.86 |
| Full blood count | 0.125 | \$ 2.37 | \$ 2.29 | \$ 24.28 |
| Blood urea and electrolyte test | 0.125 | \$ 4.20 | \$ 2.53 | \$ 2.87 |
| C-reactive protein test | 0.125 | \$ 2.34 | \$ 0.32 | \$ 5.15 |
| HIV test | 0.125 | \$ 4.38 | \$ 3.87 | \$ 17.13 |
| COVID19 test (PCR) | 0 | \$ 23.98 | \$ 23.98 | \$ 23.98 |
| Malaria test | 0.125 | \$ 0.19 | \$ 0.19 | \$ 0.19 |
| Haemoglobin test | 0.125 | \$ 2.29 | \$ 2.29 | \$ 2.29 |
| **Total:** | | \$ 35.29 | \$ 33.32 | \$ 105.88 |
| **7.c. Case Management: Hospital-based (critical case)** |  |  |  |  |
| Inpatient ward bed-day (critical) | 0.33 | 30.65 | 32.29 | 97.41 |
| ITU bed-day | 0.67 | \$ 104.48 | \$ 101.99 | \$ 662.71 |
| Additional resourcing per COVID-related complication |  |  |  |  |
| Acute respiratory distress syndrome (ARDS) | 0.47 | \$ 22.46 | \$ 22.46 | \$ 22.46 |
| Acute kidney injury days | 0.04 | \$ 10.60 | \$ 10.60 | \$ 10.60 |
| Acute cardiac injury days | 0.06 | \$ 46.25 | \$ 46.25 | \$ 46.25 |
| Liver dysfunction days | 0.06 | \$ 89.32 | \$ 89.32 | \$ 89.32 |
| Pneumothorax days | 0.01 | \$ 6.66 | \$ 6.77 | \$ 7.02 |
| Hospital-acquired pneumonia days | 0.05 | \$ 18.85 | \$ 18.85 | \$ 18.85 |
| Bacteraemia days | 0.01 | \$ 32.55 | \$ 32.55 | \$ 32.55 |
| Urinary tract infection days | 0.01 | \$ 9.03 | \$ 9.03 | \$ 9.03 |
| Septic shock days | 0.05 | \$ 0.64 | \$ 0.67 | \$ 0.75 |
| Diagnostics |  |  |  |  |
| Pulse oximetry | 10 | \$ 0.00 | \$ 0.00 | \$ 0.00 |
| Chest X-ray | 10 | \$ 27.35 | \$ 2.79 | \$ 21.86 |
| Full blood count | 10 | \$ 2.37 | \$ 2.29 | \$ 24.28 |
| Blood urea and electrolyte test | 10 | \$ 4.20 | \$ 2.53 | \$ 2.87 |
| C-reactive protein test | 10 | \$ 2.34 | \$ 0.32 | \$ 5.15 |
| Venous blood gas test | 10 | \$ 4.23 | \$ 4.23 | \$ 4.23 |
| HIV test | 0.1 | \$ 4.38 | \$ 3.87 | \$ 17.13 |
| COVID19 test (PCR) | 0 | \$ 23.98 | \$ 23.98 | \$ 23.98 |
| Malaria test | 0.1 | \$ 0.19 | \$ 0.19 | \$ 0.19 |
| Haemoglobin test | 0.1 | \$ 2.29 | \$ 2.29 | \$ 2.29 |
| **Total:** | | \$ 505.56 | \$ 221.18 | \$ 1,081.94 |
| **7.d. Case Management: Death** |  |  |  |  |
| Body Bag | 1 | \$ 64.52 | \$ 64.52 | \$ 64.52 |
| **Total:** | | \$ 64.52 | \$ 64.52 | \$ 64.52 |

#### S.2.2.4. Quantities

##### Planning and Management Activities (Activities 1-5)

Quantities of working days required for planning and management activities were estimated from expert consultation as part of the Diseases Control Priorities project. For other activities, quantities were estimated based on requirements for similar activities for tuberculosis (TB) such as contact tracing from the VALUE TB study and previous studies in South Africa (more below).

##### Diagnosis and Clinical Management (Activities 6–7)

The number of days per patient in general ward and in ICU was set at 8 and 10 respectively, and was set to match the assumptions in the epidemiological model [16, 19, 20]. Following expert clinician advise we assumed that one-third of critical patient bed days would be treated the general ward and two-thirds in the ICU.

The likelihood of additional COVID-related complications (per day) were estimated using evidence on the clinical course of COVID from patients in Wuhan, China^17^, as were assumptions on the duration of symptoms^18^ [21]. The number of diagnostic tests per hospitalisation was carried out in consultation with expert clinicians in essential critical care.

#### S.2.2.5 Input unit costs

##### S2.2.5.1 Estimation of non-bed-day costs (Pakistan)

An ingredients-based approach was used to calculate most of the service costs and prices for Pakistan. The data used was collected as part of the Disease Control Priorities 3-Universal Health Coverage (DCP3-UHC) project. For other countries primary data from the TB studies was used (see below).

For Pakistan, staff-related costs were constructed using federal-level pay scales. For most outputs, the number of minutes of staff required per activity were estimated via expert opinion obtained from clinicians working in the Health Planning, System Strengthening & Information Analysis Unit (HPSIU) in the Ministry of National Health Services Regulations and Coordination of Pakistan. For outputs where this was unavailable, health economists agreed a plausible assumed value.

Drug regimens were costed using resource use data obtained through expert opinion (HPSIU) and a number of price sources. An assessment of strengths and weaknesses of different price sources was conducted and hierarchy of sources was established. The primary source of price data was the Sindh Health Department Procurement Price list. If a price was unavailable, the Federal Wholesale Price List for Generic Medicines was used as a second option. As a last resort, private sector market prices were used.

Cost on supplies and equipment were similarly constructed. Resource use was determined through expert opinion (HPSIU) and price source hierarchy established. The primary source was the Medical Emergency Resilience Fund 2019–2020, and a secondary source was private sector market prices.

For all countries, for additional diagnostic and radiology costs (beyond those available from the TB data) were estimated using available literature and market prices. We assessed strengths and weaknesses of different price sources. For example, we used the ‘Costing and Pricing of Services in Private Hospitals of Lahore: Summary Report’ as our primary source as it contained a methodological appendix that suggested that an ingredients-based approach consistent with ours was followed. If some prices were unavailable we used user fees from the Pakistan Institute of Medical Sciences, procurement prices from the Medical Emergency Resilience Fund procurement prices and user fees from the Aga Khan University Hospital. Space costs were estimated using data from budget documents from the Federal government (Islamabad Capital Territory Health Infrastructure PC-1).

Oxygen therapy costs per bed-day were calculated by estimating the number of cylinders consumed in 24 hours at different flow rates, assumed to be 10L per minute in the general ward and 30L per minute in the ICU. Cylinder duration (hours) was estimated by dividing pressure by the number of litres per minute, assuming a standard cylinder size of 4.6kg, filled at 1,900 psi pressure [22]. Cost per cylinder was obtained from the South African online catalogue of a manufacturer that is active in both South Africa and Pakistan [23].

##### S2.2.5.2 Estimation of non-bed-day costs (Ethiopia and South Africa)

For Ethiopia the main source of cost data was the VALUE TB study. Cost data collected from a health provider perspective to estimate the economic costs of TB-related health services. Full costs of health services were estimated, reflecting their implementation in the ‘real world’ rather than using a ‘per protocol’ costing approach. Cost data collection was retrospective, over a one-year period to minimize the risk of bias due to seasonality. Resource use was measured in the VALUE TB study using both top-down and bottom-up methods wherever possible, to allow for comparison. The costs included in the current cost model reflected an average of top-down and bottom-up costs by site.

For South Africa, we used primary data from the XTEND trial (nurses and lay health workers)^19^. This was updated using more recent data on contact tracing and update prices collected on an on-going trial of TB preventative therapy (WHIP_3_TB) (http://ClinicalTrials.gov, 2020).

Some of the COVID-19 interventions were outside the scope of the VALUE TB, XTEND, and WHIP_3_TB studies. Values for which a primary unit cost was partially or entirely unavailable from Value TB are listed in the Table below. For these interventions, resource use from Pakistan (DCP) was then used with local prices (see list below).

**Table S.2.2.5.3.1.** Inputs for which unit costs were estimated for South Africa and Ethiopia using resource use from DCP

|  |
| --- |
| Planning & coordination activities |
| Working day (mid-level facility) |
| Working day (junior level govt) |
| Working day (senior level govt) |
| Meeting/ training costs per day |
| Media costs per day |
| Health hotline (day) |
| **Infection control** |
| Ambulance trip |
| Isolation pod/ diagnostic visit |
| Deep clean |
| **Home-based care** |
| Home-based care bed-day |
| community-based care via GP |
| **Inpatient treatment** |
| Inpatient ward bed-day including PPE (normative scenario) |
| ICU bed-day, including PPE (normative scenario) |
| Severe case ward bed-day, including PPE |
| Critical case ward bed-day, including PPE |
| ITU bed-day ('real-world scenario') |
| Body disposal |
| **Additional resourcing per COVID related complication** |
| Acute respiratory distress syndrome (ARDS) |
| Acute kidney injury |
| Acute cardiac injury |
| Liver dysfunction |
| Pneumothorax |
| Hospital-acquired pneumonia |
| Bacteraemia |
| Urinary tract infection |
| Septic shock |
| **Investigations (lab tests)** |
| Pulse oximetry |
| Venous blood gas |
| Mid-stream urine test |
| CoVID confirmatory lab test (PCR) |
| Malaria |
| Haemoglobin |

Where costs were applied from Pakistan health care inputs were classified as either tradeable, as in the case of medical equipment and supplies for diagnostic tests and procedures, and non-tradeable, including non-medical supplies, capital and overhead costs. For tradable inputs, where country-specific price estimates were not available from primary data or from the published literature, the estimate from Pakistan was applied to other countries. For non-tradable inputs, the estimate from Pakistan was adjusted by an amount reflecting the difference in the two countries’ GDP (PPP). The rationale behind this approach is that, while exchange rate may be influenced by government policy, PPP seeks to equalise the purchasing power of different currencies and, as such, may better reflect differentials in salaries and other non-tradable prices across countries.

**Table S2.2.5.4.** Relative GDP adjustment factors

|  |  |  | Relative GDP (PPP) |  |  |  |  |
| --- | --- | --- | --- | --- | --- | --- | --- |
| | Exchange rate (US\$) | GDP per capita by country (US\$ PPP) | Pakistan | South Africa | Kenya | India | Ethiopia |
| Pakistan | 155.00 | 5,567.06 | 1.00 | 0.41 | 0.87 | 0.72 | 2.75 |
| South Africa | 32.26 | 2,022.14 | 2.46 | 1.00 | 3.73 | 1.76 | 6.77 |
| Kenya | 76.92 | 7,762.88 | 0.62 | 0.25 | 1.00 | 0.45 | 1.71 |
| India | 104.17 | 3,467.56 | 1.39 | 0.57 | 1.18 | 1.00 | 3.84 |
| Ethiopia | 16.95 | 13,686.88 | 0.36 | 0.15 | 0.45 | 0.26 | 1.00 |

##### S2.2.5.5 Estimation of bed-day costs (all countries)

We took an ingredients approach to estimating the costs of general ward and ICU ward bed days, as these were major cost drivers in our cost model. We estimated the plausible number of nursing hours per bed day in an LMIC setting through consultation with members of the research team who have expertise in critical care in LMICs. In ICU the assumption of nurse to patient ratio would be 1:1; in the general ward the ratio would be 1:6 during the day time and 1:20 in the night.

To understand the full range of inputs required we obtained the underlying costing data set provided by the authors of a recent primary costing of critical carer^20^. The paper reports the results of a detailed activity-based costing in a hospital in Karachi, disaggregated by phase of care. We used the cost data for the ward stay phase, removing any supplies or equipment specific to the surgery, to estimate the average generic costs of a bed-day.

All bed-day costs were compared to and validated against available country-specific estimates from the published literature and from ongoing research and WHO CHOICE (see Table S2.4). Rapid literature searches were conducted on the Medline, Embase and EconLit databases on 8^th^ and 9^th^ April 2020 to identify records reporting on the costs of ICU care in each of the study countries (see table below).

**Table S2.2.5.5.1.** Comparison of bed-day costs by setting

| | Unit cost (2019 US\$) | | |
| --- | --- | --- | --- |
|  | Extrapolated | From literature | Source |
| Kenya | 362.49 | 349.25 | (Lalani et al., 2018) |
| India | 512.44 | 585.80 | (Mathai et al., 2015) |
| South Africa | 1240.96 | 1644.27 | (Mahomed and Mahomed, 2019) |

We estimated the additional costs of ICU beds compared to standard hospital beds using an ingredients-based approach to cost the equipment and supplies not present in standard hospital wards. We used the procurement price of equipment and assumed depreciation over ten (ventilators and suction pumps) or five years (all other equipment). Supply costs included central and arterial lines, ventilator tubing, and sedatives.

##### S2.2.5.6 COVID specific costs

Finally, we calculated the costs of personal protective equipment (PPE) per health worker per day (see Table below) and allocated a cost per PPE per minute to clinical staff. We also calculated costs of hygiene per bed day. We estimated the costs of PPE and hygiene supplies using a list of necessary supplies from a COVID-related budget from the Ministry of Health of Pakistan, which included local prices sourced by the Aga Kahn University. This was complemented for other countries using the WHO’s Essential Supplies Forecasting Tool (ESFT) [7]. We divided supplies into single-use and disposable. We determined plausible quantities and useful life for supplies following clinical guidelines and expert opinion.

**Table S2.2.5.6.1.**
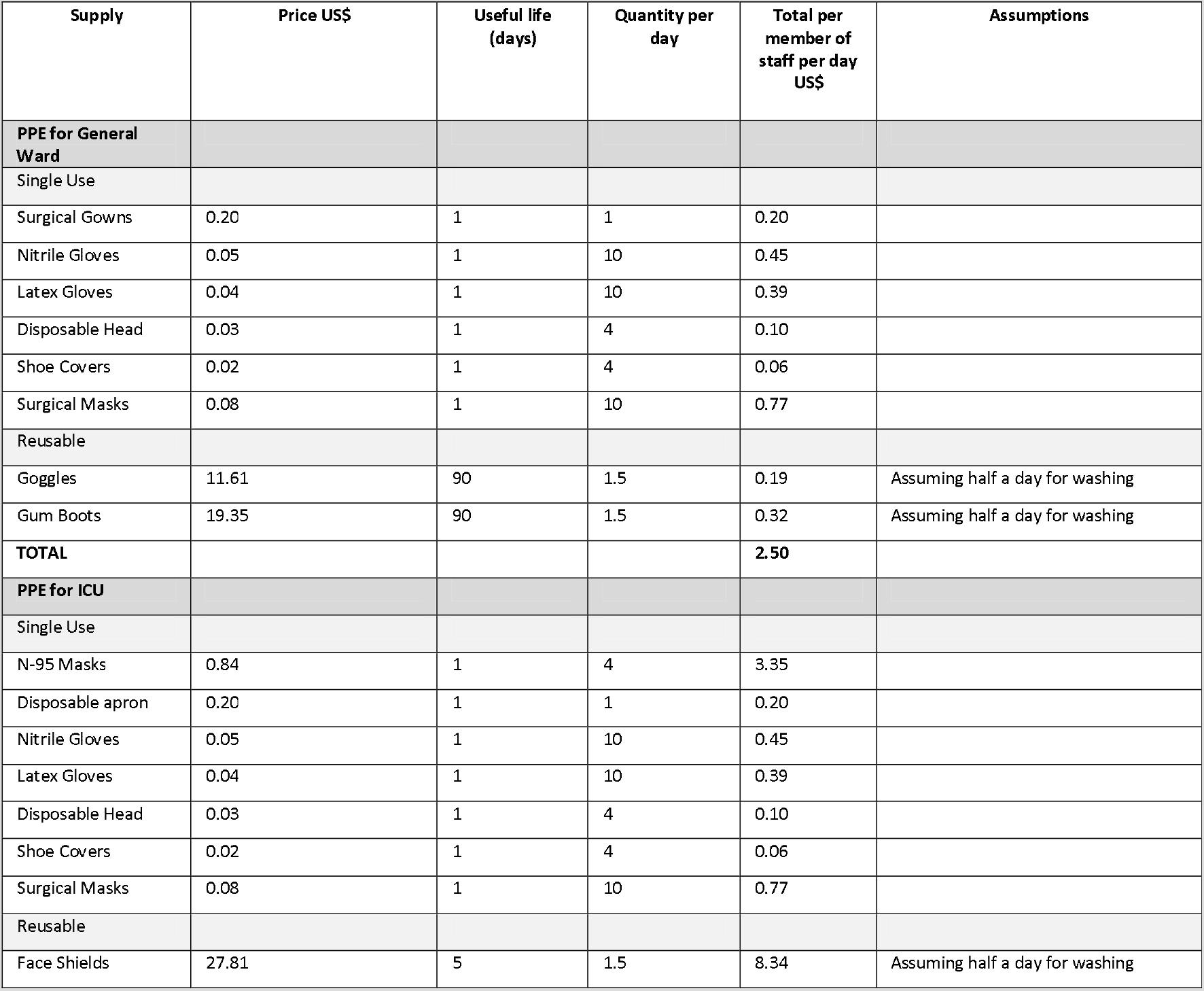
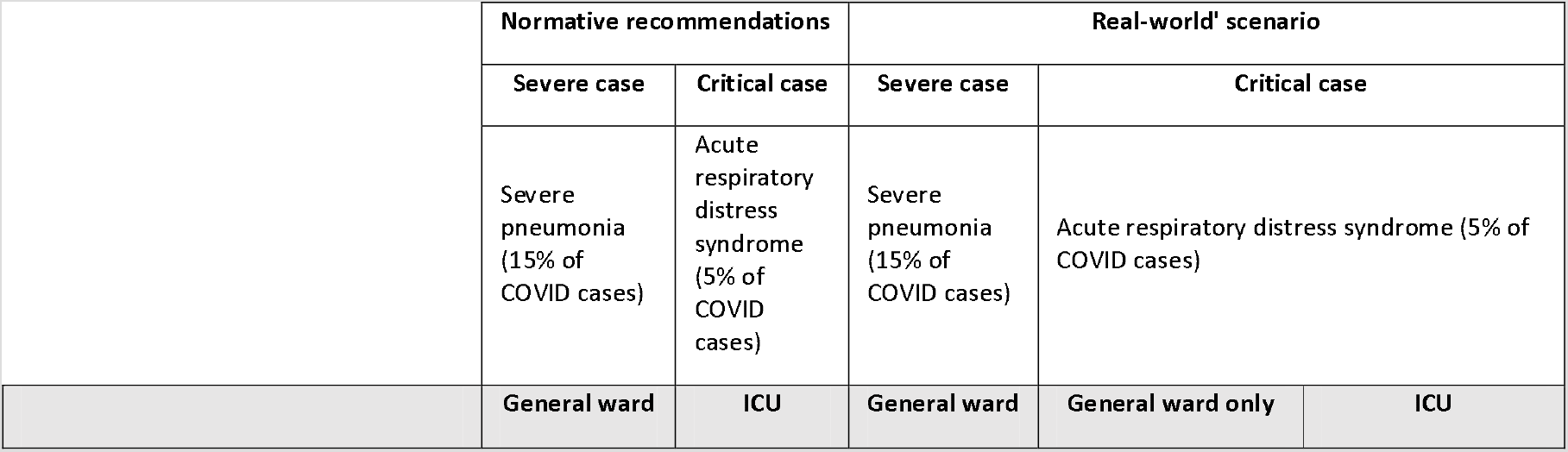
PPE costs per general ward bed day and per ICU bed day

**Table S2.2.5.6.2.**
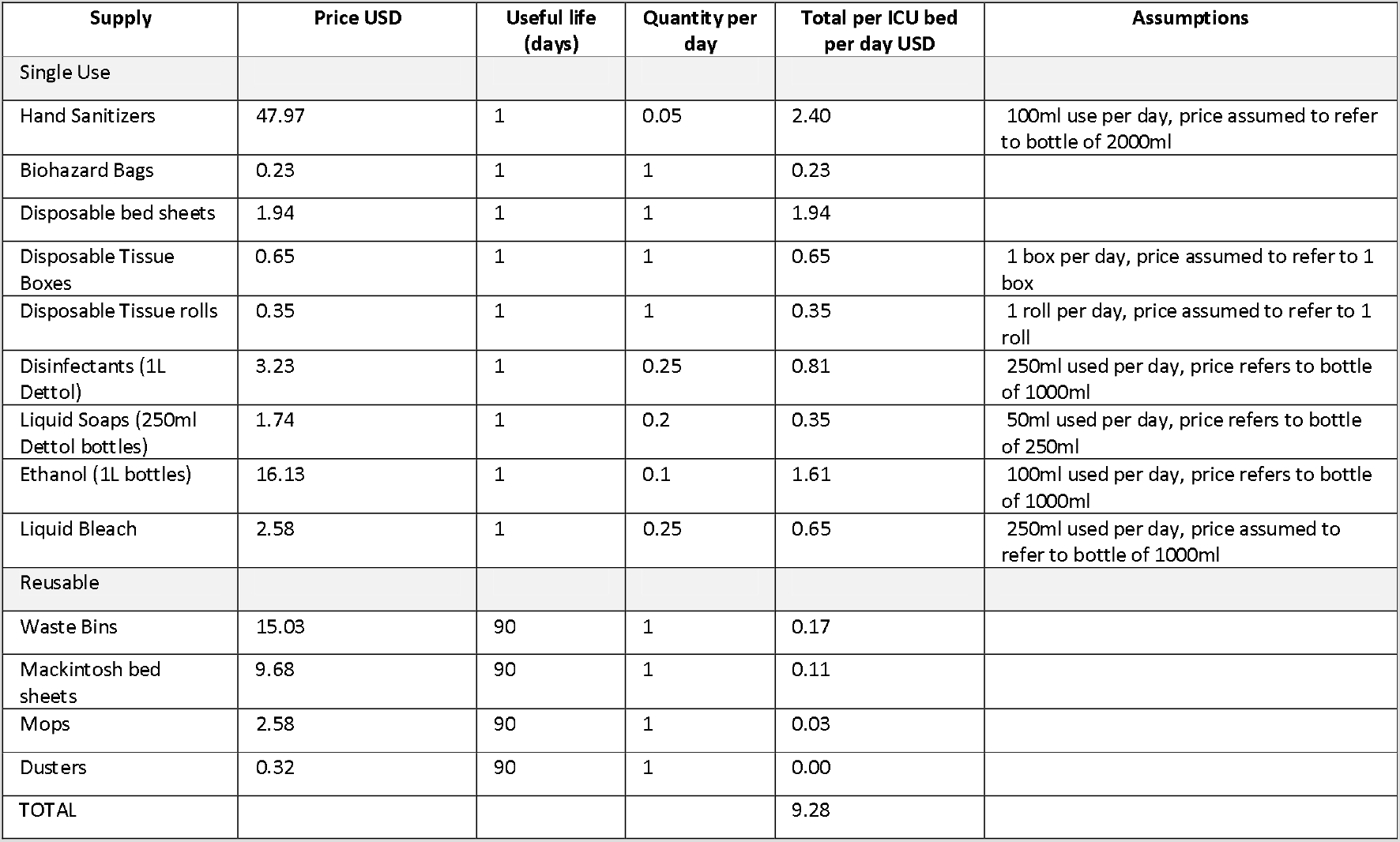
Hygiene costs per General Ward and ICU bed day

| Supply | Price USD | Useful life (days) | Quantity per day | Total per ICU bed per day USD | Assumptions |
| --- | --- | --- | --- | --- | --- |
| Single Use |  |  |  |  |  |
| Hand Sanitizers | 47.97 | 1 | 0.05 | 2.40 | 100ml use per day, price assumed to refer to bottle of 2000ml |
| Biohazard Bags | 0.23 | 1 | 1 | 0.23 |  |
| Disposable bed sheets | 1.94 | 1 | 1 | 1.94 |  |
| Disposable Tissue Boxes | 0.65 | 1 | 1 | 0.65 | 1 box per day, price assumed to refer to 1 box |
| Disposable Tissue rolls | 0.35 | 1 | 1 | 0.35 | 1 roll per day, price assumed to refer to 1 roll |
| Disinfectants (1L Dettol) | 3.23 | 1 | 0.25 | 0.81 | 250ml used per day, price refers to bottle of 1000ml |
| Liquid Soaps (250ml Dettol bottles) | 1.74 | 1 | 0.2 | 0.35 | 50ml used per day, price refers to bottle of 250ml |
| Ethanol (1L bottles) | 16.13 | 1 | 0.1 | 1.61 | 100ml used per day, price refers to bottle of 1000ml |
| Liquid Bleach | 2.58 | 1 | 0.25 | 0.65 | 250ml used per day, price assumed to refer to bottle of 1000ml |
| Reusable |  |  |  |  |  |
| Waste Bins | 15.03 | 90 | 1 | 0.17 |  |
| Mackintosh bed sheets | 9.68 | 90 | 1 | 0.11 |  |
| Mops | 2.58 | 90 | 1 | 0.03 |  |
| Dusters | 0.32 | 90 | 1 | 0.00 |  |
| **TOTAL** |  |  |  | **9.28** |  |

Oxygen supplementation therapy is the main form of treatment for COVID 19. There are different methods of oxygen delivery which utilise different types of supplies, equipment and require different average levels of oxygen flow. We calculated costs for 6 types of oxygen delivery techniques and assumed a distribution across severe and critical patients according to members of our research team with clinical expertise in critical care in LMICs. See Table below.

**Table S2.2.5.6.3.**
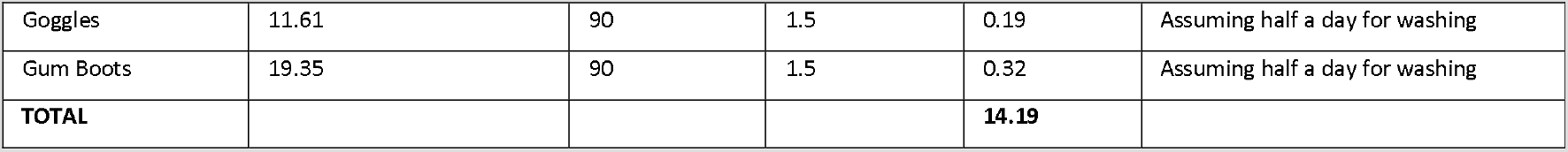
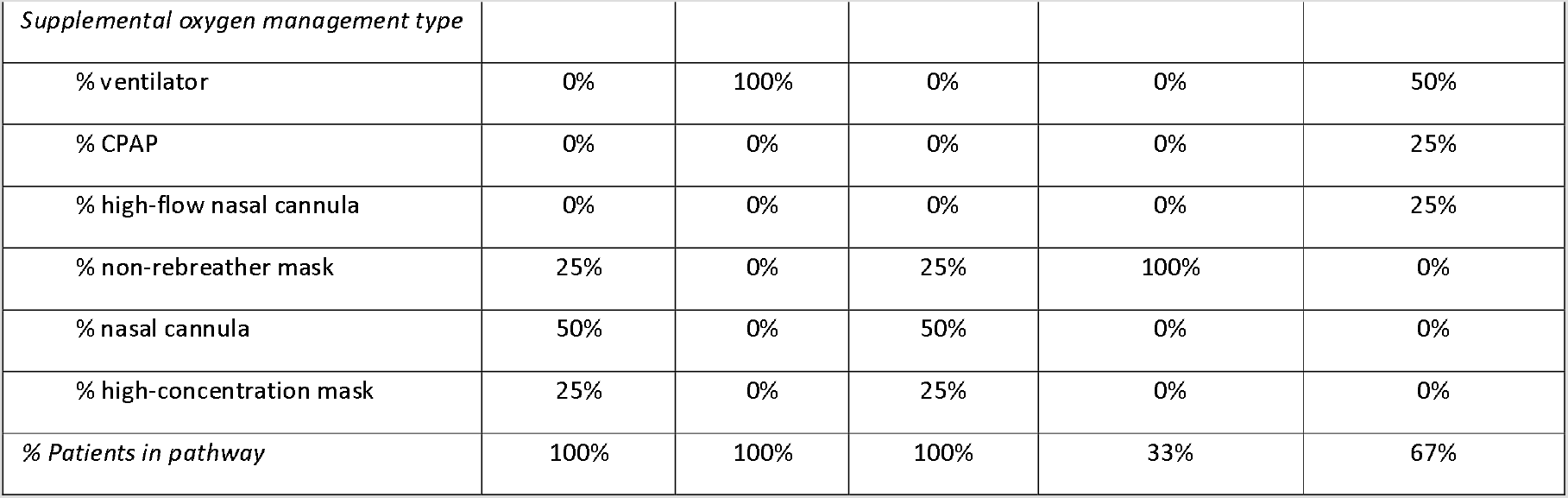

### S2.3 Extrapolation of unit costs in base countries to calculate unit costs across 84 LICs, LMICs and UMICs

We used the unit costs obtained in our three base countries to extrapolate unit costs to other LICs, LMICs and UMICs. We grouped countries according to income group. Costs for LICs were extrapolated using unit costs from Ethiopia, costs for LMICs were extrapolated from the unit costs from Pakistan, and those for UMICs from the unit costs from South Africa.

In order to carry out the extrapolation, each cost ingredient for each of the unit costs was classified as a tradeable good, non-tradeable good, or staff cost.

Tradeable goods are generally defined as those that can easily be traded in the international market and include goods such as medical or other supplies and medications. The unit costs for our three base countries were initially converted from each local currency into 2019 US$ using market exchange rates. To convert the tradeable good from the base country (e.g. Ethiopia) to a ‘second’ country (e.g. Afghanistan) we apportioned the percentage of the unit cost that was composed of tradeable goods in 2019 US$ from the base country to the second country.

To convert staff costs from a base country to a second country we used conversion rates from Serje et al (2018) [18]. Serje et al (2018) use regression analysis on a dataset containing wages from health workers of different skill levels for 193 countries in order to predict wages by country income level relative to GDP per capita. We used the multiples per GDP per capita presented in the paper in order to convert the staff wages from the base country to the second country. See the table from Serje et al (2018) below.

**Table S2.3.1.** Health worker earnings as a multiple of GDP per capita (Serje et al 2018)

| **World bank income categories** | **Health worker cadre** | **Average earnings index**<br>**(multiple of GDP per capita)** |
| --- | --- | --- |
| High-income countries | Physicians | 1.9 |
|  | Nurses and midwives | 1.5 |
|  | Other health workers | 0.9 |
| Upper-middle income countries | Physicians | 2.7 |
|  | Nurses and midwives | 2.2 |
|  | Other health workers | 1.3 |
| Lower-middle income countries | Physicians | 5.1 |
|  | Nurses and midwives | 4.2 |
|  | Other health workers | 2.4 |
| Lower-income countries | Physicians | 7.8 |
|  | Nurses and midwives | 6.4 |
|  | Other health workers | 3.7 |
| Global | Physicians | 4.4 |
|  | Nurses and midwives | 3.6 |
|  | Other health workers | 2.1 |

### S2.4 Calculation of total costs per country using country-specific unit costs, modelled data on the number of cases, hospitalisations and deaths, as well as other epidemiological and economic assumptions

The unit cost in each of the 84 countries was used to calculate the total costs per activity per country. The table below explains the quantities that those unit costs were multiplied by in order to calculate the total costs per country.

**Table S2.4.1.**

| Activity | Unit Type | Quantities per country | Value | Source |
| --- | --- | --- | --- | --- |
| 1.a. Emergency Response<br>Mechanisms: National level | Per country<br>per day | Number of working days per year | 260 | Assumption |
| 1.b. Emergency Response<br>Mechanisms: Training of health<br>staff | One-off per<br>site | Total number of clinical sites * | Variable per<br>Country | Calculated by assuming<br>one site for every 200<br>hospital beds available in<br>the country [24] |
| 2. Risk communication &<br>community engagement | Per country<br>per day | Number of calendar days per year | 365 | N/A |
| 3.a. Case finding, contact tracing<br>and management: Contact<br>tracing | Per person<br>contacted | Total number of COVID19 cases * | Variable by<br>country | See Table S1.3.1 |
|  |  | % cases that are symptomatic * | 69% | [25] |
|  |  | % of symptomatic cases tested * | 10% | Assumption |
|  |  | Average number of contacts per<br>COVID19-positive case | 7 | [26] |
| 3.b. Case finding, contact tracing<br>and management: Quarantine of<br>contacts | Per person<br>quarantined | Total number of COVID19 cases * | Variable by<br>country | See Table S1.3.1 |
|  |  | % cases that are symptomatic * | 69% | [25] |
|  |  | % of symptomatic cases tested * | 10% | Assumption |
|  |  | Average number of contacts per<br>COVID19-positive case | 7 | [26] |
| 4.a. Surveillance: Case<br>notification | Per positive<br>case | Total number of COVID19 cases * | Variable per<br>Country | See Table S1.3.1 |
|  |  | % cases that are symptomatic * | 69% | [25] |
|  |  | % of symptomatic cases tested * | 10% | Assumption |
| 4.b. Surveillance: Reporting<br>(national level) | Per country<br>per day | Total number of clinical sites * | Variable per<br>Country | Calculated by assuming<br>one site for every 200<br>hospital beds available in<br>the country [24] |
|  |  | Weeks per year | 52 | N/A |
| 5. Public health measures: Hygiene education | Per education campaign | Months per year | 12 | N/A |
| 6. Screening and diagnosis | Per person screened and tested | (Total number of COVID19 cases * | Variable per Country | See Table S1.3.1 |
|  |  | % of cases requiring hospitalisation * | 18.50% | [16, 19] |
|  |  | Number of people tested per positive case) + | 11.31 | See Table 2.4.2 |
|  |  | (Total number of COVID19 cases * | Variable per Country | See Table S1.3.1 |
|  |  | % cases that are symptomatic * | 69% | [25] |
|  |  | % of symptomatic cases tested * | 10% | Assumption |
|  |  | Number of people tested per positive case) | 11.31 | See Table 2.4.2 |
| 7.a. Case Management: Home-based care | Per person requiring home-based care | Proportion of borderline mild-to-severe cases | 10% | Assumption |
| 7.b. Case Management: Hospital-based (severe case) | Per day of hospitalisation (severe case) | Average number of days of hospitalisation for severe cases | 8 | [16, 20] |
| 7.c. Case Management: Hospital-based (critical case) | Per day of hospitalisation (critical case) | Average number of days of hospitalisation for critical cases | 10 | [16, 19] |
| 7.d. Case Management: Death | Per COVID-related death | Total number of deaths from COVID19 | Variable per Country | See Table S1.3.1 |

**Table 2.4.2.** Test positivity rate by country and average

| Country | % of positive tests | Source |
| --- | --- | --- |
| South Africa | 0.169 | [27] |
| Kenya | 0.103 | [26] |
| Ethiopia | 0.0739 | [28] |
| India | 0.0612 | [29] |
| Pakistan | 0.0351 | [30] |
| **Average** | **0.08844** |  |

### S2.5 Calculation of country-specific per capita costs, as well as per capita costs as a proportion of gross domestic product (GDP) per capita and various measures of health expenditure per capita

Total costs per country were used to calculate the COVID-19-related costs per capita per country per scenario by dividing the total costs by the population of the country [17]. The cost per capita was then calculated as a proportion of GDP per capita [17] and three measures of health expenditure per capita [31]: 1) total health expenditure including out-of-pocket payments, 2) total health expenditure excluding out-of-pocket payments, and 3) government health spending per capita. Data on GDP per capita and health expenditure per capita per country can be found in the table below.

**Table S.2.5.1.**
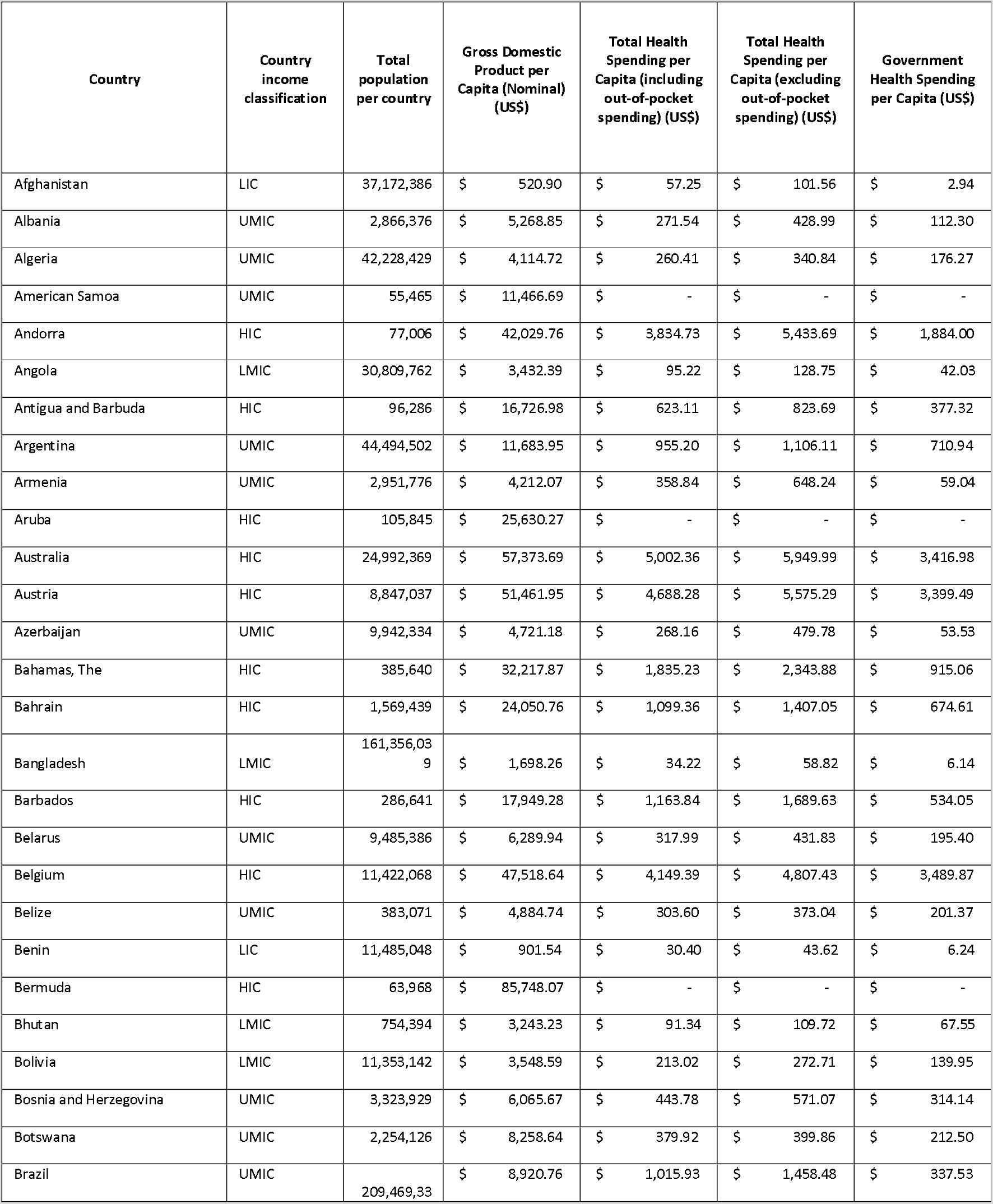
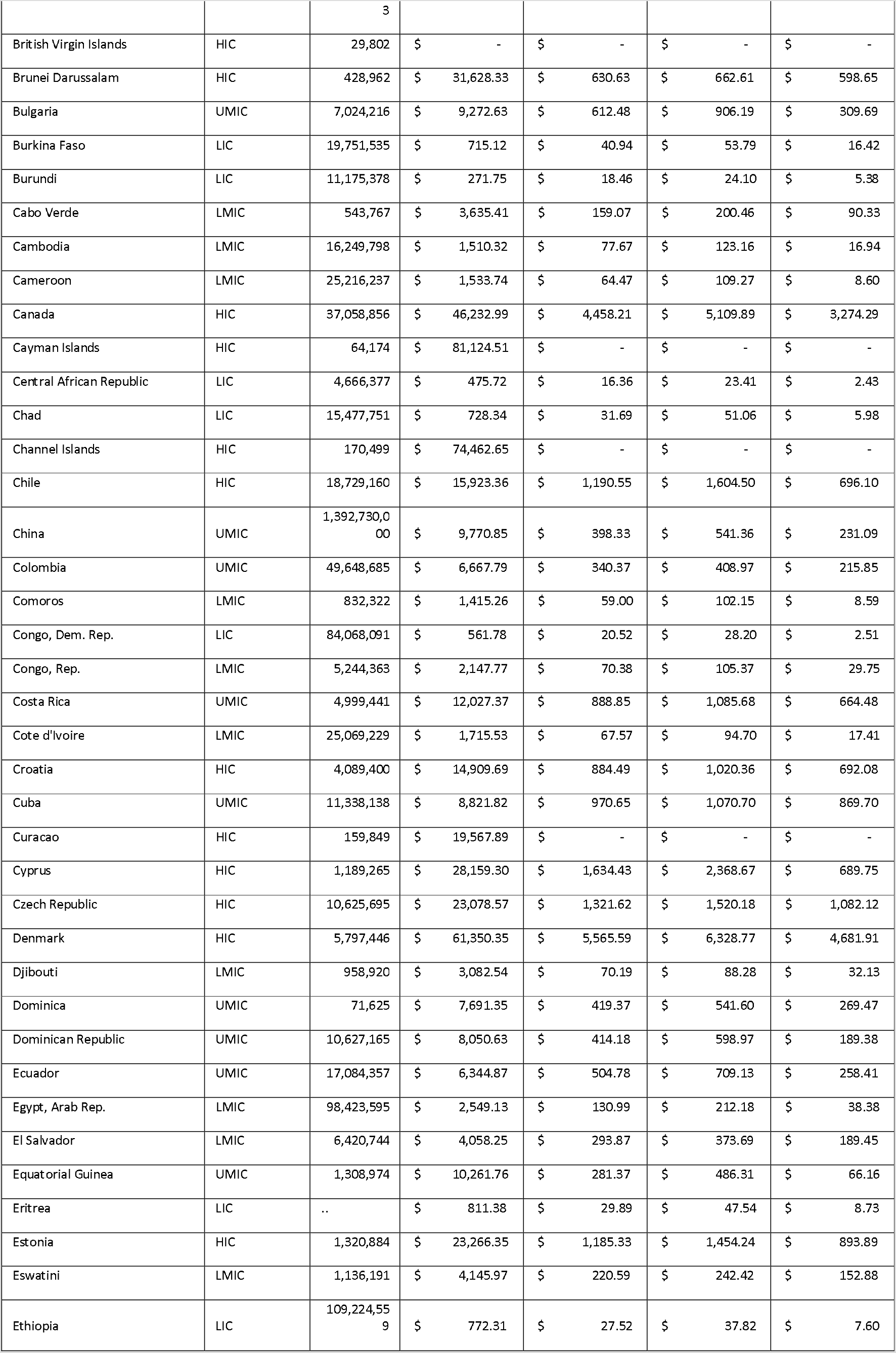
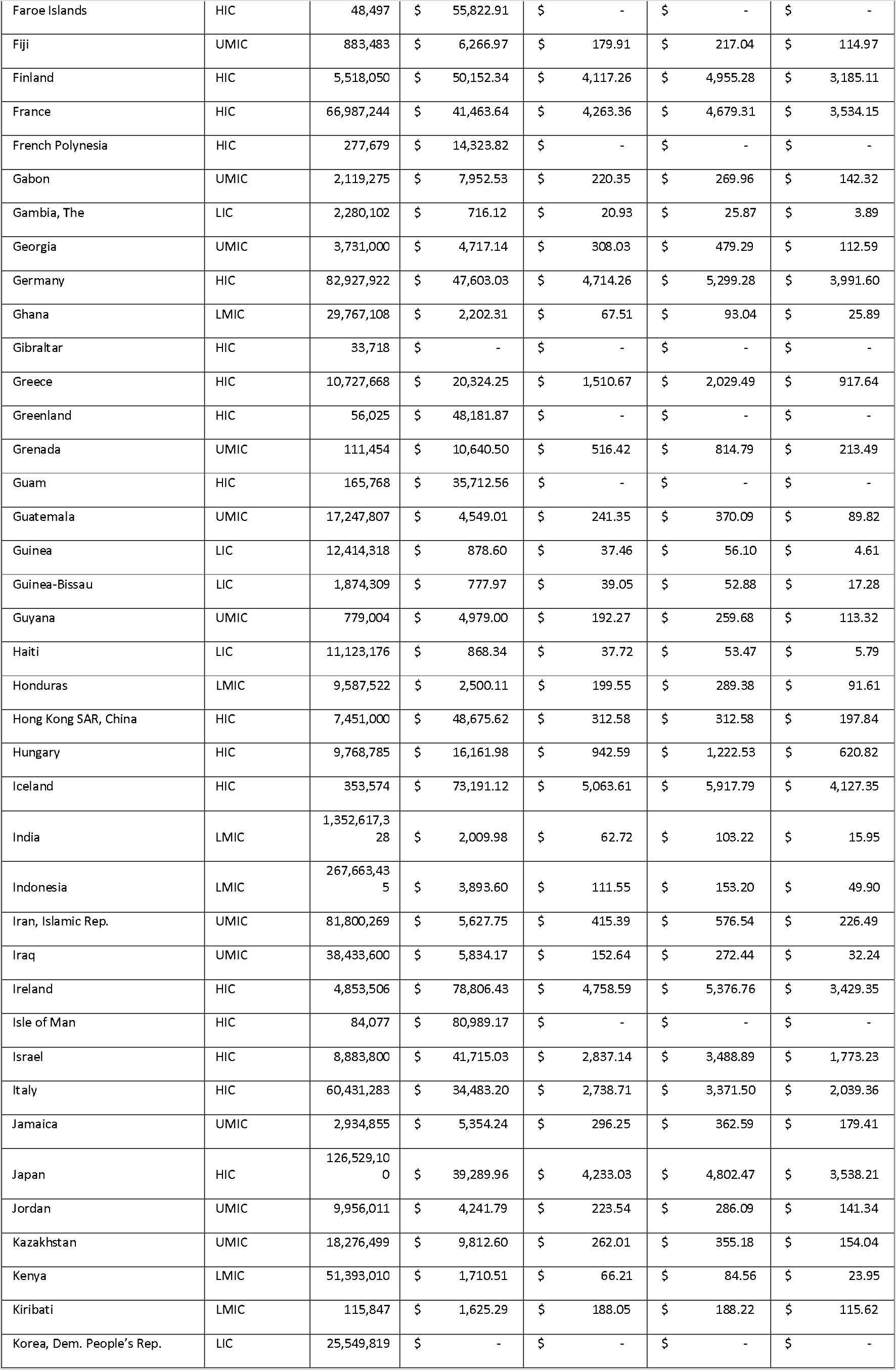
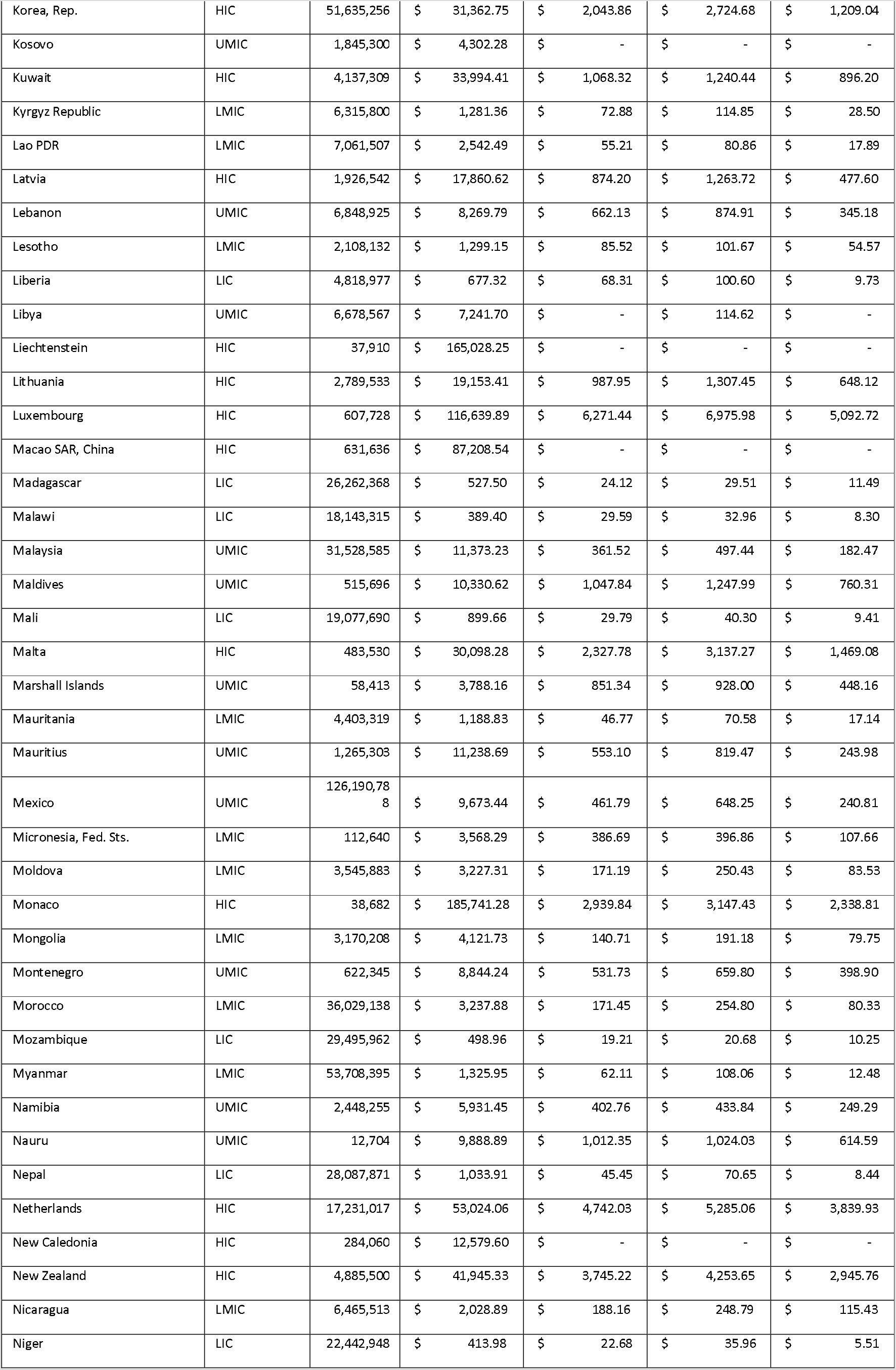
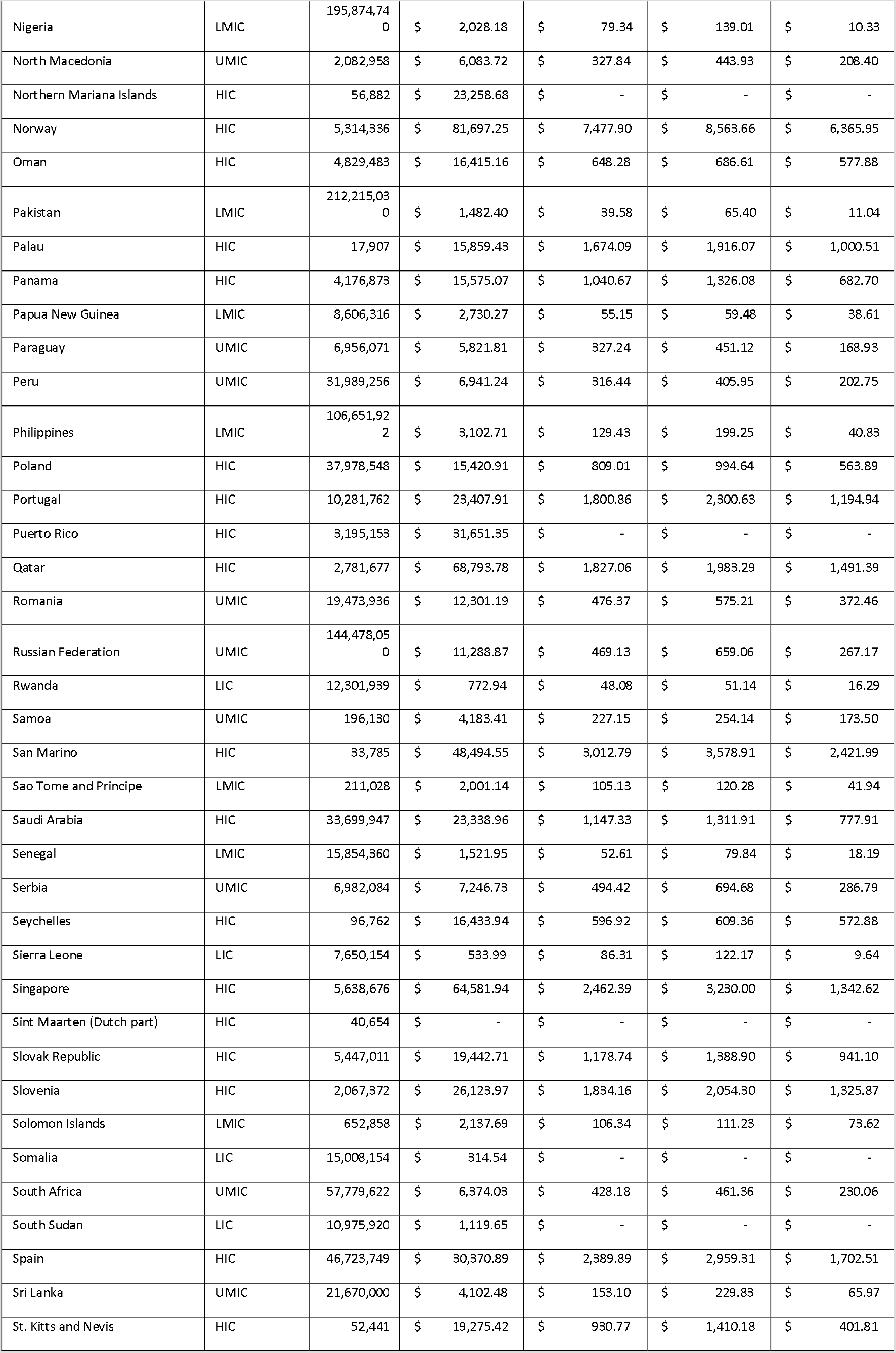
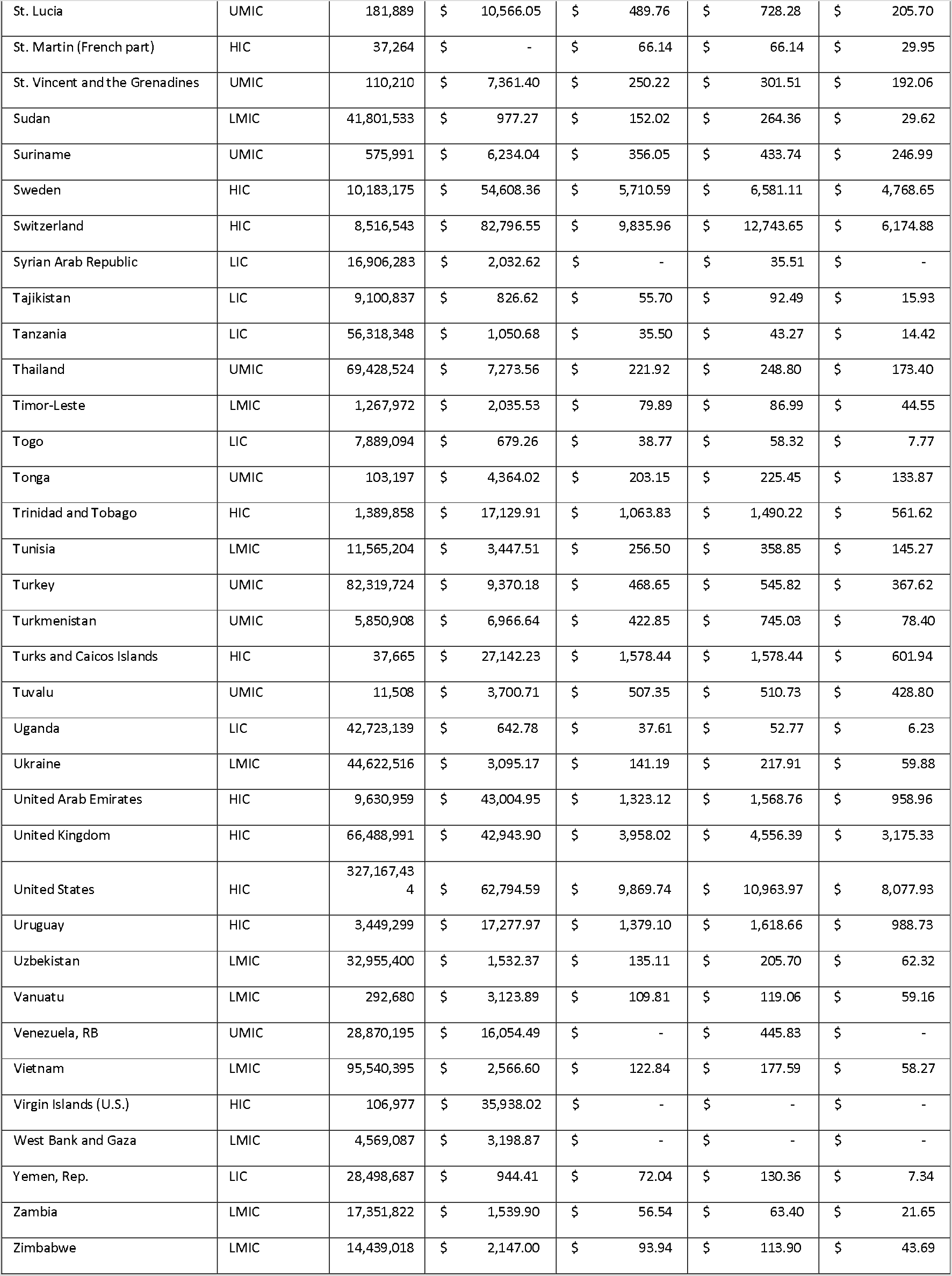
GDP, Health Spending and Government Health Spending

